# The relative effects of non-pharmaceutical interventions on early Covid-19 mortality: natural experiment in 130 countries

**DOI:** 10.1101/2020.10.05.20206888

**Authors:** Jonathan Stokes, Alex James Turner, Laura Anselmi, Marcello Morciano, Thomas Hone

## Abstract

**Background:** Concurrent non-pharmaceutical interventions have been implemented around the world to control Covid-19 transmission. Their general effect on reducing virus transmission is proven, but they can also be negative to mental health and economies, and transmission behaviours can also change in absence of mandated policies. Their relative impact on Covid-19 attributed mortality rates, enabling policy selection for maximal benefit with minimal disruption, is not well established.

**Methods:** We exploited variations in nine non-pharmaceutical interventions implemented in 130 countries (3250 observations) in two periods chosen to limit reverse causality: i) prior to first Covid-19 death (when policymakers could not possibly be reacting to deaths in their own country); and, ii) 14-days-post first Covid-19 death (when deaths were still low, on average). We examined associations with daily deaths per million in each subsequent 24-day period (the time between virus transmission and mortality) which could only be affected by the policy period. A mean score of strictness and timeliness was coded for each intervention. Days in each country were indexed in time by first reported Covid-19 death to proxy for virus transmission rate. Multivariable linear regression models of Covid-19 mortality rates on all concurrent interventions were adjusted for seasonality, potential confounders, and potential cross-country differences in their mortality definitions. Robustness was checked by removing countries with known data reporting issues and with non-linear, negative binomial, models.

**Results:** After adjusting for multiple concurrent interventions and confounders, and accounting for both timing and strictness of interventions, earlier and stricter school (−1.23 daily deaths per million, 95% CI -2.20 -0.27) and workplace closures (−0.26, 95% CI -0.46 -0.05) were associated with lower Covid-19 mortality rates. Only controlling for strictness international travel controls, and only controlling for timing later restrictions on gatherings, were also associated with lower Covid-19 mortality. Other interventions, such as stay-at-home orders or restrictions on public transport, were not significantly associated with differences in mortality rates across countries. Findings were robust across multiple statistical approaches.

**Conclusions:** Focusing on ‘compulsory’, particularly school closing, not ‘voluntary’ reduction of social interactions with mandated policies appears to have been the most effective strategy to mitigate early Covid-19 mortality.

## Background

Non-pharmaceutical interventions have generally played a critical role in reducing the transmission of Covid-19 [1, 2]. Countries around the world have introduced, at different times, to varying strictness levels, and in different combinations, a range of interventions including: public information campaigns, school and workplace closures, public event bans and restrictions on gatherings, public transport shutdowns, restrictions on internal movement (within countries), international travel controls, and stay-at-home (‘lockdown’) requirements [3]. All interventions aim to reduce virus transmission, and related morbidity and mortality, by reducing social contacts. Each policy has an obvious mechanism to achieve these social contact changes, but social behaviours can also change voluntarily without mandated policies. Their implementation can also negatively affect the economy and other health outcomes, including mental health and chronic conditions [4-7]. Assessing the relative effectiveness of concurrent interventions is essential to inform the design, including timeliness and strictness, of high-impact mitigation strategies to enable policy selection for maximal benefit with minimal disruption.

Public attention on effectiveness of Covid-19 measures has largely focused on different countries’ strategies, with certain countries singled out as exemplars based on unadjusted descriptive analysis [8]. However, early decisions on the timing and strictness of measures were driven by political and international factors (such as supply of tests and personal protective equipment), and informed by predictive epidemiological modelling. For example, a prominent report estimated over half a million deaths in the UK and two million deaths in the US before end of October 2020 without interventions, not accounting for any effects of health systems being potentially overwhelmed [9]. Particularly in early parts of the epidemic, the estimates from these predictive models, with a lack of observed impact at anything like the scale of Covid-19 interventions, were based on assumptions of the reproduction rate of the virus and ability of each intervention to alter this rate. Many subsequent studies have since used updated knowledge and prospective simulations to assess the potential impact of interventions [10-12].

However, evidence is still needed empirically on the effectiveness of non-pharmaceutical interventions on Covid-19 mortality, particularly comparing specific intervention effects. A number of studies have examined effects of interventions observed in previous epidemics [13, 14], effects of single interventions for Covid-19, predominantly stay-at-home requirements, without controlling for concurrent interventions which also affect the same outcomes [15-17], or efforts in single countries as a bundle of interventions without looking at relative effects of each [18-20]. More recent studies have reported observational evidence on the infection rate from multiple countries of bundles of interventions introduced in tandem, or in quick succession [1, 2], but the methods used likewise do not allow relative effects to be estimated.

We analysed observational data from 130 countries and exploited cross-country variation in implementation to assess the relative association between strictness and timing of individual non-pharmaceutical interventions and Covid-19 mortality, adjusting for effects of concurrent interventions. We allowed for a time lag between policy implementation and mortality to mitigate reverse causality bias and used multivariable regression models. Results inform the understanding of the drivers of differences in mortality across countries and policy choice for future waves.

## Methods

### Data

Our outcome of interest was Covid-19 attributed mortality rate. We obtained daily Covid-19 deaths for 130 countries from the European Centre for Disease Prevention and Control (ECDC) website (https://www.ecdc.europa.eu/en/covid-19/data-collection) on 1^st^ June 2020. Covid-19 mortality was measured as country daily deaths per one million population, with World Bank country population estimates from 2018 used as the denominator. Days in each country were indexed in time by first reported Covid-19 death to proxy for virus transmission rate and to make rates comparable over time.

Our variables of interest measured countries’ implementation, timing, and strictness of non-pharmaceutical interventions. We obtained data on daily national policy stringency from the Oxford COVID-19 Government Response Tracker (OxCGRT) on 1^st^ June 2020 [3]. Data are collected by over one hundred members of the OxCGRT team from publicly available sources including news reports and government briefings, using a standardised template. We examined nine non-pharmaceutical interventions which were categorised according to their strictness (see Table 1).

**Table 1:** Policy stringency coding (adapted from Hale et al. 2020)[3].

| Indicator | Coding |
| --- | --- |
| School closing | 0 - No measures<br>1 - Recommend closing<br>2 - Require closing (only some levels or categories, eg just high school, or just public schools)<br>3 - Require closing all levels |
| Workplace closing | 0 - No measures<br>1 - Recommend closing (or work from home)<br>2 - Require closing (or work from home) for some sectors or categories of workers<br>3 - Require closing (or work from home) all-but-essential workplaces (e.g. grocery stores, doctors) |
| Cancelled public events | 0 - No measures<br>1 - Recommend cancelling<br>2 - Require cancelling |
| Restrictions on gatherings | 0 - No restrictions<br>1 - Restrictions on very large gatherings (the limit is above 1000 people)<br>2 - Restrictions on gatherings between 100-1000 people<br>3 - Restrictions on gatherings between 10-100 people<br>4 - Restrictions on gatherings of less than 10 people |
| Closing public transport | 0 - No measures<br>1 - Recommend closing (or significantly reduce volume/route/means of transport available)<br>2 - Require closing (or prohibit most citizens from using it) |
| Stay at home requirements | 0 - No measures<br>1 - Recommend not leaving house<br>2 - Require not leaving house with exceptions for daily exercise, grocery shopping, and 'essential' trips<br>3 - Require not leaving house with minimal exceptions (e.g. allowed to leave only once every few days, or only one person can leave at a time, etc.) |
| Restrictions on internal movement | 0 - No measures<br>1 - Recommend closing (or significantly reduce volume/route/means of transport)<br>2 - Require closing (or prohibit most people from using it) |
| International travel controls | 0 - No measures<br>1 - Screening<br>2 - Quarantine arrivals from high-risk regions<br>3 - Ban on high-risk regions<br>4 - Total border closure |
| Public information campaigns | 0 - No COVID-19 public information campaign<br>1 - Public officials urging caution about COVID-19<br>2 - Coordinated public information campaign (e.g. across traditional and social media) |

We also collated country-level co-variates (potential confounders), which might affect the outcome and the uptake of non-pharmaceutical interventions, as detailed in Table 2. We used the most up-to-date available period comparable across countries (2018).

**Table 2:** Co-variates data.

| Measure | Reason for inclusion | Source |
| --- | --- | --- |
| Population density (people per sq. km) | As an infectious disease, higher density of population is likely to aid spread | https://data.worldbank.org/indicator/en.pop.dnst |
| % Population aged 65+ | Older persons more vulnerable to adverse effects of infection | https://data.worldbank.org/indicator/sp.pop.65up.to.zs |
| % Population male | Adverse effect of infection might vary by sex | https://data.worldbank.org/indicator/SP.POP.TOTL.MA.ZS |
| Life expectancy at birth (years) | To adjust for relative baseline health prior to pandemic | https://data.worldbank.org/indicator/SP.DYN.LE00.IN |
| Hospital beds (per 1000 people) | To adjust for relative hospital capacity prior to pandemic | https://data.worldbank.org/indicator/sh.med.beds.zs |
| Physicians (per 1000 people) | To adjust for relative workforce capacity prior to pandemic | https://data.worldbank.org/indicator/SH.MED.PHYS.ZS |
| GDP PPP (current international \$) | Gross Domestic Product (at Purchasing Power Parity), comparable measure of country wealth and relative average living standard. To adjust for relative deprivation across countries | https://data.worldbank.org/indicator/NY.GDP.MKTP.PP.CD |
| Manufacturing, value added (%GDP) | Healthcare treatment responses to the pandemic required scale-up of various equipment (e.g. ventilators, testing equipment, and personal protective equipment), globally. Extent of manufacturing base might conceivably have changed how a country was able to respond to changes in demand internally | https://data.worldbank.org/indicator/nv.ind.manf.zs |
| Health expenditure (%GDP) | To adjust for relative importance given to health budgets prior to pandemic | https://data.worldbank.org/indicator/SH.XPD.CHEX.GD.ZS |
| International tourism, number of arrivals | The virus originated in Wuhan, China. Infectious disease spread from an external source (for all other countries) will conceivably vary by extent of international movement | https://data.worldbank.org/indicator/ST.INT.ARVL |
| Governance (Voice and Accountability) | Different governance structures might impact when and how policies were introduced, and how strictly they were adhered to. We use a measure which captures the extent to which a country's citizens are able to select their government, freedom of expression, association and media, i.e. extent of democracy | https://info.worldbank.org/governance/wgi/Home/Documents#wgiDataCrossCtry |
| Region | The virus originated in the East Asia & Pacific region (Wuhan, China), so region might affect relative timing of virus arrival and any associated technological/virus evolution changes over time | https://data.worldbank.org/indicator/en.pop.dnst |
| Testing policy (h2) | With Covid-19 testing policy closely tied to attribution of Covid-19 deaths, testing policies will be inextricably linked to the outcome, i.e. more testing will offer more opportunity to attribute a death to Covid-19. We control for this difference by controlling for the extent of testing policy in a given country at a given time<br>Coding: 0 - No testing policy; 1 - Only those who both (a) have symptoms AND (b) meet specific criteria (e.g. key workers, admitted to hospital, came into contact with a known case, returned from overseas); 2 - Testing of anyone showing COVID-19 symptoms; 3 - Open public testing (eg "drive through" testing available to asymptomatic people) | https://www.bsg.ox.ac.uk/research/research-projects/coronavirus-government-response-tracker |
| Contact tracing (h3) | As above, contact tracing is closely linked and reliant on testing policy/capacity. We additionally control for the extent of contact tracing policy in a given country at a given time<br>Coding: 0 - No contact tracing; 1 - Limited contact tracing - not done for all cases; 2 - Comprehensive contact tracing – done for all cases. | https://www.bsg.ox.ac.uk/research/research-projects/coronavirus-government-response-tracker |

### Analysis

We first highlighted descriptively variations in the implementation of non-pharmaceutical interventions across selected countries. We plotted both the stringency and timing of intervention implementation against daily number of deaths per 1,000,000 population over indexed time. We smoothed the series by plotting estimates from local linear regressions [21]. We also examined the unadjusted associations between interventions and Covid-19 mortality with scatterplots.

We then used the known time lag between the virus transmission (which is affected by non-pharmaceutical interventions) and mortality to address potential reverse causality caused by policymakers introducing non-pharmaceutical interventions in response to a rising mortality rate, which would downwards bias the effect of introducing the interventions. Evidence suggests a mean lag between virus transmission and symptom onset of 6 days [22, 23], and a further mean lag of 18 days between onset of symptoms and death [24], resulting in an overall likely mean lag of 24 days between intervention introduction and associated Covid-19 mortality. This duration underpins a choice of 24-day periods for modelling impacts and the decision to measure implementation timing and strictness prior to the period of mortality analysis.

We considered two distinct time periods of 24 days to account for the changing magnitude of effects of an exponentially transmitted virus: i) 0-24 days and ii) 14-38 after the first Covid-19 indexed death. The analysis at 0-24 days is very unlikely to suffer from bias caused by reverse causality since policymakers cannot possibly be reacting to deaths within their own country. The 14-38 days analysis, on the other hand, has a small risk of this bias if policymakers react quickly to deaths within the first 14 days. However, the mean number of cumulative deaths in the analysis sample at 14 days was 31 (SD 68) suggesting the mortality rate was still relatively low in most countries.

For each of the two 24-day periods we calculated the strictness and timing of each of the nine non-pharmaceutical interventions prior to the start of the period (i.e. policies implemented before the day of the first reported death for the 0-24 day period, and up to 14 days after the first reported death for the 14-38 day period).

#### Primary analysis

For our primary analysis we generated an aggregated measure of strictness and timeliness to account for the changes in interventions over time. Mean intervention strictness was calculated for each of the 24-day time periods: i) from 30 days before (to ensure comparable time periods over which the average was taken) the first Covid-19 death; and ii) from 30 days before until 14 days after the first Covid-19 death. Countries with earlier and stricter non-pharmaceutical interventions had higher mean strictness values.

#### Secondary analysis

Due to the dynamic nature of policy implementation, it is inadequate to observe only policy strictness or timing alone. However, doing so assisted in interpretation of the analysis of the combined mean score. For secondary analysis we took the maximum strictness value (see Table 1) for each policy in each country over the analysis period. To account for differences in the start date of virus transmission in each country and ensure country comparability, we constructed a variable which counted the days between the implementation of an intervention and the first recorded Covid-19 death (e.g. -10 if implemented 10 days before the first recorded Covid-19 death). Intervention timing was modelled categorically to avoid imposing linear effects.

For each analysis we ran multivariable regression models of the lagged nine concurrent non-pharmaceutical intervention status on daily Covid-19 death per million population over the subsequent pooled 24-day cross-section. We also included a range of covariates (Table 2), a set of categorical indicators for day-of-the-week and a set of categorical indicators for week-of-the-year to capture seasonality, and a time fixed effect (number of days since first death in country) to account for the magnitude of effects of death varying over the 24-day analysis period due to exponential virus spread. Standard errors were clustered at the country-level. Further details on the empirical strategy are provided in the Additional file (section 1).

We excluded countries with less than a population of 100,000 and we analysed data for countries (n=130) with no missing policy indicators, outcome, or covariates at each time point.

### Sensitivity analysis

Firstly, there may be bias from varying definitions of a Covid-19 attributed death and recording practices across countries. The European Observatory categorises deaths as either ‘clinical-diagnosis-based’ or ‘test-based’ [25]. Country’s choice of definition may over/under-inflate reported Covid-19 deaths and may be associated with the timing/stringency of non-pharmaceutical interventions. To examine this, for the 28 countries with available data on death definition, we used logistic regressions to examine the association between each mean non-pharmaceutical intervention implementation and the Covid-19 death definition. We also controlled for a country’s testing and contact tracing policies in all regression models (Table 2), more stringent testing/tracing increases a country’s ability to attribute deaths to Covid-19.

Secondly, accuracy of Covid-19 deaths may also vary due to challenges in determining underlying cause of death. For example, there have been questions over the accuracy of mortality reporting in China [26], and there is a high per-capita Covid-19 mortality rate in Belgium due to the inclusion of all deaths in care homes regardless of cause [27]. We therefore conducted a sensitivity analysis omitting these two countries from analyses.

Thirdly, as daily deaths per million is an over-dispersed count variable, we repeated all analyses using negative binomial regression (reporting incidence rate ratios) to check the consistency of the direction and significance of policy effect across non-linear models.

## Results

Figure 1 shows the introduction of non-pharmaceutical interventions to address Covid-19 transmission and daily Covid-19 deaths for selected countries. Notably, elucidating mortality impacts from separate interventions using visual aids or statistically without controlling for those co-introduced is problematic given the introduction of multiple interventions.

**Figure 1:**
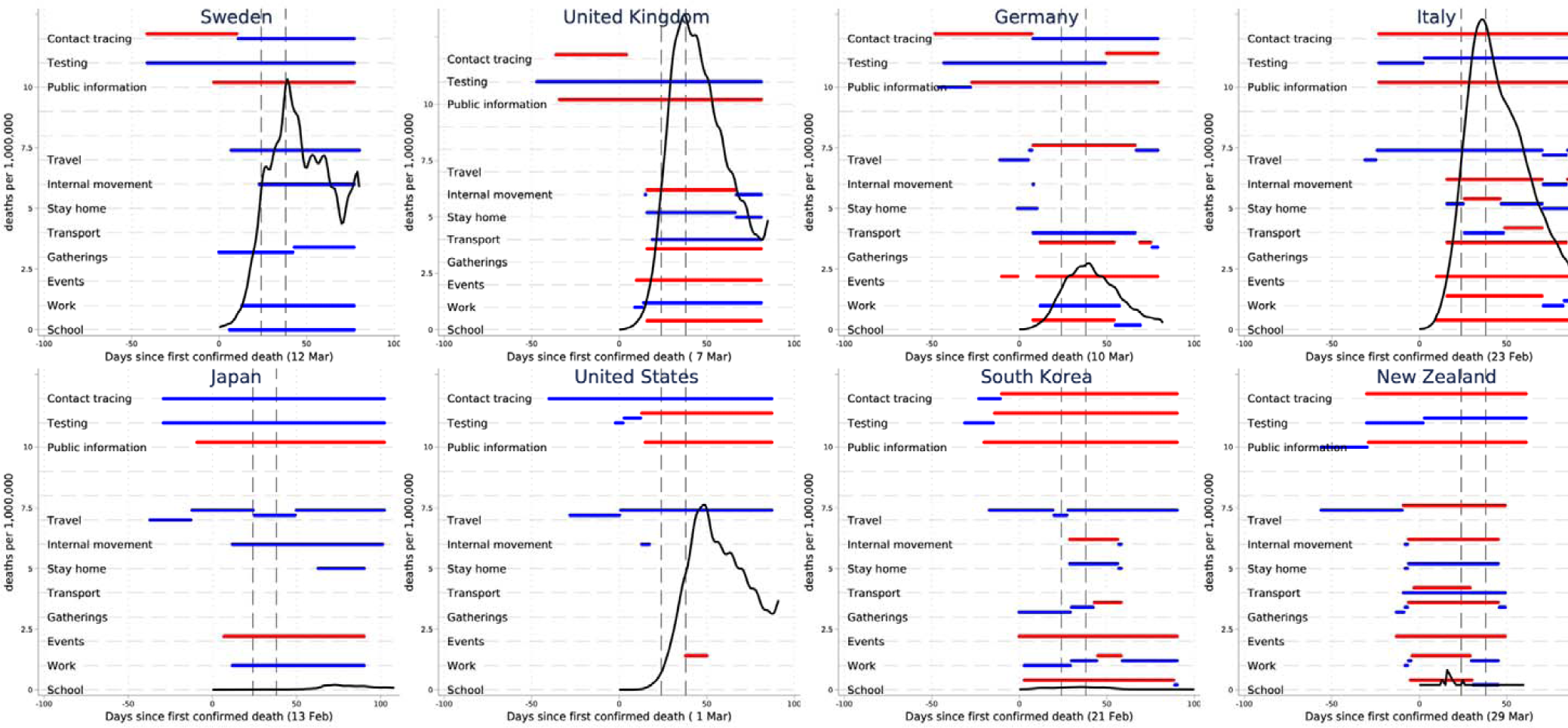
Implementation of non-pharmaceutical interventions and daily Covid-19 death rates for 8 selected countries. Notes: Dotted horizontal line indicates strictness of implementation, with maximum implementation in red, any other implementation in blue. Locally weighted regressions (bandwidth = 0.2) of the raw daily deaths per million on time. Dashed vertical lines identify the periods of analysis, 24- and 38-days after first confirmed Covid-19 death. D of first confirmed death observed in parenthesis.

In unadjusted analysis (illustrated in the scatter graphs in Additional file section 2), earlier and stricter intervention implementation of all nine non-pharmaceutical interventions were associated with lower Covid-19 deaths, as expected, although effects were small in magnitude. Table 3 shows the variation in intervention exposure across countries in each analysis period.

**Table 3:**
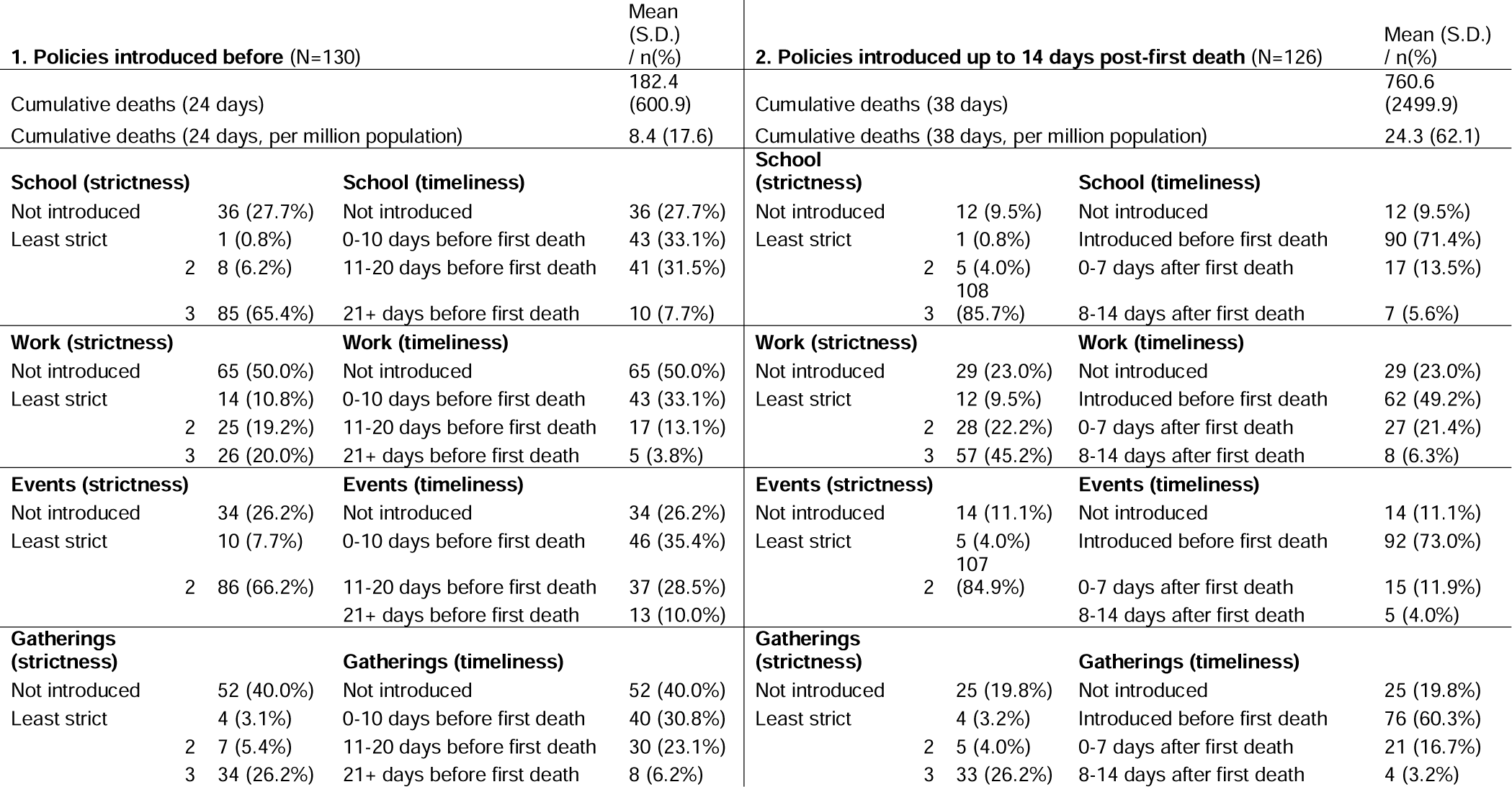
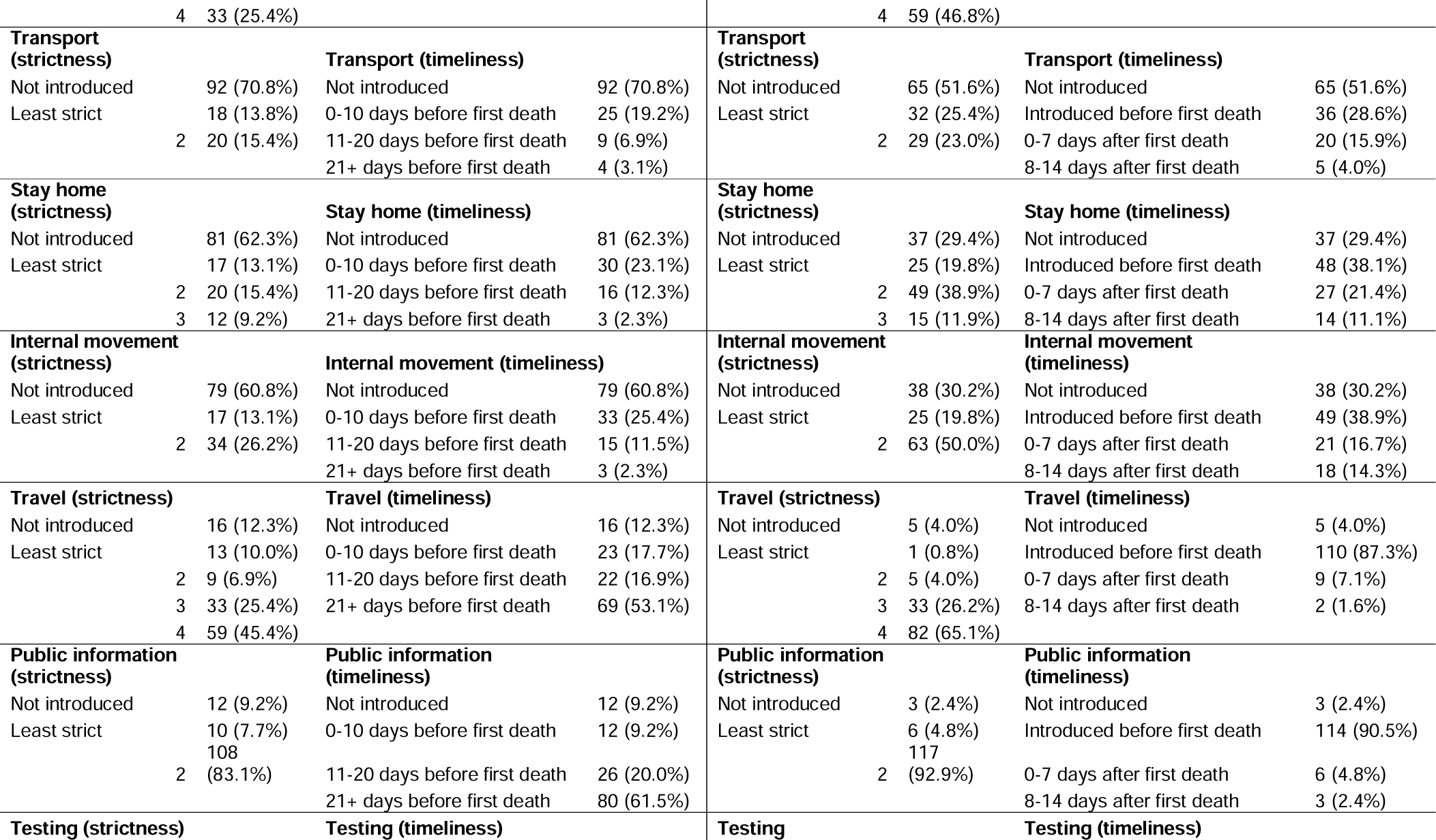
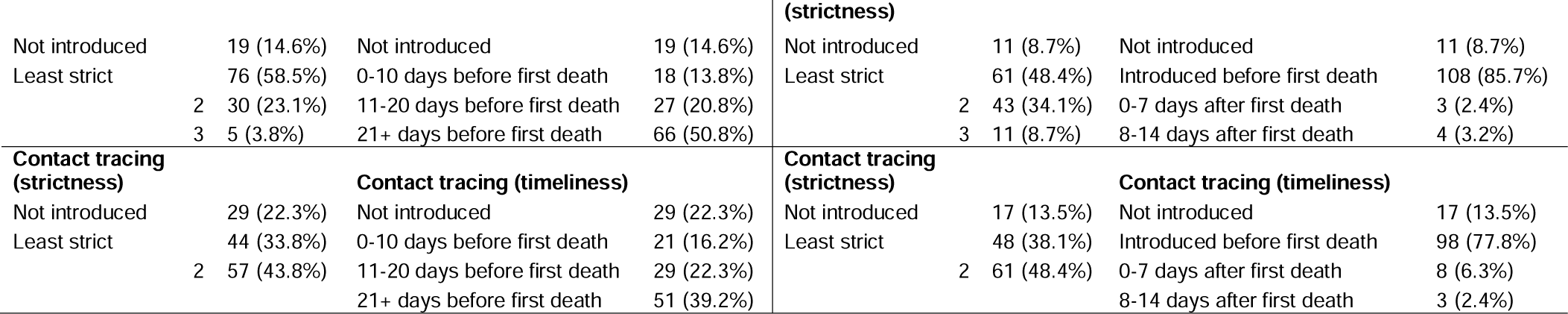
Country variation in timing/strictness of policies at each analysis period.

Mean strictness/timing of non-pharmaceutical interventions were not associated with Covid-19 death definitions suggesting death definition is unlikely to bias the estimated coefficients (see Additional file section 3).

### Primary analysis

Figure 2 shows the results from the pooled cross-sectional regression analysis including both policy strictness and timing (through the mean score). Stricter/earlier workplace closures were associated with fewer Covid-19 deaths in the early part of the epidemic (first 24 days), -0.26 per million (95% CI -0.46 -0.05) with a one unit increase in the mean score (equivalent to, for example, a strictness score of 1 over the entire time period, 2 for half the time period, 3 for a third of the time period, or 4 for a quarter of the time period). In the 14-38 days analysis, where the magnitudes are expected to be larger due to exponential virus spread, stricter/earlier school closures were associated with the largest reductions in Covid-19 deaths of -1.23 per million (95% CI -2.20 -0.27). Mean scores of other interventions were not associated with Covid-19 mortality.

**Figure 2:**
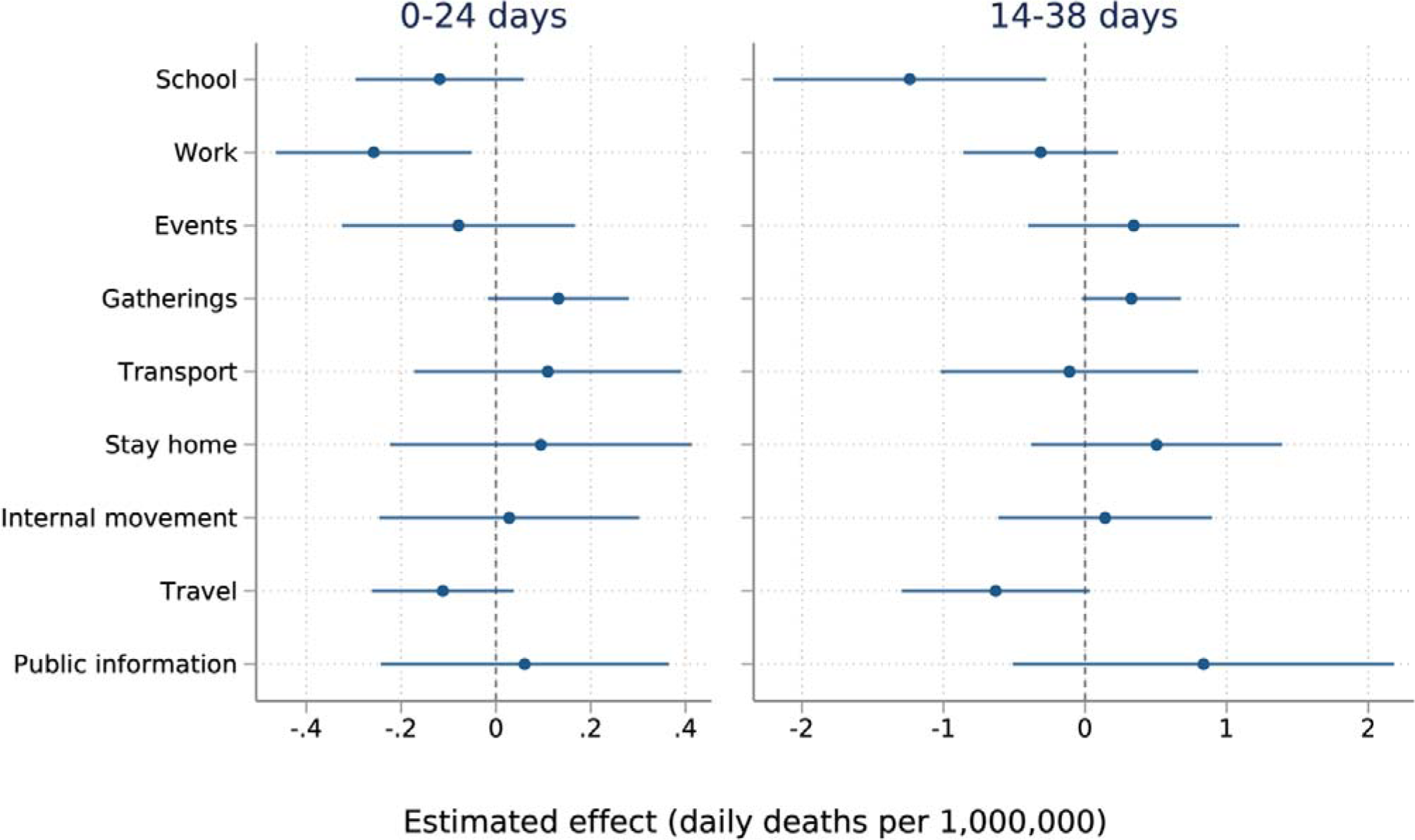
Regression results examining policy strictness and timing combined (mean score) *Notes:* Estimated parameters of two regressions adjusted for a range of covariates (Table 2), a set of categorical indicators for day-of-the-week and a set of categorical indicators for week-of-the-year to capture seasonality, and the time (number of days since first death in country) to account for the magnitude of effects of death varying over the 24-day analysis period due to exponential virus spread. Standard errors were clustered at the country-level. *Sample size:* 130 countries (3250 observations) for 0-24 days analysis; 126 countries (3150 observations) for 14-38 days analysis.

Early/strict school closing was consistently associated with lower mortality rates across all sensitivity checks in the 14-38 days analysis and had a negative but not statistically significant estimate in all 0-24 days analyses (see Additional file section 4). Stricter/earlier workplace closures was robust to the analysis without China/Belgium at 0-24 days. The result was negative but not significant in the negative binomial model. The estimate was also statistically significant in the 14-38 days analysis without China/Belgium. Earlier/stricter restrictions on gatherings was not significant in the primary analysis but was significantly associated with a slightly larger mortality rate in both robustness checks at 0-24 days and only in the analysis without China/Belgium at 14-38 days.

### Secondary analysis

Results from the fully-adjusted pooled cross-sectional regressions examining intervention strictness alone (Figure 3) and timing alone (Figure 4) show that implementation of school closing prior to first recorded death appears to be driving the effect of school closing on mortality rates, with required rather than recommended closing (least strict) also potentially preferable. The driver of the workplace finding was less obvious, but again the earlier implementation appeared to be the main driver of the result.

**Figure 3:**
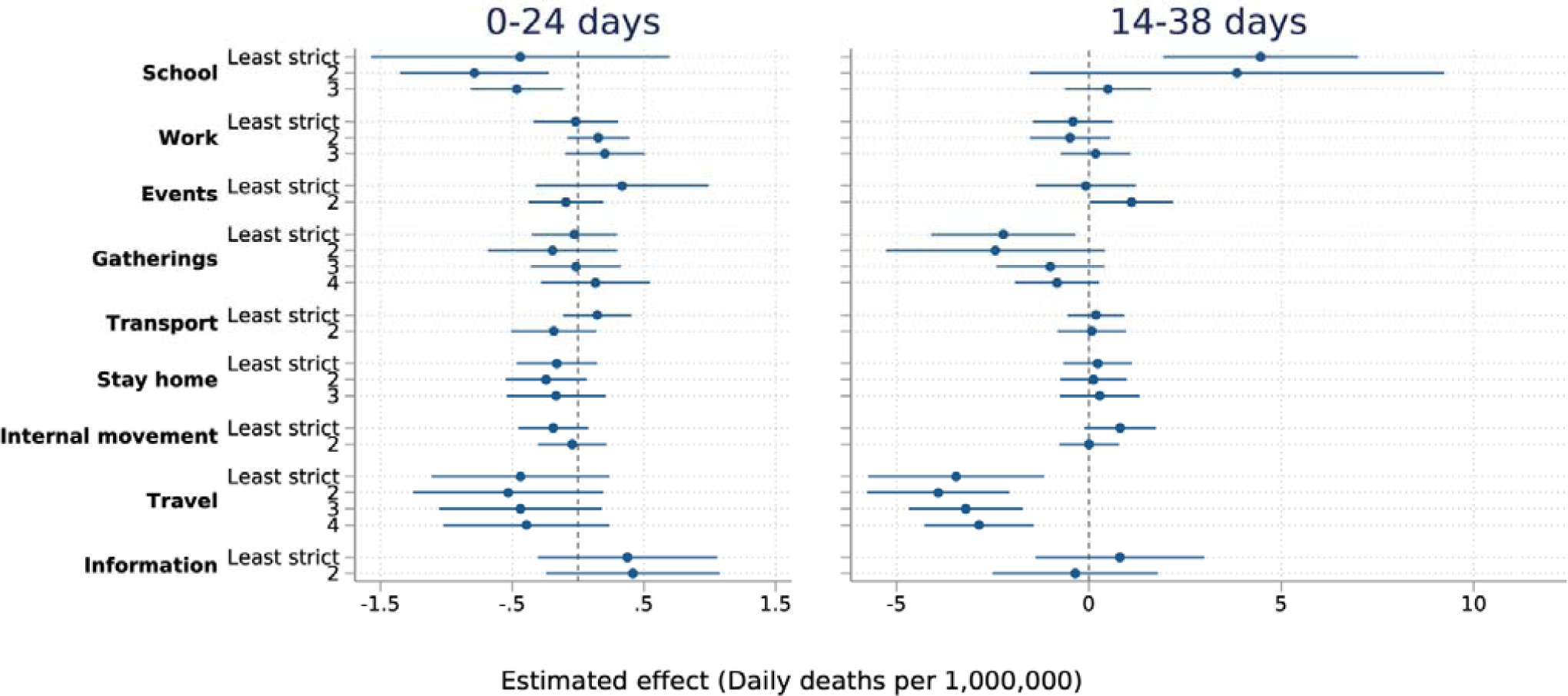
Regression results examining policy strictness alone. *Notes:* Estimated parameters of two regressions adjusted for a range of covariates (Table 2), a set of categorical indicators for day-of-the-week and a set of categorical indicators for week-of-the-year to capture seasonality, and the time (number of days since first death in country) to account for the magnitude of effects of death varying over the 24-day analysis period due to exponential virus spread. Standard errors were clustered at the country-level. Baseline is ‘not introduced during intervention analysis period’ for each policy. *Sample size:* 130 countries (3250 observations) for 0-24 days analysis; 126 countries (3150 observations) for 14-38 days analysis.

**Figure 4:**
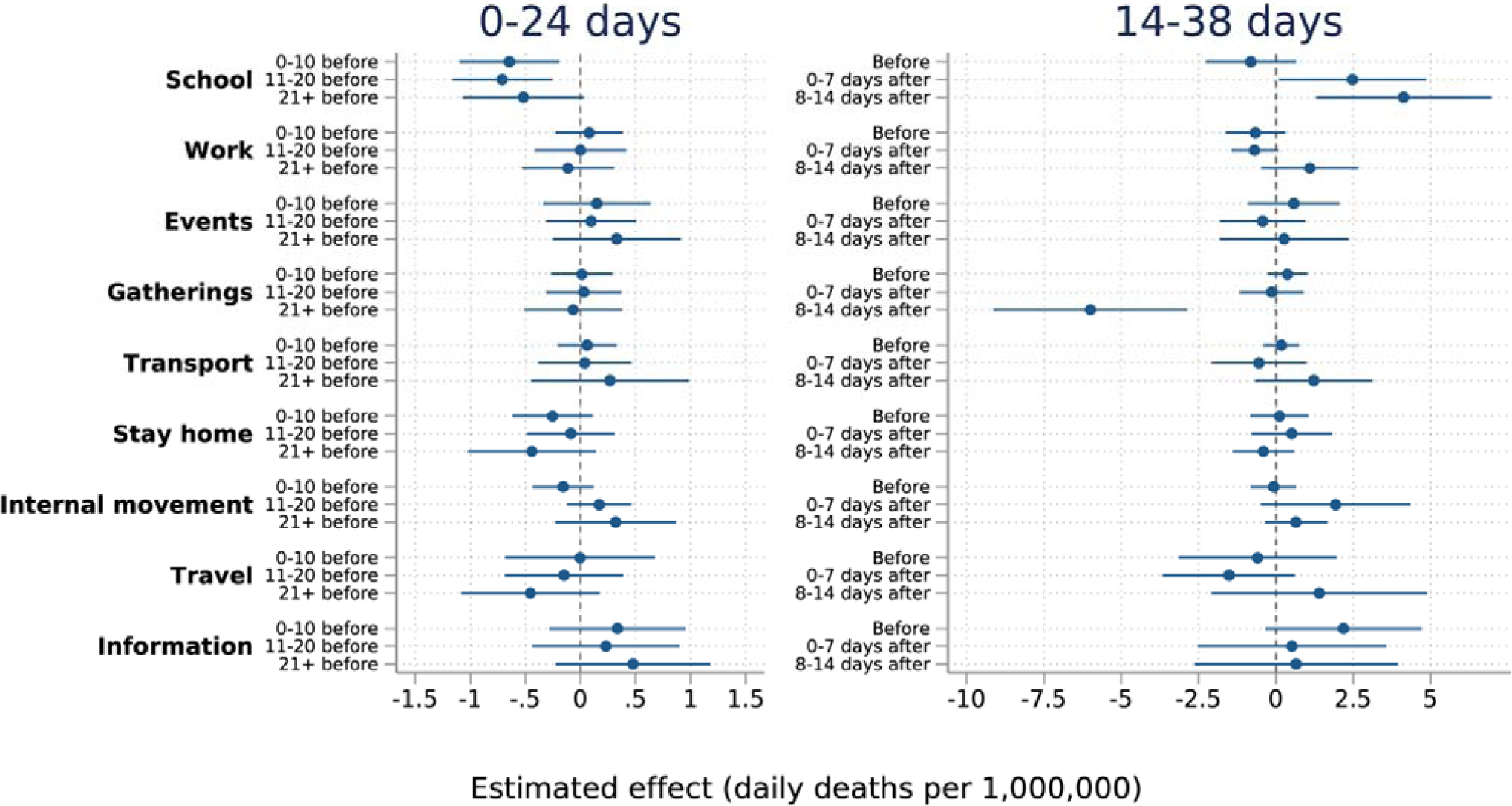
Regression results examining policy timing alone. Notes: Estimated parameters of two regressions adjusted for a range of covariates (Table 2), a set of categorical indicators for day-of-the-week and a set of categorical indicators for week-of-the-year to capture seasonality, and the time (number of days since first death in country) to account for the magnitude of effects of death varying over the 24-day analysis period due to exponential virus spread. Standard errors were clustered at the country-level. Baseline is ‘not introduced during intervention analysis period’ for each policy. *Sample size:* 130 countries (3250 observations) for 0-24 days analysis; 126 countries (3150 observations) for 14-38 days analysis.

Results from additional sensitivity analyses (see Additional file section 4) included stricter international travel controls consistently associated with lower Covid-19 mortality rates in the 14-38 days analyses. Strictness of restrictions on gatherings was associated with slightly lower Covid-19 mortality rates in the 14-38 days analyses. Later restrictions on gatherings were also associated with lower Covid-19 mortality across all 14-38 analyses. Later implementers tended to introduce the strictest version (restrictions on gatherings of less than 10 people) compared to earlier implementers of this policy (75% implementing the strictest version 8-14 days after first death, versus 50% at 0-7 days and 60% of those that introduced before first death).

## Discussion

Using data from 130 countries, we examined the effects of non-pharmaceutical interventions on Covid-19 attributed mortality. We used information on policy implementation prior to the period over which we analyse deaths to mitigate the bias from reverse causality. We found that earlier and stricter implementation of some non-pharmaceutical interventions contributed to relatively lower numbers of Covid-19 deaths. Notably, the earlier/stricter implementation of school and workplace closures were associated with lower Covid-19 mortality rates. Without controlling for policy timing stricter international travel controls, and without controlling for policy strictness later restrictions on gatherings, were also associated with lower Covid-19 mortality across all 14-38 day analyses.

There are key limitations of this analysis to consider when interpreting the findings. First, although we control for a range of potential confounders, there is a risk of unobserved time-varying confounding. However, other methods typically used to examine causal effects of interventions, such as difference-in-differences, are biased in settings with multiple policies implemented across time and geography, because pre-intervention trends for one intervention are impacted by any effects of pre-existing policies [28]. Instead, we use the known lag period between intervention effect and our outcome of interest to control for reverse causality as much as possible.

Secondly, this study only examines the impact of nationally recorded policies, meaning subnational interventions were not captured. Furthermore, we were unable to measure compliance and regional variation in implementation, as well as voluntary changes in population behaviours. We were also unable to estimate longer-term effects due to limited statistical power and increased risk of bias due to reverse causality in later periods. A key limitation is the nature of these interventions. Whilst we examine them independently and look for separate effects, there are likely to be interaction effects. Additionally, we examine a narrow window of time in the initial stages of the outbreak. Alternative analysis methods will likely be necessary to reduce bias of reverse causality in these later periods, however.

Thirdly, we are only able to examine a single, albeit important, outcome, mortality, due to comparable data availability and the necessity of a known lag period between intervention and outcome. The relative effectiveness of non-pharmaceutical interventions against other outcomes, such as mental health and economic outcomes, and potential trade-offs across outcomes, are also important. As noted above, there is also variation in definitions and accuracy of mortality reporting across countries [25], however robustness checks and adjusting for testing and contact tracing policies suggest this does not greatly impact the findings. Excess mortality, mortality compared to previous years, is arguably a better measure to also take account of unattributed and indirect deaths. Unfortunately, excess mortality relies on calculations (compared to a pre-period) and is not comparatively available for as many countries, necessary for statistical power to incorporate concurrent interventions. However, a recent study also showed that the mortality data we use is able to correctly signal the true trajectory of the trend when compared to excess mortality data available in 17 countries [29]. Mortality also lies on a complex pathway from initial virus transmission that involves population health and behaviours, underlying health inequalities, and healthcare access and quality. Further understanding the patterns and causal mechanisms which have affected mortality rates is essential for future outbreaks but is not possible with current data.

The findings from this study align with evidence from previous pandemics that also concluded that interventions such as workplace closures, school closures, and quarantine periods were most effective for reducing cases [13]. There was previously much less evidence available on the effects of internal and international travel restrictions, bans on mass gatherings and public information campaigns [13]. A rapid systematic review early in the Covid-19 pandemic examining the effects of school closures reported a dearth of evidence on the topic, but highlighted that modelling studies assumed a 2-4% reduction in deaths, much less than attributed to other non-pharmaceutical interventions [14].

Recently published Covid-19 specific studies on the impacts of single policies have mostly focused on stay-at-home interventions. For example, a Californian study estimated 1661 fewer deaths over one month from the introduction of a stay-at-home policy (at an estimated cost of 400 job losses per life saved) [15], whilst a study from US states estimated stay-at-home interventions were associated with approximately 50% fewer deaths, with early adopting states experiencing larger reductions in mortality [16]. The impact of stay-at-home interventions have been contrasted between Denmark and Sweden which suggested the number of deaths would have been 167% higher in Denmark without stay-at-home interventions [17]. These findings contrast to our study which found no association between mandatory stay-at-home interventions on cross-country Covid-19 mortality after adjusting for other non-pharmaceutical interventions concurrently introduced. One study examining the effects of multiple policies in Hong Kong shows that the combined package of policies introduced, including border restrictions, quarantine, and social distancing, were associated with reduced transmission of Covid-19 [18]. A recent study also showed that school closures, particularly implemented early, have been effective at reducing incidence (−62%) and mortality (−58%) in the US [20].

Other cross-country comparisons of non-pharmaceutical interventions have identified that travel restrictions were associated with slower geographical spread and initial case numbers, but failed to quantify the specific effect [19]. One study from six countries (China, South Korea, Italy, Iran, France, and the US) predicted that combined non-pharmaceutical interventions might have prevented or delayed a total of 62 million cases [2]. Our findings are also consistent with a study examining interventions implemented in 30 European countries which reported school closures, non-essential businesses closures, and prohibiting mass gatherings to be effective in reducing Covid-19 deaths [30]. The most recent cross-country comparison examined effects of multiple interventions simultaneously on disease incidence using meta-analysis. The authors reported a general overall finding that “physical distancing interventions were associated with reductions in the incidence of covid-19 globally” [1], but were not able to look at relative effects. Nevertheless, detailed examination of their findings shows that the largest effect on reducing incidence was a combination of school closures, workplace closures and restrictions on mass gatherings, directly in line with our findings.

### Implications for policy and practice

The finding that selected non-pharmaceutical interventions are associated with lower Covid-19 mortality is not surprising given the known reductions in social contacts and transmission from these interventions, and their historical use to control epidemics. However, it may be unexpected that workplace and, particularly, school closures were associated with relatively lower Covid-19 mortality across countries whilst interventions such as stay-at-home measures were not. One plausible interpretation is that schools and workplaces involve ‘compulsory’ interactions with others as individuals feel obliged to attend in person and may be concerned for loss of earning or educational opportunities. This compares to interventions targeting other sources of human interaction which are more ‘voluntary’ and may reduce irrespective of whether mandated policies are introduced (therefore giving no additional observable effect of introducing the intervention). This mechanism is supported by multiple studies examining the effects of non-pharmaceutical interventions on mobility. One US study found mobility reductions in all US states, even those not adopting formal stay-at-home measures, due to individuals’ voluntary behaviour changes [31]. Notably, mobility fell *prior* to stay-at-home implementation in US states which introduced measures [32], and mobility fell a comparable amount (52%) in US counties *without* stay-at-home measures compared to those with (61%) [33]. Likewise, examination of Google mobility data in Sweden, with a lack of stay-at-home measures, shows mobility nevertheless decreased in retail and workplaces by roughly -25% and transit -35% by end of March [34]. Unfortunately, this mobility data is not comparable across countries in order to analyse this specific mechanism within this study. We do not suggest that these other interventions are not effective, therefore, but the cumulative evidence above suggests that mandatory introduction might be unnecessary if people change their social interaction behaviours voluntarily.

School closing having the largest effect is also interesting, in that children are known to be relatively less personally affected by the negative effects of Covid-19 than older adults [35]. This result highlights the well-documented problem of asymptomatic transmission in Covid-19 [36]. The findings suggest caution as schools and universities begin to return around the world while the virus remains in circulation. The extremely low individual risk to these groups might encourage more risky behaviour which can subsequently affect wider households and communities. This relative risk behaviour can already be seen in media around the world reporting house parties/raves and u-turns on policies opening University campuses as infection rates subsequently accelerate, for instance [37, 38]. There is the need to examine specific interventions to incentivise societally-optimal behaviour and internalising of the population-level risk for these low-risk individuals. In the meantime, this might require a more mandatory approach to policy implementation.

Overall, policymakers should note that efforts to reduce social interactions through restricting compulsory activities, such as school and work, may be more effective in reducing mortality than targeting actions where individuals have more freedom in choice or where activities are likely diminished due to other actions. However, these results only apply to the early stages of the Covid-19 pandemic. For identifying responses to future waves or other novel pathogens understanding the complex behaviour changes from specific interventions is necessary for appropriate mitigation. There is no guarantee that the behaviour changes outlined above would be replicated if population complacency or a primary focus on other immediate pressures, such as financial hardship, prevail in future waves.

#### Future research

Future research should examine long-term effects of interventions, such as whether countries suppressing stricter/earlier are more vulnerable to future outbreaks. Understanding the inter-connected nature of different interventions and their impacts on population behaviours, including how these behaviours change over time, is key. The important role of different countries’ health systems, supply chains and/or cultures should also be understood as important mediators in preventing Covid-19 mortality. Understanding how the targeting of specific interventions to certain populations and protecting high-risk groups (e.g. elderly) and institutions (e.g. hospitals and care homes) will hopefully reveal more nuanced policy responses for future outbreaks, and integrating policy options into wider behavioural science, for example through nudging or encouraging select behaviours [39], might offer long-term solutions that are more acceptable to populations. Lastly, future research should quantify the trade-offs against the large economic considerations that not only have monetary costs, but profound societal and long-term health impacts [4, 5].

## Conclusions

Early workplace and, particularly, school closures were associated with the lowest Covid-19 mortality rates across 130 countries. Focusing on protecting individuals from social interactions by targeting more ‘compulsory’ places (including schools and workplaces) as opposed to more ‘voluntary’ interactions and changing behaviours of those with lower individual-risk appear to have been most effective strategies mitigating early Covid-19 mortality.

## Data Availability

The datasets analysed during the current study are available from:
-Policy data: https://covidtracker.bsg.ox.ac.uk/ -Covid-19 mortality data: https://www.ecdc.europa.eu/en/publications-data/downloadtodays-data-geographic-distribution-covid-19-cases-worldwide

https://covidtracker.bsg.ox.ac.uk/

https://www.ecdc.europa.eu/en/publications-data/downloadtodays-data-geographic-distribution-covid-19-cases-worldwide

## Availability of data and materials

The datasets analysed during the current study are available from:

– Policy data: https://covidtracker.bsg.ox.ac.uk/
– Covid-19 mortality data: https://www.ecdc.europa.eu/en/publications-data/downloadtodays-data-geographic-distribution-covid-19-cases-worldwide

## Authors’ contributions

All authors helped conceptualise the study. JS analysed the data and wrote the first draft. All authors contributed to the interpretation of the analysis and contributed to the final draft.

## References

1. Islam N, Sharp SJ, Chowell G, Shabnam S, Kawachi I, Lacey B, Massaro JM, D’Agostino RB, White M: Physical distancing interventions and incidence of coronavirus disease 2019: natural experiment in 149 countries. BMJ 2020,370:m2743.

2. Hsiang S, Allen D, Annan-Phan S, Bell K, Bolliger I, Chong T, Druckenmiller H, Huang LY, Hultgren A, Krasovich E: The effect of large-scale anti-contagion policies on the COVID-19 pandemic. Nature 2020:1–9.

3. Hale T, Webster S, Petherick A, Phillips T, Kira B: Oxford covid-19 government response tracker. Blavatnik School of Government 2020, 25.

4. Adams-Prassl A, Boneva T, Golin M, Rauh C: The Impact of the Coronavirus Lockdown on Mental Health: Evidence from the US. In.; 2020.

5. Fernandes N: Economic effects of coronavirus outbreak (COVID-19) on the world economy. Available at SSRN 3557504 2020.

6. Chetty R, Friedman JN, Hendren N, Stepner M: Real-Time Economics: A New Platform to Track the Impacts of COVID-19 on People, Businesses, and Communities Using Private Sector Data. In.: Mimeo; 2020.

7. Maringe C, Spicer J, Morris M, Purushotham A, Nolte E, Sullivan R, Rachet B, Aggarwal A: The impact of the COVID-19 pandemic on cancer deaths due to delays in diagnosis in England, UK: a national, population-based, modelling study. The Lancet Oncology 2020, 21(8):1023–1034.

8. Gibney E: Whose coronavirus strategy worked best? Scientists hunt most effective policies. Nature 2020.

9. Ferguson N, Laydon D, Nedjati Gilani G, Imai N, Ainslie K, Baguelin M, Bhatia S, Boonyasiri A, Cucunuba Perez Z, Cuomo-Dannenburg G: Report 9: Impact of non-pharmaceutical interventions (NPIs) to reduce COVID19 mortality and healthcare demand. 2020.

10. Flaxman S, Mishra S, Gandy A, Unwin HJT, Mellan TA, Coupland H, Whittaker C, Zhu H, Berah T, Eaton JW: Estimating the effects of non-pharmaceutical interventions on COVID-19 in Europe. Nature 2020:1–8.

11. Walker PG, Whittaker C, Watson OJ, Baguelin M, Winskill P, Hamlet A, Djafaara BA, Cucunubá Z, Mesa DO, Green W: The impact of COVID-19 and strategies for mitigation and suppression in low-and middle-income countries. Science 2020.

12. Davies NG, Kucharski AJ, Eggo RM, Gimma A, Edmunds WJ, Jombart T, O’Reilly K, Endo A, Hellewell J, Nightingale ES: Effects of non-pharmaceutical interventions on COVID-19 cases, deaths, and demand for hospital services in the UK: a modelling study. The Lancet Public Health 2020.

13. Cheatley J, Vuik S, Devaux M, Scarpetta S, Pearson M, Colombo F, Cecchini M: The effectiveness of non-pharmaceutical interventions in containing epidemics: a rapid review of the literature and quantitative assessment. medRxiv 2020.

14. Viner RM, Russell SJ, Croker H, Packer J, Ward J, Stansfield C, Mytton O, Bonell C, Booy R: School closure and management practices during coronavirus outbreaks including COVID-19: a rapid systematic review. The Lancet Child & Adolescent Health 2020, 4(5):397–404.

15. Friedson AI, McNichols D, Sabia JJ, Dave D: Did california’s shelter-in-place order work? early coronavirus-related public health effects. In.: National Bureau of Economic Research; 2020.

16. Dave DM, Friedson AI, Matsuzawa K, Sabia JJ: When do shelter-in-place orders fight COVID-19 best? Policy heterogeneity across states and adoption time. In.: National Bureau of Economic Research; 2020.

17. Juranek S, Zoutman F: The effect of social distancing measures on the demand for intensive care: Evidence on covid-19 in scandinavia. 2020.

18. Cowling BJ, Ali ST, Ng TWY, Tsang TK, Li JCM, Fong MW, Liao Q, Kwan MYW, Lee SL, Chiu SS et al: Impact assessment of non-pharmaceutical interventions against coronavirus disease 2019 and influenza in Hong Kong: an observational study. The Lancet Public Health 2020, 5(5):e279–e288.

19. Imai N, Gaythorpe KA, Abbott S, Bhatia S, van Elsland S, Prem K, Liu Y, Ferguson NM: Adoption and impact of non-pharmaceutical interventions for COVID-19. Wellcome Open Research 2020, 5(59):59.

20. Auger KA, Shah SS, Richardson T, Hartley D, Hall M, Warniment A, Timmons K, Bosse D, Ferris SA, Brady PW et al: Association Between Statewide School Closure and COVID-19 Incidence and Mortality in the US. Jama 2020, 324(9):859–870.

21. Cleveland WS: Robust locally weighted regression and smoothing scatterplots. Journal of the American statistical association 1979, 74(368):829–836.

22. Bi Q, Wu Y, Mei S, Ye C, Zou X, Zhang Z, Liu X, Wei L, Truelove SA, Zhang T: Epidemiology and transmission of COVID-19 in 391 cases and 1286 of their close contacts in Shenzhen, China: a retrospective cohort study. The Lancet Infectious Diseases 2020.

23. Backer JA, Klinkenberg D, Wallinga J: Incubation period of 2019 novel coronavirus (2019-nCoV) infections among travellers from Wuhan, China, 20–28 January 2020. Eurosurveillance 2020, 25(5):2000062.

24. Verity R, Okell LC, Dorigatti I, Winskill P, Whittaker C, Imai N, Cuomo-Dannenburg G, Thompson H, Walker PG, Fu H: Estimates of the severity of coronavirus disease 2019: a model-based analysis. The Lancet infectious diseases 2020.

25. How comparable is COVID-19 mortality across countries? [https://analysis.covid19healthsystem.org/index.php/2020/06/04/how-comparable-is-covid-19-mortality-across-countries/]

26. Trump questions accuracy of China’s coronavirus death toll [http://www.nbcnews.com/news/world/trump-questions-accuracy-china-s-coronavirus-death-toll-n1185116>]

27. Why Belgium has the highest coronavirus death rate in the world [http://www.independent.co.uk/news/world/europe/coronavirus-belgium-death-toll-lockdown-trump-who-uk-spain-italy-a9494186.html>]

28. Goodman-Bacon A, Marcus J: Using Difference-in-Differences to Identify Causal Effects of COVID-19 Policies. 2020.

29. Dergiades T, Milas C, Panagiotidis T: Effectiveness of Government Policies in Response to the COVID-19 Outbreak. Available at SSRN 3602004 2020.

30. Hunter PR, Colon-Gonzalez F, Brainard JS, Rushton S: Impact of non-pharmaceutical interventions against COVID-19 in Europe: a quasi-experimental study. medRxiv 2020.

31. Gupta S, Nguyen TD, Rojas FL, Raman S, Lee B, Bento A, Simon KI, Wing C: Tracking public and private response to the COVID-19 epidemic: Evidence from state and local government actions. In.: National Bureau of Economic Research; 2020.

32. Engle S, Stromme J, Zhou A: Staying at home: mobility effects of covid-19. Available at SSRN 2020.

33. Dasgupta N, Funk MJ, Lazard A, White BE, Marshall SW: Quantifying the social distancing privilege gap: a longitudinal study of smartphone movement. medRxiv 2020.

34. Community Mobility Reports [https://www.google.com/covid19/mobility/]

35. Götzinger F, Santiago-García B, Noguera-Julián A, Lanaspa M, Lancella L, Calò Carducci FI, Gabrovska N, Velizarova S, Prunk P, Osterman V et al: COVID-19 in children and adolescents in Europe: a multinational, multicentre cohort study. The Lancet Child & Adolescent Health 2020, 4(9):653–661.

36. Gandhi M, Yokoe DS, Havlir DV: Asymptomatic Transmission, the Achilles’ Heel of Current Strategies to Control Covid-19. New England Journal of Medicine 2020, 382(22):2158–2160.

37. ‘Everybody just wants to get out now’: The Europeans seeking thrills at illegal parties despite coronavirus [https://www.independent.co.uk/news/world/europe/coronavirus-europe-illegal-ravesparties-london-paris-berlin-a9662821.html]

38. Yamey G, Walensky RP: Covid-19: re-opening universities is high risk. BMJ 2020, 370:m3365

39. Thaler RH, Sunstein CR: Nudge: improving decisions about health. Wealth, and Happiness 2008, 6.

