## Additional file for "The relative effects of non-pharmaceutical interventions on early Covid-19 mortality: natural experiment in 130 countries"

### Appendix

#### Section 1: Pooled cross-sectional analysis specifications (details of empirical strategy)

We built the models up to the fully-adjusted model outlined in the main text. For all models, we ran a set of linear regression models using daily deaths per million as the outcome. Our estimates of interest are the $\beta$ terms, an estimate of the effect of each individual policy on the outcome. The specifications were as follows:

1) Simple unadjusted models. We ran nine separate models estimating the effect of each policy individually

$$Deaths_{it} = \alpha + \beta PStr_i/PTime_i/PMean_i + \varepsilon_{it}$$

Where:

i=country; t= day relative to the day of the first Covid-19 death
$PStr_i$, $PTime_i$, and $PMean_i$ were three sets of policy variables:

> $PStr_i$ for each policy indicated the maximum level of policy stringency (coded as in Table 1) over the policy analysis period (policy period up to first death/ policy period up to 14-days post-first death)
>
> $PTime_i$ for each policy indicated the categorical variable for introduction timing (for the policy period up to first death: 0=not introduced over period, 1=introduced 0-10 days before first death, 2=11-20 days before first death, 3=20+ days before first death; for the policy period up to 14-days post-first death: 0=not introduced over period, 1=introduced before first death, 2=introduced 0-7 days after first death, 3=8-14 days after first death)
>
> $PMean_i$ for each policy was the mean stringency score over the policy analysis period

$\varepsilon_{it}$ was the error term, clustered by country

2) Estimate controlling for the effects of all other implemented policies in a single model

$$Deaths_{it} = \alpha + \beta_{1-9} PStr_i/PTime_i/PMean_i + X_i + \varepsilon_{it}$$

Where:
$X_i$ was the relevant policy variable (PStr/PTime/PMean) for testing and contact tracing policies, to control for differences in Covid-19 death definitions

3) Estimate adding date of first case, as a continuous variable, to control for potential virus evolution, and/or advances in scientific understanding over calendar time.

$$Deaths_{it} = \alpha + \beta_{1-9} PStr_i/PTime_i/PMean_i + X_i + \varepsilon_{it}$$

Where:
$X_i$ was date of first death within the country (plus the relevant policy variable [PStr/PTime/PMean] for testing and contact tracing policies), to control for the fact that countries where the outbreak occurred early had less time to prepare

4) Controlling for all additional co-variates, outlined in Table 2 of the manuscript file.

$$Deaths_{it} = \alpha + \beta_{1-9}PStr_i/PTime_i/PMean_i + X_i + \varepsilon_{it}$$

Where:
$X_i$ was a set of country baseline covariates (Table 2, plus the relevant policy variable [PStr/PTime/PMean] for testing and contact tracing policies and date of first case in the country)

5) The fully-adjusted model reported in the main manuscript

$$Deaths_{it} = \alpha + \beta_{1-9}PStr_i/PTime_i/PMean_i + X_i + \delta_t + DoW_{it} + WoY_{it} + \varepsilon_{it}$$

Where:
$\delta_t$ was the day since day of first death (time fixed effect – varied from 0-24/14-38), adjusting for 'point on the curve' – relative time
$DoW_{it}$ was a set of seven dummies indicating which day of the week is the day t for country i
$WoY_{it}$ was a set of dummies indicating the week of the year, both adjusting for seasonality

### Section 2: Scatter plots

*Policy strictness against cumulative Covid-19 deaths (24-days)*

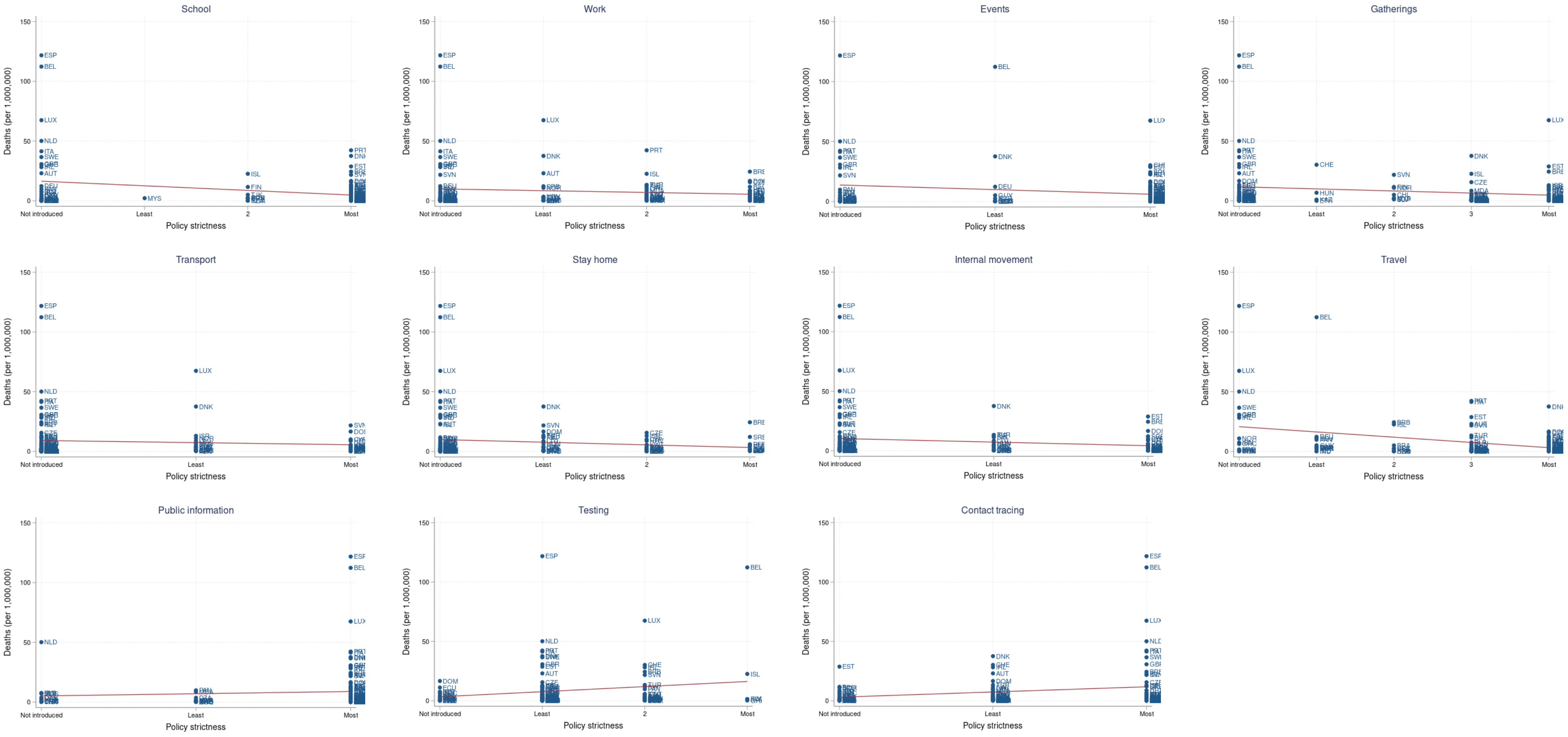

*Policy timing against cumulative Covid-19 deaths (24-days)*

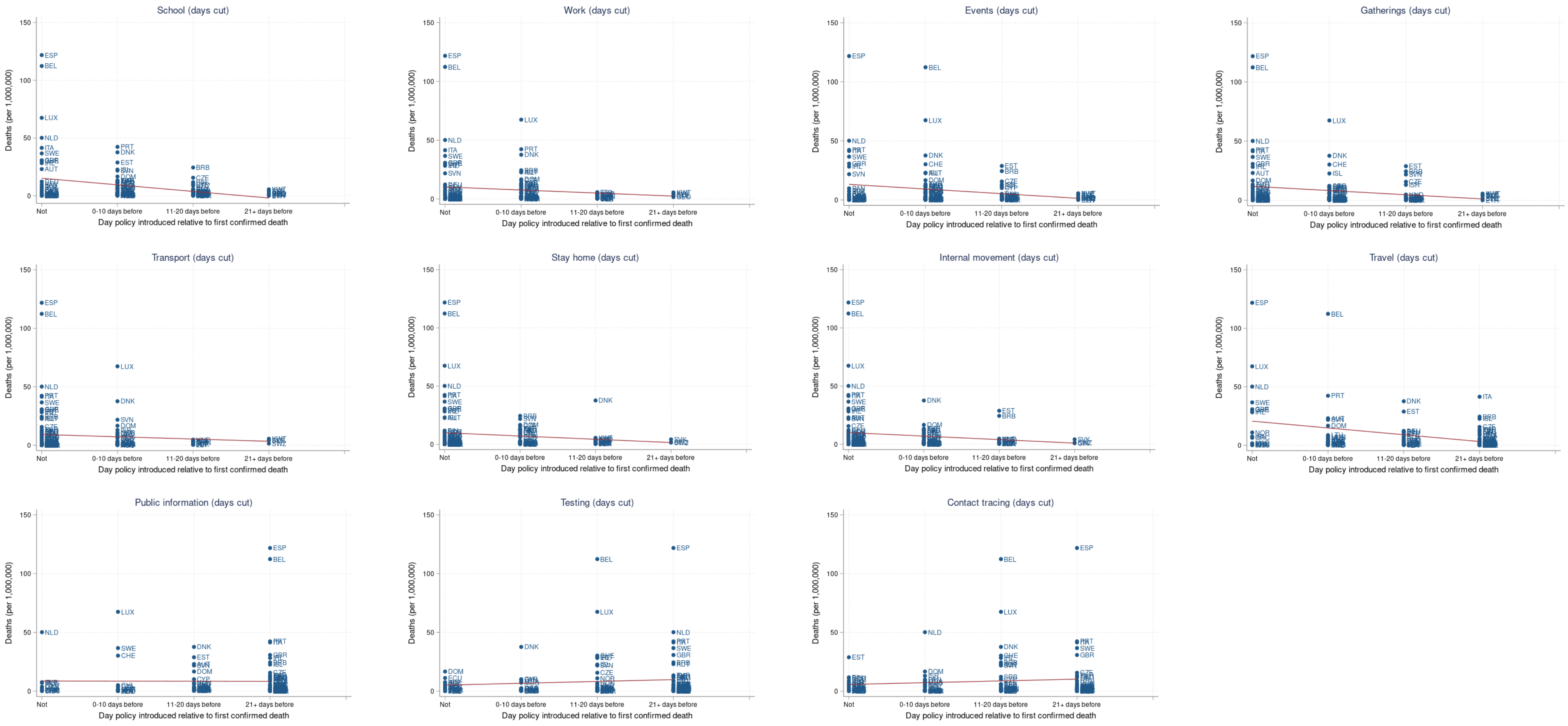

*Mean policy score (combining strictness and timing, larger score indicates earlier/stricter implementation) against cumulative Covid-19 deaths (24-days; policies implemented before first death)*

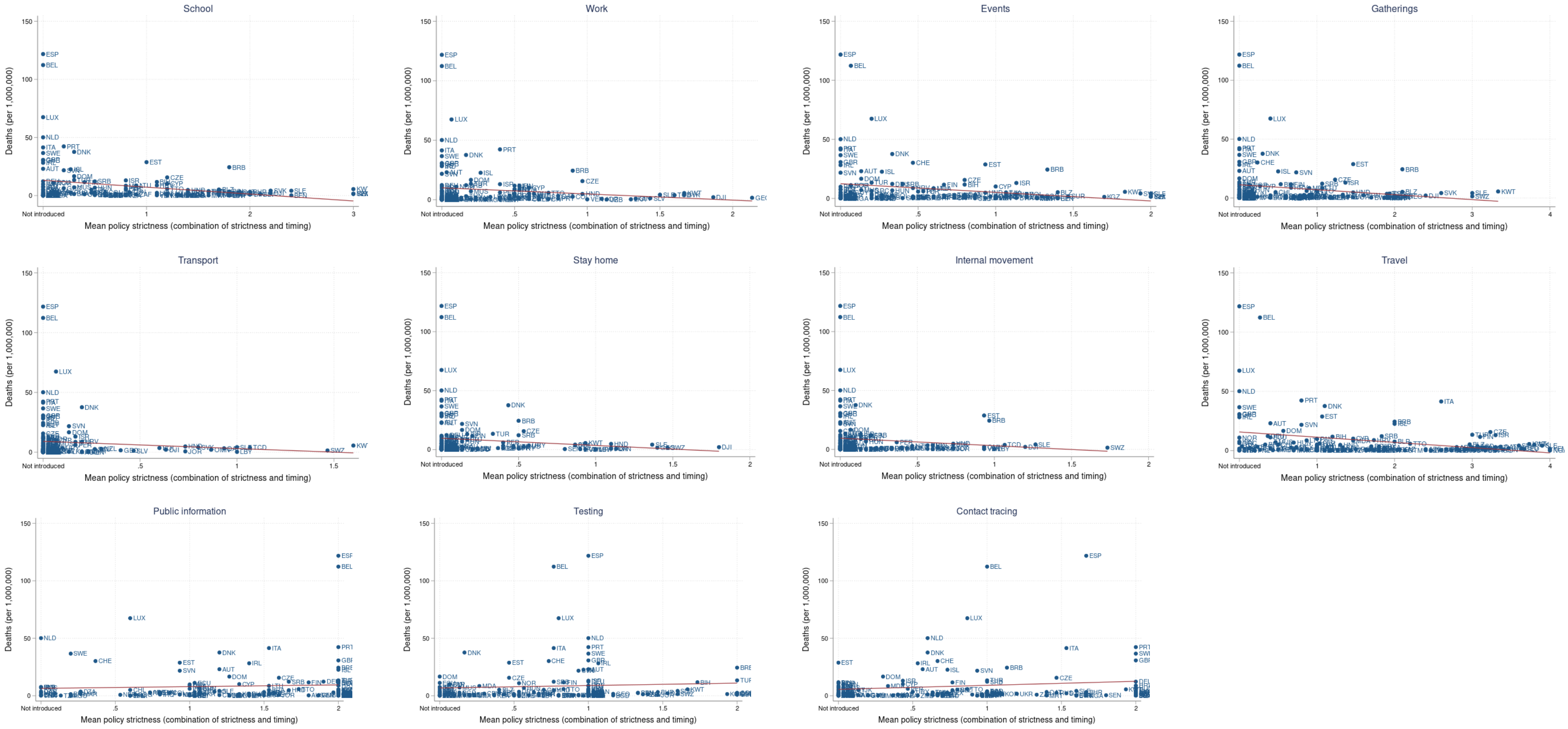

*Mean policy score (combining strictness and timing, larger score indicates earlier/stricter implementation) against cumulative Covid-19 deaths (38-days; policies implemented up to 14 days after first death)*

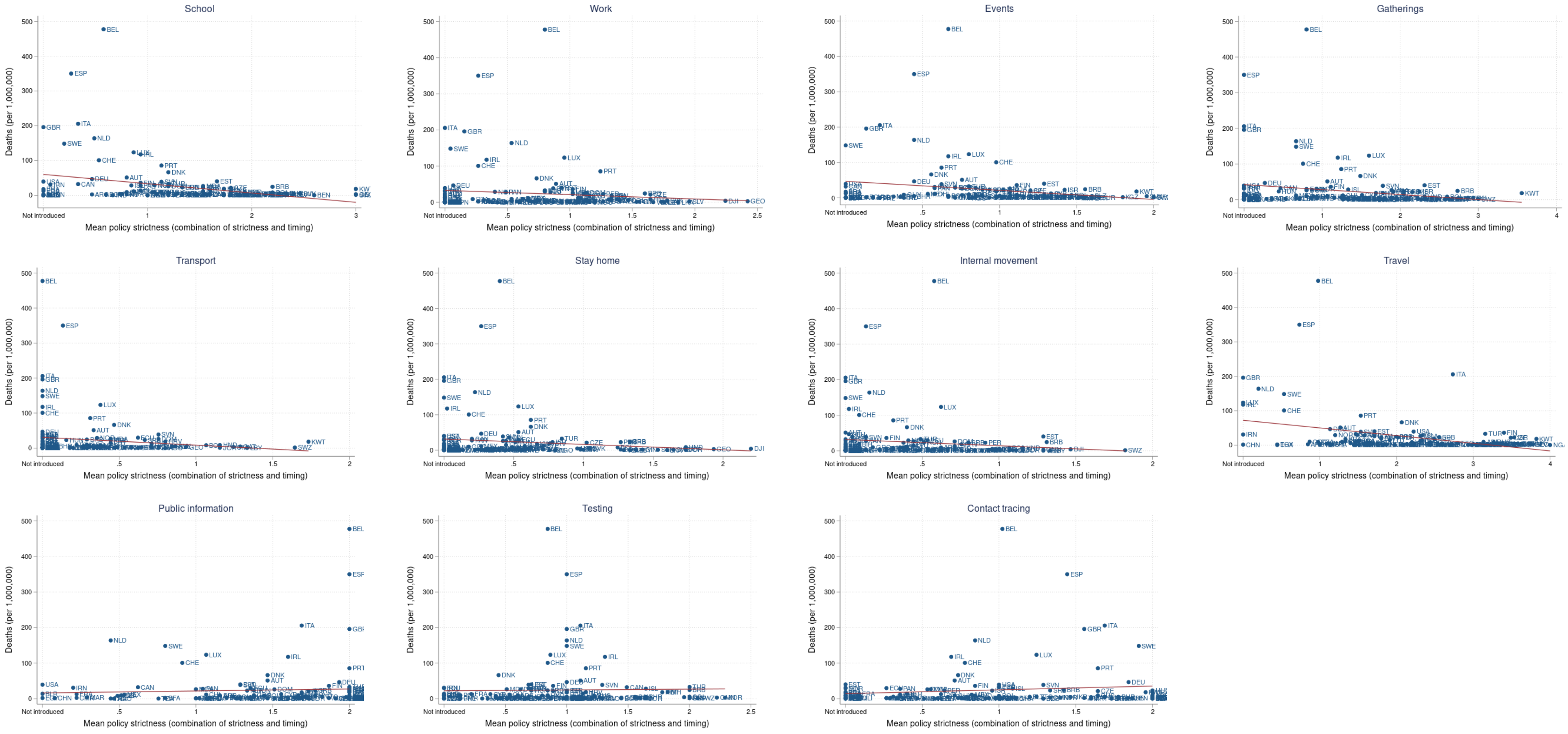

#### Section 3: Covid-19 definition check

Shows the attempt to predict the definition of Covid-19 death adopted by mean policy score across the 28 countries where definition has been classified by the European Observatory.[1] There is no statistically significant policy, except for 'testing policy' itself which strongly predicts whether a testing definition is used, as expected.

*Table 1: Attempting to predict definition of Covid-19 death from mean policy score (policies enacted prior to first death)*

| | (1) | (2) | (3) | (4) | (5) | (6) | (7) | (8) | (9) | (10) | (11) |
|---|---|---|---|---|---|---|---|---|---|---|---|
| **Country using 'testing definition' of Covid-19 death (yes/no)** | OR 95% CI | OR 95% CI | OR 95% CI | OR 95% CI | OR 95% CI | OR 95% CI | OR 95% CI | OR 95% CI | OR 95% CI | OR 95% CI | OR 95% CI |
| School | 0.207 | | | | | | | | | | |
| | (0.023 to 1.834) | | | | | | | | | | |
| Work | | 0.082 | | | | | | | | | |
| | | (0.002 to 3.243) | | | | | | | | | |
| Events | | | 0.190 | | | | | | | | |
| | | | (0.019 to 1.855) | | | | | | | | |
| Gatherings | | | | 0.358 | | | | | | | |
| | | | | (0.082 to 1.558) | | | | | | | |
| Transport | | | | | 0.001 | | | | | | |
| | | | | | (0.000 to 1052.351) | | | | | | |
| Stay home | | | | | | 0.002 | | | | | |
| | | | | | | (0.000 to 8.379) | | | | | |
| Internal movement | | | | | | | 0.003 | | | | |
| | | | | | | | (0.000 to 601.084) | | | | |
| Travel | | | | | | | | 0.542 | | | |
| | | | | | | | | (0.188 to 1.568) | | | |

| | | | | | | | | | | | |
|---|---|---|---|---|---|---|---|---|---|---|---|
| Public information | | | | | | | | | 1.082 | | |
| | | | | | | | | | (0.351 to 3.336) | | |
| Testing | | | | | | | | | | 12.016** | |
| | | | | | | | | | | (2.351 to 61.419) | |
| Contact tracing | | | | | | | | | | | 1.771 |
| | | | | | | | | | | | (0.609 to 5.153) |
| Observations | 28 | 28 | 28 | 28 | 28 | 28 | 28 | 28 | 28 | 28 | 28 |
| Pseudo R-squared | 0.067 | 0.047 | 0.052 | 0.034 | 0.022 | 0.057 | 0.049 | 0.046 | 0.001 | 0.148 | 0.031 |

### Section 4: Regression results

#### Comparison of coefficient plots for robustness checks

**Mean score (combining timing and strictness)**

Main model (linear specification)

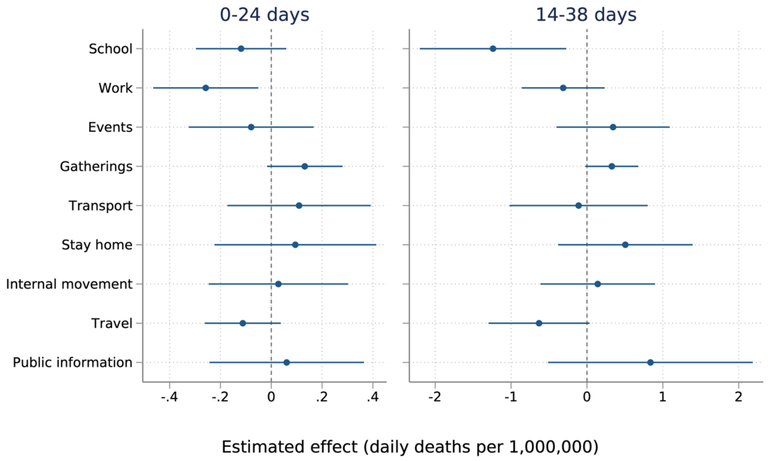

Dropping China/Belgium specification

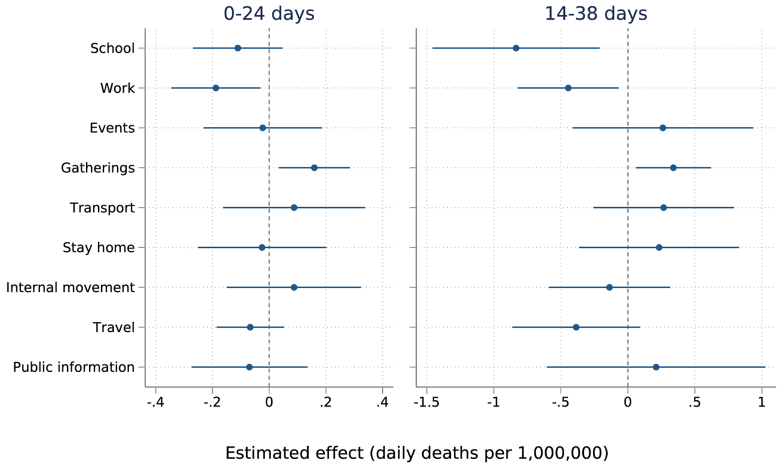

Negative binomial (count model) specification

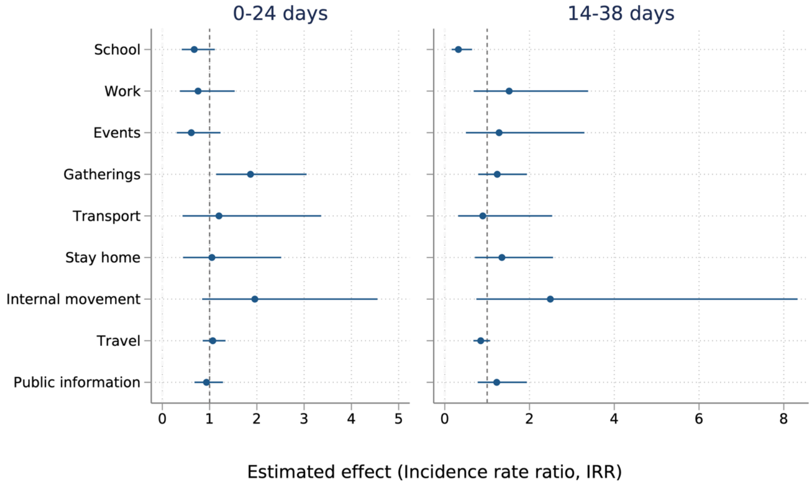

#### Timing alone

##### Main model (linear specification)

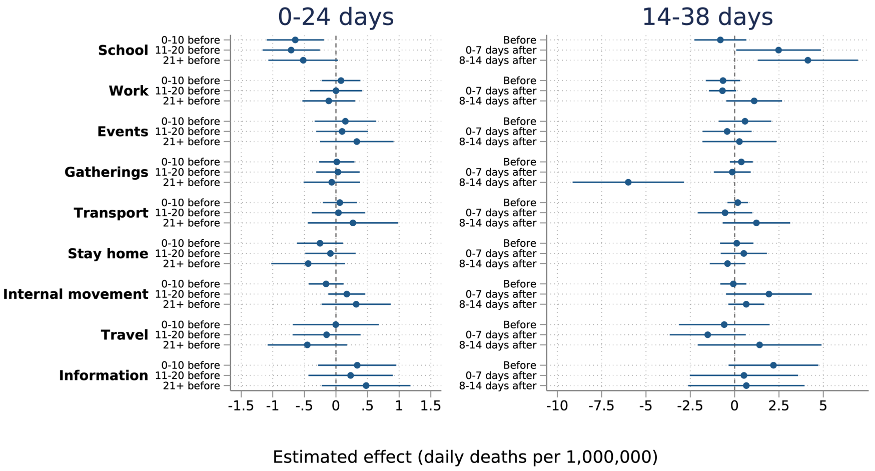

##### Dropping China/Belgium specification

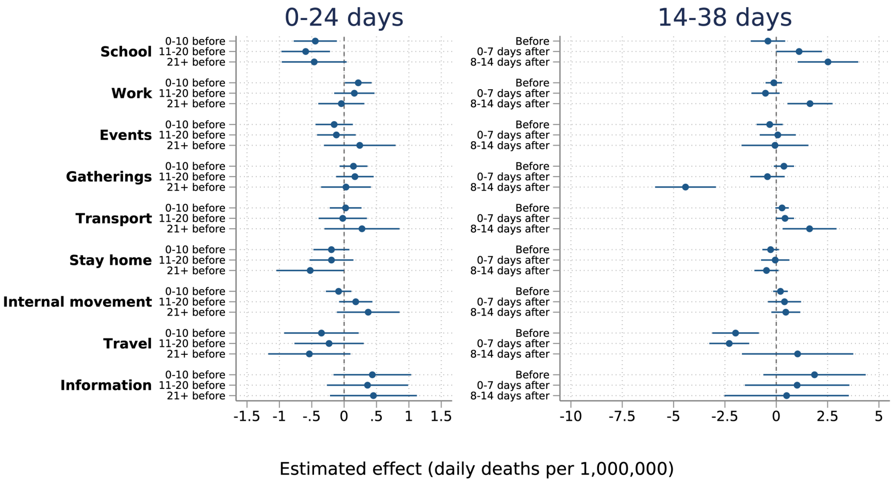

##### Negative binomial (count model) specification

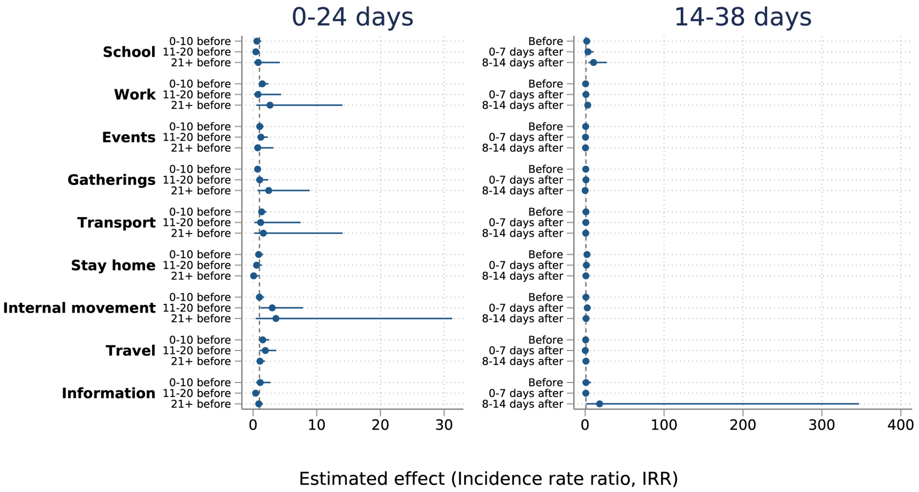

#### Strictness alone

##### Main model (linear specification)

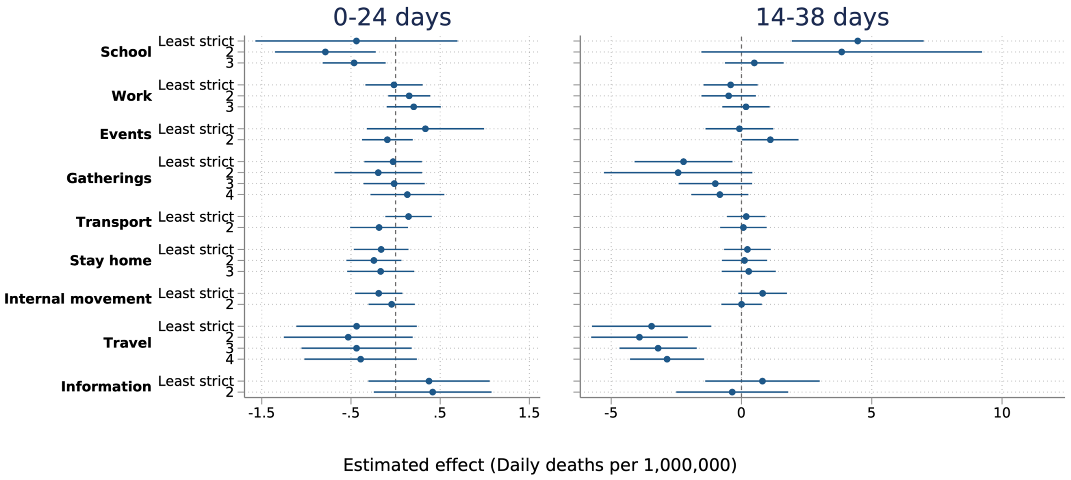

##### Dropping China/Belgium specification

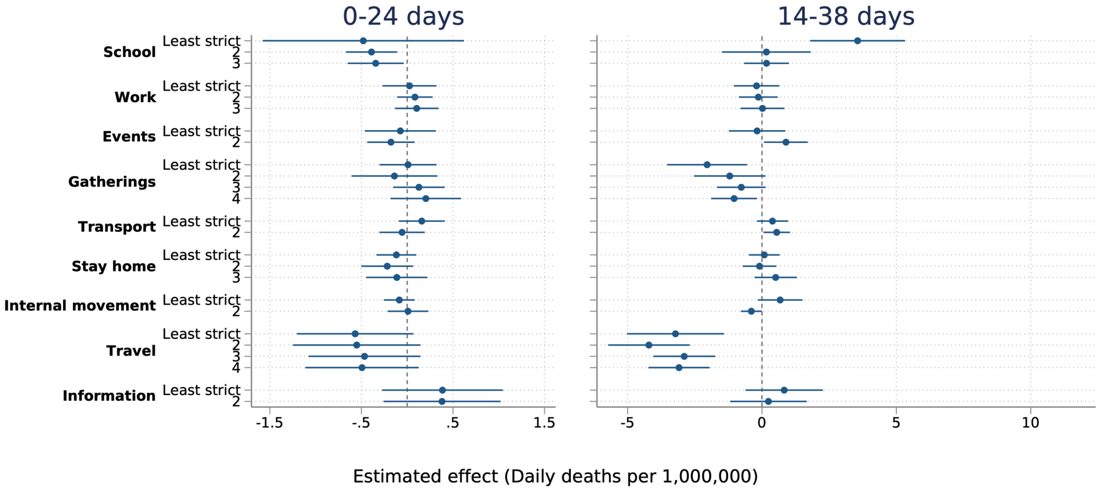

##### Negative binomial (count model) specification

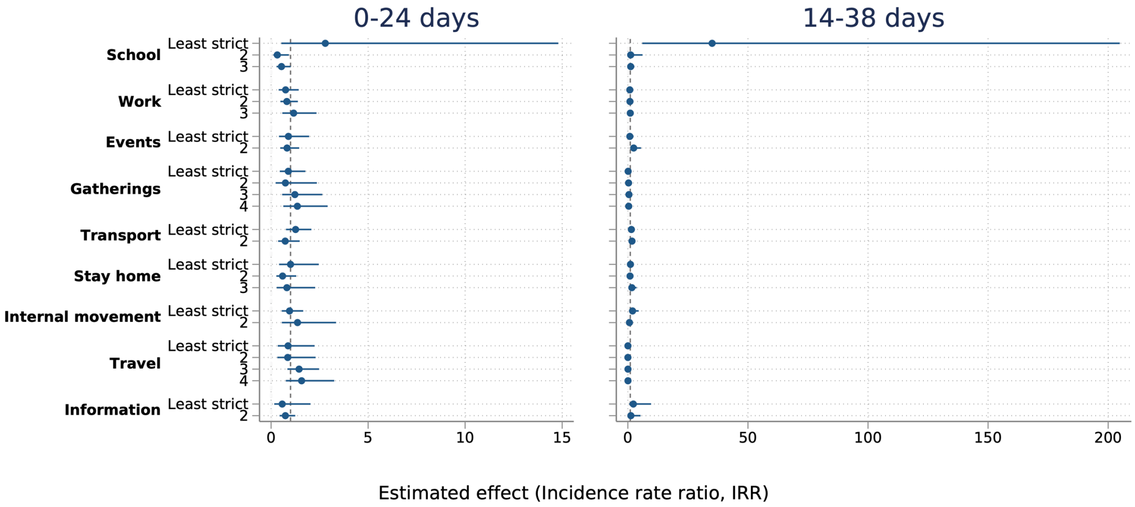

#### Regression results – linear specifications

*Regression results, mean policy strictness (combination of timing and strictness)*

| | 24 Days (Policies implemented before first death) | | | | | 38 Days (Policies implemented within 14 days after first death) | | | | |
|---|---|---|---|---|---|---|---|---|---|---|
| **Average deaths per day over time period, per million** | **(1)** | **(2)** | **(3)** | **(4)** | **(5) #** | **(1)** | **(2)** | **(3)** | **(4)** | **(5) #** |
| **School** | -0.237** | -0.267* | -0.257* | -0.145 | -0.119 | -1.071** | -1.305* | -1.049* | -1.208** | -1.238* |
| | (-0.382 to -0.092) | (-0.477 to -0.057) | (-0.464 to -0.051) | (-0.328 to 0.038) | (-0.297 to 0.059) | (-1.744 to -0.398) | (-2.335 to -0.276) | (-1.863 to -0.236) | (-2.029 to -0.387) | (-2.203 to -0.273) |
| **Work** | -0.207* | -0.007 | -0.005 | -0.198* | -0.258* | -0.525* | 0.105 | 0.087 | -0.223 | -0.313 |
| | (-0.382 to -0.032) | (-0.169 to 0.154) | (-0.162 to 0.152) | (-0.367 to -0.028) | (-0.465 to -0.051) | (-1.016 to -0.033) | (-0.376 to 0.587) | (-0.377 to 0.551) | (-0.797 to 0.352) | (-0.861 to 0.234) |
| **Events** | -0.292** | 0.074 | 0.079 | -0.028 | -0.079 | -1.040** | 0.247 | 0.229 | 0.223 | 0.344 |
| | (-0.493 to -0.090) | (-0.179 to 0.327) | (-0.176 to 0.333) | (-0.231 to 0.175) | (-0.325 to 0.167) | (-1.730 to -0.351) | (-0.595 to 1.089) | (-0.616 to 1.074) | (-0.446 to 0.892) | (-0.403 to 1.091) |
| **Gatherings** | -0.168* | -0.029 | -0.029 | 0.106 | 0.132 | -0.577* | 0.088 | 0.092 | 0.333 | 0.328 |
| | (-0.295 to -0.040) | (-0.217 to 0.160) | (-0.218 to 0.160) | (-0.058 to 0.270) | (-0.017 to 0.280) | (-1.018 to -0.137) | (-0.316 to 0.492) | (-0.303 to 0.486) | (-0.007 to 0.673) | (-0.021 to 0.677) |
| **Transport** | -0.240** | 0.200 | 0.195 | 0.172 | 0.109 | -0.903* | 0.182 | 0.154 | -0.058 | -0.110 |
| | (-0.420 to -0.061) | (-0.117 to 0.518) | (-0.116 to 0.507) | (-0.099 to 0.443) | (-0.172 to 0.391) | (-1.640 to -0.165) | (-0.619 to 0.982) | (-0.623 to 0.930) | (-0.928 to 0.813) | (-1.020 to 0.801) |
| **Stay home** | -0.245** | 0.080 | 0.077 | -0.015 | 0.095 | -0.628** | 0.546 | 0.560 | 0.453 | 0.506 |
| | (-0.423 to -0.067) | (-0.208 to 0.369) | (-0.205 to 0.358) | (-0.263 to 0.233) | (-0.224 to 0.413) | (-1.061 to -0.195) | (-0.164 to 1.257) | (-0.140 to 1.261) | (-0.342 to 1.248) | (-0.380 to 1.392) |
| **Internal movement** | -0.256* | -0.018 | -0.012 | 0.089 | 0.028 | -0.737* | -0.192 | -0.185 | 0.144 | 0.142 |
| | (-0.505 to -0.006) | (-0.390 to 0.353) | (-0.388 to 0.363) | (-0.130 to 0.308) | (-0.246 to 0.302) | (-1.363 to -0.111) | (-1.107 to 0.723) | (-1.065 to 0.696) | (-0.617 to 0.906) | (-0.613 to 0.897) |
| **Travel** | -0.176** | -0.207** | -0.206** | -0.097 | -0.112 | -0.868** | -0.993** | -0.986** | -0.663* | -0.631 |

| | | | | | | | | | | |
|---|---|---|---|---|---|---|---|---|---|---|
| | (-0.294 to -0.059) | (-0.354 to -0.060) | (-0.351 to -0.062) | (-0.226 to 0.031) | (-0.262 to 0.038) | (-1.381 to -0.355) | (-1.586 to -0.401) | (-1.569 to -0.404) | (-1.206 to -0.120) | (-1.296 to 0.034) |
| **Public information** | 0.060 | 0.227 | 0.233 | 0.066 | 0.061 | 0.190 | 1.205 | 1.288 | 0.941 | 0.837 |
| | (-0.135 to 0.256) | (-0.065 to 0.520) | (-0.086 to 0.552) | (-0.237 to 0.369) | (-0.243 to 0.365) | (-0.559 to 0.940) | (-0.014 to 2.424) | (-0.025 to 2.601) | (-0.473 to 2.354) | (-0.511 to 2.186) |
| **Testing** | 0.084 | 0.068 | 0.063 | 0.168 | 0.166 | 0.072 | 0.025 | -0.063 | 0.313 | 0.348 |
| | (-0.037 to 0.205) | (-0.109 to 0.244) | (-0.130 to 0.255) | (-0.000 to 0.336) | (-0.016 to 0.349) | (-0.218 to 0.363) | (-0.460 to 0.511) | (-0.650 to 0.524) | (-0.260 to 0.886) | (-0.258 to 0.953) |
| **Contact tracing** | 0.140 | 0.188* | 0.174* | 0.089 | 0.039 | 0.414 | 0.631* | 0.437 | 0.308 | 0.190 |
| | (-0.026 to 0.306) | (0.010 to 0.365) | (0.001 to 0.347) | (-0.088 to 0.265) | (-0.160 to 0.238) | (-0.039 to 0.867) | (0.119 to 1.143) | (-0.031 to 0.904) | (-0.203 to 0.819) | (-0.462 to 0.841) |
| **Date of first case** | | | -0.001 | 0.000 | -0.005 | | | -0.017 | -0.025 | -0.032 |
| | | | (-0.008 to 0.006) | (-0.013 to 0.014) | (-0.023 to 0.014) | | | (-0.047 to 0.013) | (-0.080 to 0.029) | (-0.099 to 0.035) |
| **Population density (people per sq.km)** | | | | -0.000 | -0.000 | | | | -0.000 | -0.000 |
| | | | | (-0.000 to 0.000) | (-0.000 to 0.000) | | | | (-0.001 to 0.000) | (-0.001 to 0.000) |
| **% Population aged 65+** | | | | 0.010 | 0.007 | | | | -0.037 | -0.030 |
| | | | | (-0.026 to 0.046) | (-0.034 to 0.047) | | | | (-0.162 to 0.088) | (-0.173 to 0.113) |
| **% Population male** | | | | 0.002 | -0.001 | | | | 0.025 | 0.032 |
| | | | | (-0.016 to 0.020) | (-0.023 to 0.020) | | | | (-0.057 to 0.108) | (-0.059 to 0.123) |
| **Life expectancy at birth (years)** | | | | 0.018 | 0.020 | | | | 0.037 | 0.031 |
| | | | | (-0.001 to 0.038) | (-0.001 to 0.042) | | | | (-0.033 to 0.107) | (-0.044 to 0.106) |

| | | | | | | | | | | |
|---|---|---|---|---|---|---|---|---|---|---|
| **Hospital beds (per 1000 people)** | | | | -0.060* | -0.042 | | | | -0.113 | -0.074 |
| | | | | (-0.116 to -0.004) | (-0.104 to 0.020) | | | | (-0.361 to 0.135) | (-0.372 to 0.223) |
| **Physicians (per 1000 people)** | | | | -0.009 | -0.041 | | | | -0.006 | -0.092 |
| | | | | (-0.151 to 0.132) | (-0.189 to 0.107) | | | | (-0.485 to 0.472) | (-0.611 to 0.426) |
| **GDP PPP (current international $)** | | | | -0.000 | -0.000 | | | | -0.000 | -0.000 |
| | | | | (-0.000 to 0.000) | (-0.000 to 0.000) | | | | (-0.000 to 0.000) | (-0.000 to 0.000) |
| **Manufacturing value added (%GDP)** | | | | -0.012 | -0.014 | | | | -0.014 | -0.021 |
| | | | | (-0.027 to 0.003) | (-0.031 to 0.002) | | | | (-0.053 to 0.025) | (-0.064 to 0.021) |
| **Health expenditure (%GDP)** | | | | -0.005 | 0.012 | | | | 0.037 | 0.085 |
| | | | | (-0.062 to 0.051) | (-0.052 to 0.077) | | | | (-0.166 to 0.240) | (-0.133 to 0.303) |
| **International tourism, number of arrivals** | | | | 0.000 | 0.000 | | | | 0.000 | 0.000 |
| | | | | (-0.000 to 0.000) | (-0.000 to 0.000) | | | | (-0.000 to 0.000) | (-0.000 to 0.000) |
| **Governance (Voice and Accountability)** | | | | 0.105 | 0.131 | | | | 0.196 | 0.186 |
| | | | | (-0.046 to 0.257) | (-0.063 to 0.325) | | | | (-0.245 to 0.637) | (-0.352 to 0.724) |
| **East Asia & Pacific** | | | | . | . | | | | . | . |

| | | | | | | | | | | |
|---|---|---|---|---|---|---|---|---|---|---|
| **Europe & Central Asia** | | | | 0.710* | 0.541 | | | | 2.816* | 2.339* |
| | | | | (0.156 to 1.263) | (-0.047 to 1.128) | | | | (0.650 to 4.981) | (0.344 to 4.334) |
| **America & Caribbean** | | | | 0.196 | 0.051 | | | | 1.306 | 0.949 |
| | | | | (-0.138 to 0.530) | (-0.333 to 0.435) | | | | (-0.165 to 2.778) | (-0.538 to 2.435) |
| **East & North Africa** | | | | 0.262 | 0.202 | | | | 1.344 | 1.051 |
| | | | | (-0.081 to 0.604) | (-0.201 to 0.605) | | | | (-0.153 to 2.840) | (-0.513 to 2.614) |
| **North America** | | | | -0.302 | -0.446 | | | | 0.757 | -0.142 |
| | | | | (-0.867 to 0.263) | (-1.138 to 0.246) | | | | (-1.923 to 3.437) | (-2.602 to 2.319) |
| **South Asia** | | | | 0.041 | -0.067 | | | | 0.585 | 0.358 |
| | | | | (-0.298 to 0.379) | (-0.489 to 0.355) | | | | (-0.554 to 1.724) | (-1.141 to 1.857) |
| **Sub-Saharan Africa** | | | | 0.316 | 0.183 | | | | 2.042* | 1.649 |
| | | | | (-0.089 to 0.721) | (-0.235 to 0.601) | | | | (0.153 to 3.931) | (-0.140 to 3.439) |
| **Constant** | | 0.343** | 22.723 | -10.204 | 101.468 | | 1.746*** | 364.592 | 548.863 | 701.113 |
| | | (0.138 to 0.548) | (-140.764 to 186.210) | (-307.511 to 287.104) | (-310.878 to 513.814) | | (0.868 to 2.623) | (-294.613 to 1023.797) | (-646.540 to 1744.267) | (-776.991 to 2179.218) |
| **Observations** | 3250 | 3250 | 3250 | 3250 | 3250 | 3150 | 3150 | 3150 | 3150 | 3150 |
| **R-squared** | | 0.079 | 0.079 | 0.176 | 0.263 | | 0.203 | 0.209 | 0.348 | 0.375 |
| **Adjusted R-squared** | | 0.075 | 0.075 | 0.169 | 0.245 | | 0.201 | 0.206 | 0.342 | 0.360 |

**p-values in parentheses =** * p<0.05 ** p<0.01 *** p<0.001; # includes time, day of week and week of year fixed effects

*Regression results, policy strictness. Baseline is policy not introduced within policy analysis period*

| | 24 Days (Policies implemented before first death) | | | | | 38 Days (Policies implemented within 14 days after first death) | | | | |
|---|---|---|---|---|---|---|---|---|---|---|
| **Average deaths per day over time period, per million** | **(1)** | **(2)** | **(3)** | **(4)** | **(5) #** | **(1)** | **(2)** | **(3)** | **(4)** | **(5) #** |
| **0bn.School** | . | . | . | . | . | . | . | . | . | . |
| **1.School** | -0.585** | -0.387 | -0.394 | 0.032 | -0.438 | 4.899*** | 8.248*** | 9.117*** | 7.737** | 7.011** |
| | (-0.974 to -0.197) | (-0.897 to 0.123) | (-0.916 to 0.128) | (-0.882 to 0.945) | (-1.571 to 0.695) | (3.687 to 6.112) | (4.983 to 11.513) | (5.309 to 12.924) | (2.237 to 13.237) | (1.923 to 12.099) |
| **2.School** | -0.443* | -0.414 | -0.399 | -0.581* | -0.787** | 3.297 | 3.583 | 4.512 | 4.395 | 3.755 |
| | (-0.882 to -0.004) | (-0.940 to 0.113) | (-0.916 to 0.117) | (-1.124 to - 0.037) | (-1.352 to - 0.222) | (-3.267 to 9.860) | (-1.625 to 8.792) | (-0.867 to 9.891) | (-1.380 to 10.171) | (-1.608 to 9.118) |
| **3.School** | -0.470* | -0.331 | -0.319 | -0.337* | -0.464* | -0.203 | 0.071 | 1.061 | 1.007 | 0.402 |
| | (-0.865 to -0.076) | (-0.707 to 0.046) | (-0.691 to 0.054) | (-0.655 to - 0.019) | (-0.817 to - 0.111) | (-1.461 to 1.055) | (-1.802 to 1.945) | (-1.191 to 3.313) | (-0.743 to 2.757) | (-1.259 to 2.063) |
| **0bn.Work** | . | . | . | . | . | . | . | . | . | . |
| **1.Work** | 0.082 | 0.346* | 0.350* | 0.048 | -0.018 | 0.624 | 0.090 | 0.016 | -0.360 | -0.370 |
| | (-0.371 to 0.535) | (0.033 to 0.658) | (0.036 to 0.663) | (-0.245 to 0.342) | (-0.338 to 0.303) | (-0.602 to 1.850) | (-1.057 to 1.237) | (-1.089 to 1.120) | (-1.359 to 0.638) | (-1.357 to 0.616) |
| **2.Work** | -0.125 | 0.377* | 0.375* | 0.186 | 0.153 | 0.588 | 0.345 | 0.205 | -0.519 | -0.695 |
| | (-0.386 to 0.136) | (0.034 to 0.719) | (0.032 to 0.718) | (-0.057 to 0.429) | (-0.083 to 0.388) | (-0.601 to 1.777) | (-0.989 to 1.679) | (-1.040 to 1.450) | (-1.445 to 0.406) | (-1.699 to 0.309) |
| **3.Work** | -0.172 | 0.505** | 0.506** | 0.235 | 0.204 | 0.375 | 0.593 | 0.530 | 0.132 | -0.098 |
| | (-0.408 to 0.064) | (0.159 to 0.852) | (0.158 to 0.853) | (-0.063 to 0.533) | (-0.100 to 0.507) | (-0.513 to 1.262) | (-0.722 to 1.907) | (-0.720 to 1.779) | (-0.839 to 1.102) | (-1.066 to 0.870) |
| **0bn.Events** | . | . | . | . | . | . | . | . | . | . |
| **1.Events** | 0.176 | 0.144 | 0.145 | 0.319 | 0.335 | -0.242 | -0.416 | -0.467 | -0.264 | -0.061 |
| | (-0.729 to 1.081) | (-0.579 to 0.867) | (-0.575 to 0.866) | (-0.324 to 0.962) | (-0.322 to 0.992) | (-1.336 to 0.852) | (-2.616 to 1.784) | (-2.620 to 1.687) | (-1.527 to 1.000) | (-1.473 to 1.351) |
| **2.Events** | -0.281 | -0.099 | -0.099 | -0.030 | -0.092 | 0.179 | 1.030 | 1.152 | 0.969 | 1.269* |
| | (-0.615 to 0.053) | (-0.468 to 0.270) | (-0.468 to 0.270) | (-0.299 to 0.240) | (-0.377 to 0.193) | (-0.748 to 1.106) | (-0.545 to 2.606) | (-0.462 to 2.766) | (-0.105 to 2.042) | (0.088 to 2.451) |

| | | | | | | | | | | |
|---|---|---|---|---|---|---|---|---|---|---|
| **0bn.Gatherings** | . | . | . | . | . | . | . | . | . | . |
| **1.Gatherings** | -0.110 | 0.028 | 0.036 | -0.107 | -0.027 | -1.070 | -1.121 | -1.034 | -2.177* | -2.336* |
| | (-0.664 to 0.444) | (-0.275 to 0.330) | (-0.269 to 0.341) | (-0.392 to 0.179) | (-0.350 to 0.295) | (-2.408 to 0.267) | (-3.496 to 1.254) | (-3.035 to 0.966) | (-4.084 to -0.271) | (-4.299 to -0.373) |
| **2.Gatherings** | -0.179 | -0.028 | -0.035 | -0.114 | -0.194 | 0.024 | -3.349 | -4.120* | -3.040 | -2.523 |
| | (-0.517 to 0.160) | (-0.414 to 0.359) | (-0.441 to 0.371) | (-0.598 to 0.370) | (-0.685 to 0.297) | (-2.333 to 2.380) | (-6.815 to 0.117) | (-8.047 to -0.193) | (-6.513 to 0.433) | (-5.784 to 0.738) |
| **3.Gatherings** | -0.338* | -0.144 | -0.136 | 0.000 | -0.016 | -1.031 | -2.403 | -2.036 | -1.052 | -0.980 |
| | (-0.624 to -0.053) | (-0.427 to 0.140) | (-0.423 to 0.151) | (-0.322 to 0.323) | (-0.359 to 0.327) | (-2.381 to 0.318) | (-4.941 to 0.135) | (-4.116 to 0.045) | (-2.529 to 0.426) | (-2.432 to 0.472) |
| **4.Gatherings** | -0.213 | 0.133 | 0.132 | 0.164 | 0.132 | -0.529 | -1.741 | -1.726 | -0.876 | -0.862 |
| | (-0.532 to 0.106) | (-0.128 to 0.394) | (-0.129 to 0.393) | (-0.217 to 0.544) | (-0.282 to 0.546) | (-1.982 to 0.923) | (-4.283 to 0.801) | (-3.996 to 0.544) | (-1.996 to 0.244) | (-1.964 to 0.240) |
| **0bn.Transport** | . | . | . | . | . | . | . | . | . | . |
| **1.Transport** | 0.024 | 0.041 | 0.042 | 0.153 | 0.146 | -0.161 | -0.297 | -0.082 | 0.185 | 0.069 |
| | (-0.322 to 0.371) | (-0.207 to 0.289) | (-0.207 to 0.290) | (-0.096 to 0.402) | (-0.115 to 0.406) | (-1.273 to 0.950) | (-1.398 to 0.805) | (-1.152 to 0.989) | (-0.565 to 0.934) | (-0.710 to 0.847) |
| **2.Transport** | -0.172 | -0.146 | -0.148 | -0.117 | -0.185 | -0.833* | -0.475 | -0.521 | -0.060 | -0.058 |
| | (-0.360 to 0.016) | (-0.446 to 0.154) | (-0.453 to 0.157) | (-0.415 to 0.182) | (-0.509 to 0.140) | (-1.544 to -0.122) | (-1.482 to 0.532) | (-1.510 to 0.468) | (-1.135 to 1.015) | (-1.121 to 1.004) |
| **0bn.Stay home** | . | . | . | . | . | . | . | . | . | . |
| **1.Stay home** | -0.059 | 0.071 | 0.064 | -0.122 | -0.162 | 0.050 | 0.669 | 0.665 | 0.387 | 0.306 |
| | (-0.323 to 0.205) | (-0.212 to 0.353) | (-0.226 to 0.354) | (-0.387 to 0.144) | (-0.468 to 0.145) | (-0.830 to 0.929) | (-0.361 to 1.698) | (-0.300 to 1.630) | (-0.585 to 1.359) | (-0.658 to 1.271) |
| **2.Stay home** | -0.251* | -0.167 | -0.173 | -0.201 | -0.243 | 0.305 | 0.828 | 0.750 | 0.260 | 0.279 |
| | (-0.455 to -0.046) | (-0.393 to 0.059) | (-0.410 to 0.064) | (-0.476 to 0.073) | (-0.552 to 0.066) | (-0.820 to 1.431) | (-0.412 to 2.067) | (-0.353 to 1.853) | (-0.610 to 1.129) | (-0.631 to 1.190) |
| **3.Stay home** | -0.203 | -0.183 | -0.188 | -0.160 | -0.167 | -0.666* | 0.317 | 0.346 | 0.294 | 0.507 |
| | (-0.446 to 0.041) | (-0.501 to 0.135) | (-0.514 to 0.137) | (-0.514 to 0.193) | (-0.543 to 0.208) | (-1.318 to -0.014) | (-1.179 to 1.812) | (-1.069 to 1.760) | (-0.679 to 1.266) | (-0.547 to 1.561) |
| **0bn.Internal movement** | . | . | . | . | . | . | . | . | . | . |

| | | | | | | | | | | |
|---|---|---|---|---|---|---|---|---|---|---|
| **1.Internal movement** | -0.130 | 0.035 | 0.036 | -0.173 | -0.189 | 0.850 | 1.266 | 0.985 | 0.781 | 0.882 |
| | (-0.388 to 0.128) | (-0.214 to 0.284) | (-0.212 to 0.283) | (-0.429 to 0.084) | (-0.454 to 0.077) | (-0.476 to 2.175) | (-0.092 to 2.624) | (-0.166 to 2.135) | (-0.026 to 1.587) | (-0.085 to 1.848) |
| **2.Internal movement** | -0.240* | -0.074 | -0.070 | -0.057 | -0.044 | -0.156 | 0.120 | 0.074 | 0.055 | 0.052 |
| | (-0.453 to -0.028) | (-0.400 to 0.252) | (-0.399 to 0.259) | (-0.299 to 0.185) | (-0.303 to 0.215) | (-1.019 to 0.707) | (-0.827 to 1.067) | (-0.859 to 1.007) | (-0.782 to 0.891) | (-0.785 to 0.889) |
| **0bn.Travel** | . | . | . | . | . | . | . | . | . | . |
| **1.Travel** | -0.465 | -0.508 | -0.508 | -0.250 | -0.437 | -3.572** | -1.803 | -0.099 | -3.184* | -3.278** |
| | (-1.366 to 0.436) | (-1.119 to 0.102) | (-1.117 to 0.101) | (-0.839 to 0.338) | (-1.112 to 0.239) | (-5.997 to -1.146) | (-4.374 to 0.768) | (-3.711 to 3.512) | (-5.965 to -0.403) | (-5.732 to -0.825) |
| **2.Travel** | -0.728* | -0.716** | -0.715** | -0.479 | -0.530 | -3.434** | -2.682* | -2.358 | -4.176*** | -3.720*** |
| | (-1.410 to -0.046) | (-1.252 to -0.181) | (-1.248 to -0.182) | (-1.187 to 0.229) | (-1.253 to 0.192) | (-5.866 to -1.002) | (-5.032 to -0.331) | (-5.022 to 0.305) | (-6.216 to -2.135) | (-5.732 to -1.708) |
| **3.Travel** | -0.708* | -0.785* | -0.785* | -0.330 | -0.437 | -2.438 | -2.236* | -1.926 | -3.083*** | -3.086*** |
| | (-1.363 to -0.053) | (-1.420 to -0.151) | (-1.419 to -0.151) | (-0.916 to 0.256) | (-1.055 to 0.181) | (-4.960 to 0.083) | (-4.359 to -0.114) | (-4.389 to 0.536) | (-4.847 to -1.319) | (-4.650 to -1.523) |
| **4.Travel** | -0.824* | -0.817** | -0.810** | -0.291 | -0.391 | -2.924* | -2.624* | -2.156 | -2.838** | -2.709*** |
| | (-1.464 to -0.185) | (-1.434 to -0.201) | (-1.422 to -0.197) | (-0.913 to 0.331) | (-1.021 to 0.239) | (-5.411 to -0.437) | (-4.660 to -0.588) | (-4.630 to 0.318) | (-4.511 to -1.166) | (-4.251 to -1.168) |
| **0bn. Public information** | . | . | . | . | . | . | . | . | . | . |
| **1.Public information** | -0.169 | 0.309 | 0.326 | 0.380 | 0.375 | -0.246 | 1.088 | 1.305 | 0.980 | 0.785 |
| | (-0.486 to 0.148) | (-0.256 to 0.874) | (-0.297 to 0.949) | (-0.251 to 1.011) | (-0.305 to 1.055) | (-1.085 to 0.592) | (-1.076 to 3.253) | (-1.090 to 3.701) | (-1.045 to 3.005) | (-1.439 to 3.008) |
| **2.Public information** | 0.085 | 0.485 | 0.493 | 0.375 | 0.416 | 0.238 | 1.221 | 1.335 | -0.214 | -0.486 |
| | (-0.252 to 0.422) | (-0.091 to 1.061) | (-0.115 to 1.102) | (-0.248 to 0.997) | (-0.243 to 1.076) | (-0.631 to 1.107) | (-1.006 to 3.448) | (-0.909 to 3.579) | (-1.811 to 1.384) | (-2.602 to 1.629) |
| **0bn.Testing** | . | . | . | . | . | . | . | . | . | . |
| **1.Testing** | 0.180* | -0.055 | -0.058 | -0.128 | -0.186 | 0.419 | 0.221 | 0.232 | 0.652 | 0.443 |

| | | | | | | | | | | |
|---|---|---|---|---|---|---|---|---|---|---|
| | (0.005 to 0.356) | (-0.333 to 0.224) | (-0.341 to 0.226) | (-0.444 to 0.188) | (-0.516 to 0.144) | (-0.190 to 1.029) | (-0.664 to 1.106) | (-0.632 to 1.097) | (-0.232 to 1.535) | (-0.419 to 1.305) |
| **2.Testing** | 0.187 | -0.061 | -0.061 | -0.026 | -0.074 | 0.457 | 0.263 | 0.245 | 0.349 | 0.297 |
| | (-0.029 to 0.403) | (-0.353 to 0.230) | (-0.353 to 0.231) | (-0.323 to 0.271) | (-0.416 to 0.269) | (-0.085 to 1.000) | (-0.644 to 1.171) | (-0.602 to 1.092) | (-0.511 to 1.210) | (-0.559 to 1.153) |
| **3.Testing** | 0.956 | 0.845 | 0.837 | 0.856 | 0.903 | 1.812 | 1.797 | 1.360 | 2.162 | 2.333 |
| | (-0.581 to 2.492) | (-0.464 to 2.154) | (-0.468 to 2.142) | (-0.216 to 1.927) | (-0.160 to 1.966) | (-1.338 to 4.962) | (-0.305 to 3.899) | (-0.508 to 3.227) | (-0.214 to 4.537) | (-0.108 to 4.775) |
| **0bn.Contact tracing** | . | . | . | . | . | . | . | . | . | . |
| **1.Contact tracing** | 0.060 | 0.144 | 0.144 | 0.225* | 0.246* | 0.157 | 0.622 | 0.503 | 0.366 | 0.481 |
| | (-0.075 to 0.194) | (-0.094 to 0.381) | (-0.093 to 0.380) | (0.021 to 0.429) | (0.036 to 0.457) | (-0.174 to 0.488) | (-0.428 to 1.672) | (-0.493 to 1.498) | (-0.711 to 1.443) | (-0.591 to 1.553) |
| **2.Contact tracing** | 0.326* | 0.279 | 0.275 | 0.275* | 0.237 | 0.933* | 1.080 | 0.830 | 0.976 | 1.138* |
| | (0.055 to 0.598) | (-0.006 to 0.564) | (-0.010 to 0.559) | (0.021 to 0.529) | (-0.010 to 0.484) | (0.074 to 1.793) | (-0.236 to 2.397) | (-0.335 to 1.994) | (-0.088 to 2.041) | (0.019 to 2.258) |
| **Date of first case** | | | -0.001 | 0.003 | -0.001 | | | -0.037* | -0.025 | -0.012 |
| | | | (-0.008 to 0.006) | (-0.005 to 0.012) | (-0.011 to 0.009) | | | (-0.071 to -0.002) | (-0.063 to 0.013) | (-0.051 to 0.027) |
| **Population density (people per sq.km)** | | | | -0.000 | -0.000 | | | | -0.000 | -0.000 |
| | | | | (-0.000 to 0.000) | (-0.000 to 0.000) | | | | (-0.000 to 0.000) | (-0.000 to 0.000) |
| **% Population aged 65+** | | | | 0.042* | 0.046* | | | | 0.035 | 0.048 |
| | | | | (0.010 to 0.073) | (0.011 to 0.081) | | | | (-0.083 to 0.154) | (-0.082 to 0.177) |
| **% Population male** | | | | 0.001 | -0.001 | | | | 0.041 | 0.061 |
| | | | | (-0.021 to 0.024) | (-0.027 to 0.026) | | | | (-0.052 to 0.134) | (-0.038 to 0.161) |
| **Life expectancy at birth (years)** | | | | 0.028* | 0.026 | | | | -0.037 | -0.050 |
| | | | | (0.002 to 0.053) | (-0.000 to 0.053) | | | | (-0.135 to 0.060) | (-0.158 to 0.059) |

| | | | | | | | | | | |
|---|---|---|---|---|---|---|---|---|---|---|
| **Hospital beds (per 1000 people)** | | | | -0.088*** | -0.064** | | | | -0.160 | -0.121 |
| | | | | (-0.139 to -0.036) | (-0.109 to -0.019) | | | | (-0.346 to 0.026) | (-0.320 to 0.078) |
| **Physicians (per 1000 people)** | | | | -0.002 | -0.056 | | | | -0.237 | -0.242 |
| | | | | (-0.148 to 0.144) | (-0.201 to 0.090) | | | | (-0.718 to 0.243) | (-0.727 to 0.242) |
| **GDP PPP (current international $)** | | | | -0.000 | -0.000 | | | | -0.000*** | -0.000** |
| | | | | (-0.000 to 0.000) | (-0.000 to 0.000) | | | | (-0.000 to -0.000) | (-0.000 to -0.000) |
| **Manufacturing value added (%GDP)** | | | | -0.01 | -0.014 | | | | -0.022 | -0.029 |
| | | | | (-0.024 to 0.004) | (-0.030 to 0.001) | | | | (-0.061 to 0.016) | (-0.067 to 0.010) |
| **Health expenditure (%GDP)** | | | | -0.018 | 0.007 | | | | 0.260 | 0.284 |
| | | | | (-0.063 to 0.026) | (-0.045 to 0.058) | | | | (-0.078 to 0.598) | (-0.057 to 0.626) |
| **International tourism, number of arrivals** | | | | 0 | 0 | | | | 0.000** | 0.000** |
| | | | | (-0.000 to 0.000) | (-0.000 to 0.000) | | | | (0.000 to 0.000) | (0.000 to 0.000) |
| **Governance (Voice and Accountability)** | | | | -0.032 | -0.023 | | | | -0.031 | -0.078 |
| | | | | (-0.225 to 0.161) | (-0.222 to 0.176) | | | | (-0.590 to 0.529) | (-0.693 to 0.537) |
| **East Asia & Pacific** | | | | . | . | | | | . | . |
| **Europe & Central Asia** | | | | 0.672* | 0.433 | | | | 0.491 | 0.145 |
| | | | | (0.141 to 1.204) | (-0.176 to 1.043) | | | | (-1.221 to 2.204) | (-1.563 to 1.853) |

| | | | | | | | | | | |
|---|---|---|---|---|---|---|---|---|---|---|
| **America & Caribbean** | | | | 0.262 | 0.040 | | | | -0.089 | -0.309 |
| | | | | (-0.039 to 0.562) | (-0.304 to 0.383) | | | | (-1.537 to 1.359) | (-1.755 to 1.138) |
| **East & North Africa** | | | | 0.258 | 0.216 | | | | -1.304 | -1.422 |
| | | | | (-0.098 to 0.614) | (-0.209 to 0.642) | | | | (-3.419 to 0.811) | (-3.460 to 0.616) |
| **North America** | | | | -0.002 | -0.192 | | | | -2.668 | -2.749 |
| | | | | (-0.845 to 0.842) | (-1.180 to 0.796) | | | | (-6.860 to 1.524) | (-6.700 to 1.201) |
| **South Asia** | | | | 0.164 | -0.005 | | | | -0.017 | 0.301 |
| | | | | (-0.225 to 0.554) | (-0.388 to 0.378) | | | | (-1.754 to 1.721) | (-1.333 to 1.934) |
| **Sub-Saharan Africa** | | | | 0.591** | 0.357 | | | | -0.878 | -1.002 |
| | | | | (0.205 to 0.977) | (-0.022 to 0.736) | | | | (-2.973 to 1.217) | (-3.210 to 1.206) |
| **Constant** | . | 0.579* | 18.767 | -77.446 | 20.218 | | 0.518 | 805.782* | 548.275 | 261.603 |
| | . | (0.103 to 1.055) | (-135.955 to 173.489) | (-263.726 to 108.833) | (-195.442 to 235.879) | . | (-3.083 to 4.118) | (53.116 to 1558.447) | (-283.477 to 1380.027) | (-599.685 to 1122.891) |
| **Observations** | 3250 | 3250 | 3250 | 3250 | 3250 | 3150 | 3150 | 3150 | 3150 | 3150 |
| **R-squared** | . | 0.124 | 0.124 | 0.223 | 0.330 | . | 0.253 | 0.283 | 0.445 | 0.482 |
| **Adjusted R-squared** | . | 0.116 | 0.116 | 0.211 | 0.309 | . | 0.246 | 0.276 | 0.437 | 0.466 |

**p-values in parentheses** = * p<0.05 ** p<0.01 *** p<0.001; # includes time, day of week and week of year fixed effects

*Regression results, policy timing. Baseline is policy not introduced within policy analysis period.*

| | 24 Days (Policies implemented before first death) | | | | | | 38 Days (Policies implemented within 14 days after first death) | | | | |
|---|---|---|---|---|---|---|---|---|---|---|---|
| **Average deaths per day over time period, per million** | **(1)** | **(2)** | **(3)** | **(4)** | **(5) #** | | **(1)** | **(2)** | **(3)** | **(4)** | **(5) #** |
| **School (Not introduced)** | . | . | . | . | . | **School (Not introduced)** | . | . | . | . | . |
| **School (0-10 days before)** | -0.371 | -0.430 | -0.377 | -0.482** | -0.644** | **School (before)** | -0.594 | -1.516 | -0.874 | -1.997 | -2.526 |
| | (-0.777 to 0.036) | (-0.872 to 0.012) | (-0.769 to 0.014) | (-0.843 to -0.121) | (-1.097 to -0.191) | | (-1.810 to 0.623) | (-4.156 to 1.125) | (-3.287 to 1.538) | (-4.510 to 0.515) | (-5.255 to 0.202) |
| **School (11-20 days before)** | -0.544** | -0.642** | -0.573** | -0.565** | -0.709** | **School (0-7 days after)** | 1.942 | 0.927 | 1.501 | 0.185 | 0.005 |
| | (-0.937 to -0.151) | (-1.080 to -0.203) | (-0.951 to -0.195) | (-0.932 to -0.199) | (-1.162 to -0.256) | | (-0.552 to 4.435) | (-1.193 to 3.047) | (-0.771 to 3.774) | (-1.420 to 1.790) | (-1.688 to 1.698) |
| **School (20+ days before)** | -0.586** | -0.323 | -0.188 | -0.417 | -0.518 | **School (8-14 days after)** | 2.837 | 3.646* | 3.778* | 3.070** | 3.286** |
| | (-0.977 to -0.195) | (-0.751 to 0.104) | (-0.629 to 0.253) | (-0.919 to 0.086) | (-1.070 to 0.034) | | (-0.942 to 6.616) | (0.137 to 7.155) | (0.289 to 7.267) | (1.140 to 4.999) | (1.219 to 5.353) |
| **Work (Not introduced)** | . | . | . | . | . | **Work (Not introduced)** | . | . | . | . | . |
| **Work (0-10 days before)** | -0.002 | 0.190 | 0.199 | 0.093 | 0.080 | **Work (before)** | -0.099 | -0.335 | -0.279 | -0.770 | -1.020 |
| | (-0.272 to 0.268) | (-0.090 to 0.471) | (-0.075 to 0.473) | (-0.168 to 0.353) | (-0.226 to 0.386) | | (-0.682 to 0.484) | (-1.275 to 0.605) | (-1.173 to 0.615) | (-1.765 to 0.225) | (-2.066 to 0.027) |
| **Work (11-20 days before)** | -0.303** | -0.091 | -0.099 | -0.033 | 0.001 | **Work (0-7 days after)** | 0.830 | -0.630 | -0.691 | -0.556 | -0.928* |
| | (-0.523 to -0.083) | (-0.482 to 0.300) | (-0.497 to 0.299) | (-0.403 to 0.337) | (-0.414 to 0.415) | | (-0.684 to 2.343) | (-1.831 to 0.572) | (-1.888 to 0.506) | (-1.465 to 0.352) | (-1.804 to -0.053) |
| **Work (20+ days before)** | -0.239* | 0.030 | 0.033 | -0.133 | -0.114 | **Work (8-14 days after)** | 3.633* | 2.259* | 2.106 | 1.152 | 0.670 |
| | (-0.463 to -0.016) | (-0.242 to 0.303) | (-0.233 to 0.300) | (-0.514 to 0.248) | (-0.531 to 0.304) | | (0.460 to 6.806) | (0.079 to 4.438) | (-0.102 to 4.314) | (-0.472 to 2.777) | (-1.040 to 2.380) |

| | | | | | | | | | | | |
|---|---|---|---|---|---|---|---|---|---|---|---|
| **Events (Not introduced)** | . | . | . | . | . | **Events (Not introduced)** | . | . | . | . | . |
| **Events (0-10 days before)** | -0.096 | 0.144 | 0.158 | 0.170 | 0.149 | **Events (before)** | -0.169 | 0.859 | 0.610 | 1.148 | 1.608 |
| | (-0.492 to 0.300) | (-0.380 to 0.667) | (-0.372 to 0.689) | (-0.298 to 0.637) | (-0.338 to 0.636) | | (-1.061 to 0.723) | (-1.655 to 3.372) | (-1.759 to 2.980) | (-0.956 to 3.252) | (-0.669 to 3.885) |
| **Events (11-20 days before)** | -0.341* | 0.035 | 0.072 | 0.147 | 0.097 | **Events (0-7 days after)** | 1.405 | 0.031 | -0.263 | -0.035 | 0.706 |
| | (-0.676 to -0.005) | (-0.391 to 0.460) | (-0.396 to 0.540) | (-0.283 to 0.576) | (-0.313 to 0.508) | | (-0.585 to 3.395) | (-2.020 to 2.082) | (-2.331 to 1.805) | (-1.258 to 1.187) | (-0.698 to 2.110) |
| **Events (20+ days before)** | -0.415* | -0.129 | -0.107 | 0.306 | 0.329 | **Events (8-14 days after)** | 2.482 | -0.196 | -0.203 | -0.216 | 0.496 |
| | (-0.742 to -0.088) | (-0.541 to 0.283) | (-0.569 to 0.354) | (-0.233 to 0.845) | (-0.252 to 0.911) | | (-1.014 to 5.978) | (-3.874 to 3.481) | (-3.754 to 3.348) | (-1.994 to 1.561) | (-1.455 to 2.447) |
| **Gatherings (Not introduced)** | . | . | . | . | . | **Gatherings (Not introduced)** | . | . | . | . | . |
| **Gatherings (0-10 days before)** | -0.198 | -0.031 | -0.028 | 0.012 | 0.013 | **Gatherings (before)** | -1.087 | 0.180 | 0.085 | 0.812* | 0.772* |
| | (-0.508 to 0.112) | (-0.256 to 0.193) | (-0.249 to 0.192) | (-0.269 to 0.293) | (-0.266 to 0.292) | | (-2.391 to 0.217) | (-0.682 to 1.043) | (-0.789 to 0.959) | (0.130 to 1.495) | (0.069 to 1.476) |
| **Gatherings (11-20 days before)** | -0.313* | 0.048 | 0.036 | 0.048 | 0.032 | **Gatherings (0-7 days after)** | 0.785 | 1.364 | 1.078 | 1.357* | 1.288* |
| | (-0.603 to -0.024) | (-0.200 to 0.297) | (-0.219 to 0.291) | (-0.264 to 0.360) | (-0.311 to 0.375) | | (-1.423 to 2.993) | (-0.442 to 3.171) | (-0.604 to 2.760) | (0.129 to 2.586) | (0.074 to 2.501) |
| **Gatherings (20+ days before)** | -0.362** | 0.065 | 0.079 | -0.177 | -0.067 | **Gatherings (8-14 days after)** | -0.831 | -4.833* | -4.925** | -5.307*** | -5.648*** |
| | (-0.634 to -0.089) | (-0.295 to 0.425) | (-0.297 to 0.455) | (-0.669 to 0.314) | (-0.513 to 0.379) | | (-2.311 to 0.649) | (-8.578 to -1.089) | (-8.603 to -1.247) | (-8.203 to -2.410) | (-8.762 to -2.534) |
| **Transport (Not introduced)** | . | . | . | . | . | **Transport (Not introduced)** | . | . | . | . | . |
| **Transport (0-10 days before)** | 0.007 | 0.045 | 0.035 | 0.121 | 0.062 | **Transport (before)** | -0.695 | -0.378 | -0.332 | -0.031 | -0.108 |

| | | | | | | | | | | | |
|---|---|---|---|---|---|---|---|---|---|---|---|
| | (-0.275 to 0.288) | (-0.211 to 0.302) | (-0.222 to 0.293) | (-0.125 to 0.368) | (-0.207 to 0.331) | | (-1.443 to 0.052) | (-1.119 to 0.363) | (-1.039 to 0.376) | (-0.819 to 0.757) | (-0.930 to 0.714) |
| **Transport (11-20 days before)** | -0.261** | -0.031 | -0.049 | 0.193 | 0.040 | **Transport (0-7 days after)** | -0.642 | -0.832 | -0.670 | -1.064 | -1.000 |
| | (-0.426 to -0.097) | (-0.401 to 0.338) | (-0.432 to 0.333) | (-0.226 to 0.613) | (-0.383 to 0.462) | | (-1.414 to 0.130) | (-2.623 to 0.960) | (-2.361 to 1.021) | (-2.894 to 0.766) | (-2.724 to 0.724) |
| **Transport (20+ days before)** | -0.204* | -0.054 | -0.089 | 0.651 | 0.268 | **Transport (8-14 days after)** | 1.713 | 0.845 | 0.734 | 1.731 | 1.500 |
| | (-0.374 to -0.033) | (-0.487 to 0.380) | (-0.530 to 0.352) | (-0.061 to 1.362) | (-0.448 to 0.985) | | (-3.114 to 6.541) | (-2.111 to 3.800) | (-2.232 to 3.699) | (-0.083 to 3.546) | (-0.208 to 3.207) |
| **Stay home (Not introduced)** | . | . | . | . | . | **Stay home (Not introduced)** | . | . | . | . | . |
| **Stay home (0-10 days before)** | -0.132 | -0.138 | -0.154 | -0.200 | -0.253 | **Stay home (before)** | -0.485 | 0.373 | 0.317 | 0.123 | -0.058 |
| | (-0.345 to 0.081) | (-0.441 to 0.166) | (-0.466 to 0.158) | (-0.517 to 0.117) | (-0.616 to 0.111) | | (-1.140 to 0.170) | (-0.560 to 1.306) | (-0.641 to 1.275) | (-0.952 to 1.198) | (-1.017 to 0.901) |
| **Stay home (11-20 days before)** | -0.224 | -0.065 | -0.064 | -0.166 | -0.087 | **Stay home (0-7 days after)** | 0.650 | 0.268 | 0.360 | 0.604 | 0.555 |
| | (-0.480 to 0.032) | (-0.523 to 0.393) | (-0.528 to 0.400) | (-0.543 to 0.211) | (-0.486 to 0.311) | | (-0.896 to 2.195) | (-0.914 to 1.450) | (-0.842 to 1.562) | (-0.872 to 2.079) | (-0.832 to 1.943) |
| **Stay home (20+ days before)** | -0.303** | -0.171 | -0.237 | -0.647* | -0.440 | **Stay home (8-14 days after)** | 0.853 | -0.225 | -0.121 | -0.012 | -0.271 |
| | (-0.500 to -0.105) | (-0.646 to 0.304) | (-0.740 to 0.265) | (-1.214 to -0.079) | (-1.023 to 0.143) | | (-1.165 to 2.871) | (-1.560 to 1.109) | (-1.426 to 1.183) | (-1.030 to 1.007) | (-1.231 to 0.689) |
| **Internal movement (Not introduced)** | . | . | . | . | . | **Internal movement (Not introduced)** | . | . | . | . | . |
| **Internal movement (0-10 days before)** | -0.160 | -0.072 | -0.055 | -0.149 | -0.156 | **Internal movement (before)** | -0.409 | 0.425 | 0.468 | 0.211 | 0.295 |
| | (-0.363 to 0.042) | (-0.343 to 0.198) | (-0.326 to 0.216) | (-0.413 to 0.114) | (-0.432 to 0.119) | | (-0.978 to 0.160) | (-0.353 to 1.202) | (-0.318 to 1.254) | (-0.464 to 0.887) | (-0.389 to 0.980) |

| | | | | | | | | | | |
|---|---|---|---|---|---|---|---|---|---|---|
| **Internal movement (11-20 days before)** | -0.094 | 0.095 | 0.131 | 0.139 | 0.171 | **Internal movement (0-7 days after)** | 0.586 | 1.583 | 1.485 | 1.210 | 1.214 |
| | (-0.356 to 0.168) | (-0.269 to 0.459) | (-0.240 to 0.502) | (-0.127 to 0.404) | (-0.121 to 0.463) | | (-1.035 to 2.207) | (-0.173 to 3.339) | (-0.211 to 3.182) | (-0.374 to 2.795) | (-0.534 to 2.962) |
| **Internal movement (20+ days before)** | -0.322*** | 0.114 | 0.188 | 0.188 | 0.320 | **Internal movement (8-14 days after)** | 1.202 | 0.150 | 0.286 | -0.030 | 0.106 |
| | (-0.506 to -0.139) | (-0.222 to 0.450) | (-0.195 to 0.571) | (-0.266 to 0.642) | (-0.227 to 0.867) | | (-0.384 to 2.789) | (-1.120 to 1.421) | (-0.996 to 1.568) | (-1.064 to 1.005) | (-0.887 to 1.098) |
| **Travel (Not introduced)** | . | . | . | . | . | **Travel (Not introduced)** | . | . | . | . | . |
| **Travel (0-10 days before)** | -0.518 | -0.372 | -0.349 | 0.099 | -0.003 | **Travel (before)** | -3.026* | -1.685 | -1.609 | -0.896 | -1.035 |
| | (-1.265 to 0.229) | (-1.145 to 0.401) | (-1.133 to 0.435) | (-0.575 to 0.773) | (-0.684 to 0.678) | | (-5.479 to -0.573) | (-4.201 to 0.830) | (-4.154 to 0.936) | (-3.255 to 1.463) | (-3.356 to 1.286) |
| **Travel (11-20 days before)** | -0.719* | -0.491 | -0.482 | -0.077 | -0.149 | **Travel (0-7 days after)** | -0.818 | -1.163 | -1.212 | -1.536 | -1.348 |
| | (-1.374 to -0.063) | (-1.124 to 0.142) | (-1.122 to 0.158) | (-0.576 to 0.422) | (-0.687 to 0.389) | | (-4.552 to 2.916) | (-4.024 to 1.697) | (-4.032 to 1.608) | (-3.738 to 0.665) | (-3.496 to 0.800) |
| **Travel (20+ days before)** | -0.824* | -0.801* | -0.781* | -0.360 | -0.453 | **Travel (8-14 days after)** | -0.372 | 1.479 | 1.611 | 1.020 | 1.100 |
| | (-1.464 to -0.185) | (-1.490 to -0.112) | (-1.474 to -0.088) | (-0.970 to 0.251) | (-1.081 to 0.174) | | (-5.424 to 4.680) | (-3.677 to 6.635) | (-3.581 to 6.802) | (-2.104 to 4.145) | (-1.778 to 3.978) |
| **Public information (Not introduced)** | . | . | . | . | . | **Public information (Not introduced)** | . | . | . | . | . |
| **Public information (0-10 days before)** | 0.212 | 0.197 | 0.230 | 0.322 | 0.336 | **Public information (before)** | 0.197 | 1.004 | 1.520 | -0.154 | 0.416 |
| | (-0.349 to 0.774) | (-0.301 to 0.695) | (-0.279 to 0.739) | (-0.256 to 0.899) | (-0.282 to 0.955) | | (-0.674 to 1.067) | (-2.041 to 4.049) | (-1.609 to 4.650) | (-2.750 to 2.442) | (-2.041 to 2.874) |
| **Public information (11-20 days before)** | 0.009 | 0.308 | 0.320 | 0.190 | 0.232 | **Public information** | 0.884 | -1.400 | -0.857 | -1.988 | -1.353 |

| | | | | | | | | | | | |
|---|---|---|---|---|---|---|---|---|---|---|---|
| | | | | | | **(0-7 days after)** | | | | | |
| | (-0.332 to 0.351) | (-0.340 to 0.957) | (-0.333 to 0.973) | (-0.458 to 0.838) | (-0.436 to 0.899) | | (-1.053 to 2.821) | (-5.346 to 2.546) | (-4.775 to 3.061) | (-5.357 to 1.380) | (-4.456 to 1.749) |
| **Public information (20+ days before)** | 0.058 | 0.526 | 0.565 | 0.462 | 0.477 | **Public information (8-14 days after)** | -0.465 | 0.398 | 0.537 | -1.351 | -0.485 |
| | (-0.293 to 0.409) | (-0.134 to 1.186) | (-0.111 to 1.241) | (-0.246 to 1.170) | (-0.225 to 1.179) | | (-1.242 to 0.312) | (-3.805 to 4.602) | (-3.759 to 4.833) | (-3.866 to 1.163) | (-3.441 to 2.471) |
| **Testing (Not introduced)** | . | . | . | . | . | **Testing (Not introduced)** | . | . | . | . | . |
| **Testing (0-10 days before)** | 0.040 | -0.143 | -0.149 | -0.159 | -0.228 | **Testing (before)** | 0.607* | 0.660 | 0.563 | 0.422 | 0.239 |
| | (-0.141 to 0.220) | (-0.433 to 0.146) | (-0.442 to 0.143) | (-0.458 to 0.140) | (-0.588 to 0.131) | | (0.057 to 1.157) | (-0.252 to 1.572) | (-0.353 to 1.478) | (-0.273 to 1.117) | (-0.538 to 1.016) |
| **Testing (11-20 days before)** | 0.401* | 0.083 | 0.075 | 0.091 | 0.037 | **Testing (0-7 days after)** | 0.041 | 1.637 | 1.361 | 1.341 | 1.186 |
| | (0.020 to 0.782) | (-0.337 to 0.503) | (-0.336 to 0.486) | (-0.267 to 0.450) | (-0.354 to 0.428) | | (-0.405 to 0.487) | (-0.695 to 3.969) | (-0.715 to 3.437) | (-0.317 to 2.999) | (-0.483 to 2.856) |
| **Testing (20+ days before)** | 0.190 | 0.014 | -0.005 | 0.022 | -0.029 | **Testing (8-14 days after)** | -0.133 | 2.117 | 2.053 | 1.340* | 1.773 |
| | (-0.001 to 0.382) | (-0.283 to 0.312) | (-0.312 to 0.301) | (-0.263 to 0.307) | (-0.352 to 0.293) | | (-0.448 to 0.183) | (-0.043 to 4.277) | (-0.216 to 4.323) | (0.040 to 2.639) | (-0.218 to 3.765) |
| **Contact tracing (Not introduced)** | . | . | . | . | . | **Contact tracing (Not introduced)** | . | . | . | . | . |
| **Contact tracing (0-10 days before)** | 0.067 | 0.166 | 0.201 | 0.412** | 0.485** | **Contact tracing (before)** | 0.671* | 0.675 | 0.578 | 0.797* | 0.690 |
| | (-0.142 to 0.275) | (-0.123 to 0.456) | (-0.096 to 0.498) | (0.120 to 0.703) | (0.152 to 0.817) | | (0.098 to 1.245) | (-0.047 to 1.396) | (-0.162 to 1.317) | (0.034 to 1.559) | (-0.133 to 1.512) |
| **Contact tracing (11-20 days before)** | 0.414* | 0.560* | 0.568* | 0.526* | 0.550* | **Contact tracing (0-7 days after)** | -0.098 | 0.074 | 0.053 | 0.219 | -0.031 |
| | (0.051 to 0.778) | (0.121 to 0.999) | (0.133 to 1.003) | (0.089 to 0.964) | (0.111 to 0.989) | | (-0.402 to 0.207) | (-1.535 to 1.683) | (-1.528 to 1.633) | (-0.935 to 1.373) | (-1.287 to 1.225) |

| | | | | | | | | | | | |
|---|---|---|---|---|---|---|---|---|---|---|---|
| **Contact tracing (20+ days before)** | 0.153 | 0.351* | 0.295 | 0.221 | 0.184 | **Contact tracing (8-14 days after)** | -0.178 | 2.167 | 2.228* | 1.674 | 2.049* |
| | (-0.075 to 0.381) | (0.036 to 0.667) | (-0.021 to 0.611) | (-0.062 to 0.504) | (-0.085 to 0.453) | | (-0.489 to 0.134) | (-0.009 to 4.342) | (0.086 to 4.370) | (-0.077 to 3.425) | (0.046 to 4.052) |
| **Date of first case** | | | -0.004 | -0.004 | -0.007 | | | | -0.017 | -0.014 | -0.024 |
| | | | (-0.013 to 0.004) | (-0.018 to 0.010) | (-0.024 to 0.010) | | | | (-0.037 to 0.003) | (-0.045 to 0.016) | (-0.064 to 0.016) |
| **Population density (people per sq.km)** | | | | -0.000 | -0.000 | | | | | -0.000 | -0.000 |
| | | | | (-0.000 to 0.000) | (-0.000 to 0.000) | | | | | (-0.001 to 0.000) | (-0.001 to 0.000) |
| **% Population aged 65+** | | | | 0.043* | 0.050* | | | | | 0.076 | 0.075 |
| | | | | (0.009 to 0.077) | (0.012 to 0.089) | | | | | (-0.041 to 0.193) | (-0.049 to 0.199) |
| **% Population male** | | | | 0.001 | 0.002 | | | | | 0.008 | 0.008 |
| | | | | (-0.025 to 0.027) | (-0.028 to 0.031) | | | | | (-0.087 to 0.104) | (-0.093 to 0.108) |
| **Life expectancy at birth (years)** | | | | 0.024* | 0.024* | | | | | 0.024 | 0.034 |
| | | | | (0.003 to 0.046) | (0.002 to 0.046) | | | | | (-0.042 to 0.090) | (-0.033 to 0.100) |
| **Hospital beds (per 1000 people)** | | | | -0.074** | -0.047 | | | | | -0.205** | -0.145* |
| | | | | (-0.122 to -0.027) | (-0.101 to 0.007) | | | | | (-0.339 to -0.071) | (-0.289 to -0.002) |
| **Physicians (per 1000 people)** | | | | -0.037 | -0.098 | | | | | -0.068 | -0.140 |
| | | | | (-0.198 to 0.123) | (-0.275 to 0.079) | | | | | (-0.629 to 0.493) | (-0.755 to 0.474) |
| **GDP PPP (current international $)** | | | | -0.000 | -0.000 | | | | | -0.000 | 0.000 |
| | | | | (-0.000 to 0.000) | (-0.000 to 0.000) | | | | | (-0.000 to 0.000) | (-0.000 to 0.000) |

| | | | | | | | | | | | |
|---|---|---|---|---|---|---|---|---|---|---|---|
| **Manufacturing value added (%GDP)** | | | | -0.014 | -0.018 | | | | | -0.021 | -0.025 |
| | | | | (-0.032 to 0.004) | (-0.038 to 0.001) | | | | | (-0.063 to 0.020) | (-0.068 to 0.018) |
| **Health expenditure (%GDP)** | | | | -0.043 | -0.017 | | | | | 0.121 | 0.167 |
| | | | | (-0.095 to 0.009) | (-0.065 to 0.032) | | | | | (-0.097 to 0.338) | (-0.080 to 0.415) |
| **International tourism, number of arrivals** | | | | 0.000 | 0.000 | | | | | 0.000 | 0.000 |
| | | | | (-0.000 to 0.000) | (-0.000 to 0.000) | | | | | (-0.000 to 0.000) | (-0.000 to 0.000) |
| **Governance (Voice and Accountability)** | | | | -0.049 | -0.047 | | | | | 0.077 | 0.086 |
| | | | | (-0.233 to 0.135) | (-0.236 to 0.142) | | | | | (-0.321 to 0.475) | (-0.375 to 0.547) |
| **East Asia & Pacific** | | | | . | . | | | | | . | . |
| **Europe & Central Asia** | | | | 0.672* | 0.380 | | | | | 2.156** | 1.664* |
| | | | | (0.158 to 1.185) | (-0.186 to 0.947) | | | | | (0.616 to 3.697) | (0.208 to 3.119) |
| **America & Caribbean** | | | | 0.207 | -0.074 | | | | | 1.171 | 0.782 |
| | | | | (-0.103 to 0.517) | (-0.440 to 0.292) | | | | | (-0.014 to 2.356) | (-0.377 to 1.941) |
| **East & North Africa** | | | | 0.169 | 0.088 | | | | | 0.677 | 0.532 |
| | | | | (-0.173 to 0.510) | (-0.296 to 0.471) | | | | | (-0.751 to 2.104) | (-0.904 to 1.967) |
| **North America** | | | | 0.251 | 0.034 | | | | | -3.090 | -4.183* |
| | | | | (-0.589 to 1.090) | (-0.974 to 1.042) | | | | | (-6.981 to 0.800) | (-8.342 to -0.024) |
| **South Asia** | | | | 0.186 | 0.082 | | | | | 1.030 | 0.883 |

| | | | | | | | | | | | |
|---|---|---|---|---|---|---|---|---|---|---|---|
| | | | | (-0.250 to 0.622) | (-0.413 to 0.577) | | | | | (-0.664 to 2.724) | (-0.919 to 2.686) |
| **Sub-Saharan Africa** | | | | 0.548** | 0.340 | | | | | 1.753* | 1.410 |
| | | | | (0.152 to 0.944) | (-0.054 to 0.735) | | | | | (0.225 to 3.281) | (-0.059 to 2.879) |
| **Constant** | | 0.519 | 98.071 | 81.592 | 153.282 | | 0.004 | 372.228 | 309.713 | 523.627 | 0.004 |
| | | (-0.008 to 1.046) | (-85.321 to 281.463) | (-227.398 to 390.582) | (-224.122 to 530.687) | | (-3.895 to 3.903) | (-62.854 to 807.311) | (-364.293 to 983.719) | (-352.086 to 1399.341) | (-3.895 to 3.903) |
| **Observations** | 3250 | 3250 | 3250 | 3250 | 3250 | 3150 | 3150 | 3150 | 3150 | 3150 | 3150 |
| **R-squared** | | 0.129 | 0.132 | 0.222 | 0.326 | | 0.323 | 0.329 | 0.479 | 0.514 | 0.323 |
| **Adjusted R-squared** | | 0.120 | 0.123 | 0.209 | 0.305 | | 0.316 | 0.321 | 0.471 | 0.498 | 0.316 |

**p-values in parentheses =** * p<0.05 ** p<0.01 *** p<0.001; # includes time, day of week and week of year fixed effects

#### Regression results – Negative binomial (count model) specifications

*Regression results, mean policy strictness (combination of timing and strictness)*

| | 24 Days (Policies implemented before first death) | | | | | 38 Days (Policies implemented within 14 days after first death) | | | | |
|---|---|---|---|---|---|---|---|---|---|---|
| **Average deaths per day over time period, per million** | **(1)** | **(2)** | **(3)** | **(4)** | **(5) #** | **(1)** | **(2)** | **(3)** | **(4)** | **(5) #** |
| **School** | 0.382*** | 0.272*** | 0.276*** | 0.474** | 0.675 | 0.201*** | 0.182*** | 0.267** | 0.285*** | 0.322** |
| | (0.248 to 0.588) | (0.150 to 0.491) | (0.150 to 0.509) | (0.298 to 0.756) | (0.411 to 1.110) | (0.119 to 0.339) | (0.074 to 0.445) | (0.113 to 0.627) | (0.146 to 0.553) | (0.161 to 0.644) |
| **Work** | 0.426** | 1.515 | 1.525 | 0.769 | 0.756 | 0.421** | 1.987* | 1.871 | 1.132 | 1.519 |
| | (0.229 to 0.794) | (0.727 to 3.158) | (0.741 to 3.136) | (0.381 to 1.552) | (0.374 to 1.531) | (0.227 to 0.781) | (1.000 to 3.949) | (0.966 to 3.622) | (0.567 to 2.258) | (0.682 to 3.384) |
| **Events** | 0.325*** | 1.137 | 1.141 | 0.828 | 0.613 | 0.171*** | 1.240 | 1.239 | 1.223 | 1.282 |
| | (0.178 to 0.596) | (0.444 to 2.909) | (0.442 to 2.944) | (0.428 to 1.605) | (0.305 to 1.231) | (0.083 to 0.352) | (0.389 to 3.959) | (0.402 to 3.817) | (0.540 to 2.772) | (0.499 to 3.295) |
| **Gatherings** | 0.524** | 1.459 | 1.460 | 1.585* | 1.866* | 0.423*** | 1.167 | 1.170 | 1.120 | 1.238 |
| | (0.342 to 0.803) | (0.770 to 2.766) | (0.769 to 2.773) | (1.036 to 2.424) | (1.140 to 3.052) | (0.286 to 0.625) | (0.675 to 2.017) | (0.667 to 2.053) | (0.709 to 1.769) | (0.791 to 1.938) |
| **Transport** | 0.330** | 1.401 | 1.392 | 1.469 | 1.199 | 0.217** | 1.165 | 1.235 | 0.954 | 0.897 |
| | (0.156 to 0.699) | (0.367 to 5.349) | (0.364 to 5.316) | (0.525 to 4.110) | (0.427 to 3.360) | (0.084 to 0.559) | (0.365 to 3.723) | (0.469 to 3.251) | (0.347 to 2.619) | (0.317 to 2.535) |
| **Stay home** | 0.326** | 0.932 | 0.929 | 0.995 | 1.049 | 0.295*** | 1.308 | 1.401 | 1.504 | 1.348 |
| | (0.159 to 0.670) | (0.299 to 2.904) | (0.298 to 2.897) | (0.411 to 2.408) | (0.437 to 2.515) | (0.164 to 0.529) | (0.461 to 3.706) | (0.529 to 3.710) | (0.773 to 2.928) | (0.711 to 2.556) |
| **Internal movement** | 0.366 | 1.323 | 1.324 | 1.691 | 1.957 | 0.285* | 0.669 | 0.623 | 1.797 | 2.491 |
| | (0.120 to 1.117) | (0.226 to 7.739) | (0.232 to 7.549) | (0.786 to 3.640) | (0.842 to 4.550) | (0.108 to 0.754) | (0.198 to 2.266) | (0.219 to 1.775) | (0.651 to 4.955) | (0.745 to 8.326) |
| **Travel** | 0.563*** | 0.563*** | 0.563*** | 0.902 | 1.068 | 0.401*** | 0.474*** | 0.455*** | 0.749* | 0.849 |
| | (0.431 to 0.736) | (0.418 to 0.759) | (0.418 to 0.758) | (0.736 to 1.106) | (0.854 to 1.336) | (0.267 to 0.601) | (0.321 to 0.701) | (0.311 to 0.666) | (0.563 to 0.995) | (0.676 to 1.066) |

| | | | | | | | | | | |
|---|---|---|---|---|---|---|---|---|---|---|
| **Public information** | 1.195 | 1.622* | 1.626* | 1.150 | 0.934 | 1.223 | 2.058* | 2.060* | 1.252 | 1.226 |
| | (0.677 to 2.107) | (1.038 to 2.535) | (1.031 to 2.563) | (0.794 to 1.666) | (0.680 to 1.282) | (0.578 to 2.590) | (1.152 to 3.678) | (1.172 to 3.620) | (0.747 to 2.100) | (0.776 to 1.936) |
| **Testing** | 1.341 | 1.318 | 1.313 | 1.276 | 1.154 | 1.163 | 0.996 | 0.985 | 0.972 | 1.079 |
| | (0.899 to 2.000) | (0.764 to 2.273) | (0.751 to 2.296) | (0.876 to 1.860) | (0.781 to 1.705) | (0.640 to 2.114) | (0.553 to 1.796) | (0.524 to 1.853) | (0.614 to 1.538) | (0.674 to 1.726) |
| **Contact tracing** | 1.543* | 1.493* | 1.467* | 1.312 | 1.223 | 1.876* | 1.764* | 1.427 | 1.288 | 1.298 |
| | (1.015 to 2.345) | (1.081 to 2.062) | (1.031 to 2.086) | (0.984 to 1.749) | (0.907 to 1.649) | (1.107 to 3.180) | (1.119 to 2.780) | (0.908 to 2.241) | (0.906 to 1.831) | (0.938 to 1.798) |
| **Date of first case** | | | 0.999 | 0.999 | 0.990 | | | 0.981 | 0.983 | 0.982 |
| | | | (0.981 to 1.016) | (0.976 to 1.023) | (0.963 to 1.018) | | | (0.958 to 1.004) | (0.959 to 1.009) | (0.961 to 1.004) |
| **Population density (people per sq.km)** | | | | 1.000 | 1.000 | | | | 1.000 | 1.000 |
| | | | | (1.000 to 1.000) | (1.000 to 1.000) | | | | (1.000 to 1.000) | (1.000 to 1.000) |
| **% Population aged 65+** | | | | 0.997 | 0.989 | | | | 0.994 | 0.990 |
| | | | | (0.932 to 1.068) | (0.917 to 1.066) | | | | (0.906 to 1.090) | (0.910 to 1.076) |
| **% Population male** | | | | 1.012 | 0.988 | | | | 1.015 | 1.014 |
| | | | | (0.963 to 1.064) | (0.940 to 1.038) | | | | (0.954 to 1.080) | (0.947 to 1.086) |
| **Life expectancy at birth (years)** | | | | 1.094* | 1.158*** | | | | 1.123 | 1.123 |
| | | | | (1.012 to 1.182) | (1.061 to 1.264) | | | | (1.000 to 1.262) | (0.986 to 1.278) |
| **Hospital beds (per 1000 people)** | | | | 0.981 | 1.046 | | | | 0.931 | 0.926 |

| | | | | | | | | | | |
|---|---|---|---|---|---|---|---|---|---|---|
| | | | | (0.855 to 1.124) | (0.902 to 1.213) | | | | (0.803 to 1.078) | (0.806 to 1.063) |
| **Physicians (per 1000 people)** | | | | 0.949 | 0.809 | | | | 0.954 | 0.971 |
| | | | | (0.721 to 1.248) | (0.604 to 1.084) | | | | (0.728 to 1.251) | (0.761 to 1.237) |
| **GDP PPP (current international $)** | | | | 1.000 | 1.000 | | | | 1.000 | 1.000 |
| | | | | (1.000 to 1.000) | (1.000 to 1.000) | | | | (1.000 to 1.000) | (1.000 to 1.000) |
| **Manufacturing value added (%GDP)** | | | | 0.975* | 0.971* | | | | 1.011 | 1.013 |
| | | | | (0.953 to 0.997) | (0.949 to 0.993) | | | | (0.980 to 1.044) | (0.981 to 1.046) |
| **Health expenditure (%GDP)** | | | | 1.011 | 1.024 | | | | 1.007 | 1.008 |
| | | | | (0.900 to 1.137) | (0.919 to 1.142) | | | | (0.893 to 1.135) | (0.898 to 1.132) |
| **International tourism, number of arrivals** | | | | 1.000 | 1.000** | | | | 1.000 | 1.000 |
| | | | | (1.000 to 1.000) | (1.000 to 1.000) | | | | (1.000 to 1.000) | (1.000 to 1.000) |
| **Governance (Voice and Accountability)** | | | | 1.304 | 1.278 | | | | 1.074 | 0.958 |
| | | | | (0.884 to 1.924) | (0.786 to 2.075) | | | | (0.676 to 1.706) | (0.582 to 1.575) |
| **East Asia & Pacific** | | | | . | . | | | | . | . |
| **Europe & Central Asia** | | | | 26.200*** | 25.756*** | | | | 105.616*** | 114.795*** |

| | | | | | | | | | | |
|---|---|---|---|---|---|---|---|---|---|---|
| | | | | (7.563 to 90.764) | (6.397 to 103.694) | | | | (25.290 to 441.071) | (27.617 to 477.166) |
| **America & Caribbean** | | | | 13.066*** | 15.203*** | | | | 36.711*** | 42.479*** |
| | | | | (3.507 to 48.675) | (3.436 to 67.272) | | | | (7.627 to 176.701) | (9.674 to 186.529) |
| **East & North Africa** | | | | 10.525*** | 9.739*** | | | | 32.358*** | 27.944*** |
| | | | | (3.058 to 36.228) | (2.581 to 36.749) | | | | (7.554 to 138.605) | (7.209 to 108.311) |
| **North America** | | | | 4.983* | 2.369 | | | | 36.657*** | 40.018*** |
| | | | | (1.036 to 23.974) | (0.486 to 11.552) | | | | (7.460 to 180.135) | (7.081 to 226.163) |
| **South Asia** | | | | 1.033 | 0.840 | | | | 2.199 | 2.189 |
| | | | | (0.291 to 3.663) | (0.166 to 4.258) | | | | (0.372 to 13.018) | (0.339 to 14.129) |
| **Sub-Saharan Africa** | | | | 7.933* | 10.594* | | | | 20.613** | 36.921*** |
| | | | | (1.490 to 42.222) | (1.652 to 67.950) | | | | (2.498 to 170.100) | (4.395 to 310.164) |
| **Lnalpha** | | 1.133 | 1.134 | 0.591* | 0.069** | | 1.137 | 1.152 | 0.356*** | 0.148*** |
| | | (0.774 to 1.657) | (0.774 to 1.662) | (0.375 to 0.934) | (0.012 to 0.409) | | (0.709 to 1.824) | (0.723 to 1.835) | (0.226 to 0.560) | (0.064 to 0.343) |
| **Observations** | 3250 | 3250 | 3250 | 3250 | 3250 | 3150 | 3150 | 3150 | 3150 | 3150 |
| **Pseudo R-squared** | | 0.122 | 0.122 | 0.255 | 0.374 | | 0.193 | 0.200 | 0.360 | 0.399 |

**p-values in parentheses =** * p<0.05 ** p<0.01 *** p<0.001; # includes time, day of week and week of year fixed effects

*Regression results, policy strictness. Baseline is policy not introduced within policy analysis period*

| | **24 Days (Policies implemented before first death)** | | | | | **38 Days (Policies implemented within 14 days after first death)** | | | | |
|---|---|---|---|---|---|---|---|---|---|---|
| **Incidence Rate Ratio** | **(1)** | **(2)** | **(3)** | **(4)** | **(5) #~** | **(1)** | **(2)** | **(3)** | **(4)** | **(5) #** |
| **0bn.School** | . | . | . | . | . | . | . | . | . | . |
| **1.School** | 0.132*** | 0.244* | 0.240* | 8.235** | 2.793 | 6.478** | 489.478*** | 3747.842*** | 10.247 | 5.854 |
| | (0.074 to 0.233) | (0.069 to 0.863) | (0.068 to 0.853) | (1.908 to 35.546) | (0.527 to 14.799) | (1.693 to 24.778) | (39.697 to 6035.509) | (140.712 to 99823.470) | (0.761 to 138.057) | (0.304 to 112.667) |
| **2.School** | 0.343* | 0.522 | 0.542 | 0.465 | 0.322* | 4.686 | 13.002** | 44.631*** | 1.372 | 1.091 |
| | (0.121 to 0.973) | (0.192 to 1.420) | (0.189 to 1.554) | (0.184 to 1.174) | (0.113 to 0.914) | (0.615 to 35.676) | (1.932 to 87.489) | (7.144 to 278.805) | (0.347 to 5.422) | (0.249 to 4.784) |
| **3.School** | 0.302*** | 0.663 | 0.694 | 0.680 | 0.536 | 0.773 | 2.369 | 9.090** | 0.953 | 0.848 |
| | (0.157 to 0.583) | (0.303 to 1.452) | (0.301 to 1.602) | (0.388 to 1.189) | (0.285 to 1.007) | (0.186 to 3.214) | (0.708 to 7.923) | (2.148 to 38.466) | (0.304 to 2.991) | (0.258 to 2.782) |
| **0bn.Work** | . | . | . | . | . | . | . | . | . | . |
| **1.Work** | 1.214 | 3.272* | 3.339* | 0.933 | 0.747 | 2.177 | 0.795 | 0.839 | 0.738 | 0.672 |
| | (0.441 to 3.340) | (1.167 to 9.172) | (1.199 to 9.299) | (0.476 to 1.830) | (0.392 to 1.424) | (0.541 to 8.762) | (0.206 to 3.072) | (0.227 to 3.094) | (0.278 to 1.960) | (0.237 to 1.908) |
| **2.Work** | 0.675 | 3.243* | 3.250* | 1.084 | 0.812 | 2.108 | 1.533 | 2.033 | 0.412* | 0.346* |
| | (0.308 to 1.481) | (1.172 to 8.971) | (1.175 to 8.991) | (0.635 to 1.851) | (0.478 to 1.380) | (0.526 to 8.451) | (0.544 to 4.325) | (0.880 to 4.697) | (0.188 to 0.903) | (0.152 to 0.785) |
| **3.Work** | 0.552 | 4.960** | 4.995** | 1.618 | 1.162 | 1.706 | 1.700 | 2.425 | 1.452 | 1.275 |
| | (0.273 to 1.119) | (1.484 to 16.576) | (1.504 to 16.590) | (0.795 to 3.293) | (0.577 to 2.341) | (0.475 to 6.134) | (0.587 to 4.919) | (0.956 to 6.154) | (0.657 to 3.211) | (0.549 to 2.965) |
| **0bn.Events** | . | . | . | . | . | . | . | . | . | . |
| **1.Events** | 1.347 | 0.922 | 0.930 | 1.155 | 0.893 | 0.677 | 0.481 | 1.159 | 0.527 | 0.572 |
| | (0.339 to 5.344) | (0.303 to 2.805) | (0.305 to 2.831) | (0.532 to 2.511) | (0.407 to 1.960) | (0.110 to 4.158) | (0.056 to 4.129) | (0.126 to 10.643) | (0.136 to 2.043) | (0.134 to 2.445) |
| **2.Events** | 0.446* | 0.535 | 0.539 | 1.042 | 0.823 | 1.239 | 1.290 | 2.023 | 0.563 | 0.615 |
| | (0.216 to 0.920) | (0.242 to 1.183) | (0.246 to 1.179) | (0.568 to 1.914) | (0.468 to 1.447) | (0.386 to 3.975) | (0.292 to 5.700) | (0.340 to 12.022) | (0.109 to 2.900) | (0.121 to 3.130) |
| **0bn.Gatherings** | . | . | . | . | . | . | . | . | . | . |

| | | | | | | | | | | |
|---|---|---|---|---|---|---|---|---|---|---|
| **1.Gatherings** | 0.775 | 1.327 | 1.352 | 0.556 | 0.888 | 0.257* | 0.561 | 1.396 | 0.488 | 0.375 |
| | (0.196 to 3.064) | (0.525 to 3.358) | (0.532 to 3.433) | (0.275 to 1.125) | (0.446 to 1.767) | (0.072 to 0.918) | (0.097 to 3.239) | (0.388 to 5.023) | (0.060 to 4.003) | (0.055 to 2.559) |
| **2.Gatherings** | 0.636 | 0.994 | 0.975 | 0.800 | 0.740 | 1.017 | 0.028*** | 0.011*** | 0.084*** | 0.136* |
| | (0.271 to 1.493) | (0.412 to 2.399) | (0.388 to 2.448) | (0.313 to 2.046) | (0.232 to 2.359) | (0.205 to 5.036) | (0.007 to 0.121) | (0.003 to 0.044) | (0.021 to 0.341) | (0.025 to 0.727) |
| **3.Gatherings** | 0.310** | 0.532 | 0.542 | 0.966 | 1.226 | 0.284 | 0.077*** | 0.205** | 0.561 | 0.612 |
| | (0.132 to 0.729) | (0.253 to 1.119) | (0.249 to 1.177) | (0.473 to 1.970) | (0.569 to 2.640) | (0.078 to 1.030) | (0.022 to 0.263) | (0.065 to 0.650) | (0.272 to 1.157) | (0.315 to 1.187) |
| **4.Gatherings** | 0.566 | 1.179 | 1.175 | 1.339 | 1.355 | 0.633 | 0.192** | 0.273** | 0.294** | 0.313** |
| | (0.249 to 1.290) | (0.461 to 3.019) | (0.463 to 2.986) | (0.619 to 2.899) | (0.630 to 2.915) | (0.202 to 1.982) | (0.065 to 0.569) | (0.106 to 0.703) | (0.133 to 0.649) | (0.141 to 0.694) |
| **0bn.Transport** | . | . | . | . | . | . | . | . | . | . |
| **1.Transport** | 1.068 | 0.933 | 0.930 | 1.101 | 1.263 | 0.856 | 0.921 | 1.030 | 1.289 | 1.258 |
| | (0.430 to 2.654) | (0.446 to 1.949) | (0.449 to 1.924) | (0.662 to 1.832) | (0.768 to 2.077) | (0.290 to 2.529) | (0.426 to 1.992) | (0.509 to 2.087) | (0.781 to 2.128) | (0.770 to 2.055) |
| **2.Transport** | 0.520 | 0.723 | 0.721 | 0.729 | 0.728 | 0.258*** | 0.873 | 0.973 | 0.998 | 0.986 |
| | (0.261 to 1.034) | (0.291 to 1.797) | (0.288 to 1.804) | (0.365 to 1.454) | (0.360 to 1.473) | (0.119 to 0.560) | (0.385 to 1.979) | (0.468 to 2.023) | (0.607 to 1.641) | (0.637 to 1.527) |
| **0bn.Stay home** | . | . | . | . | . | . | . | . | . | . |
| **1.Stay home** | 0.852 | 1.819 | 1.778 | 1.211 | 1.003 | 1.059 | 2.936 | 2.472 | 2.186* | 1.965* |
| | (0.418 to 1.735) | (0.888 to 3.727) | (0.846 to 3.740) | (0.633 to 2.317) | (0.410 to 2.454) | (0.386 to 2.903) | (0.960 to 8.982) | (0.897 to 6.817) | (1.123 to 4.253) | (1.049 to 3.682) |
| **2.Stay home** | 0.373** | 0.743 | 0.725 | 0.633 | 0.593 | 1.363 | 1.937 | 1.676 | 1.877 | 1.753 |
| | (0.185 to 0.754) | (0.260 to 2.120) | (0.247 to 2.131) | (0.297 to 1.347) | (0.271 to 1.297) | (0.454 to 4.087) | (0.651 to 5.765) | (0.639 to 4.396) | (0.947 to 3.719) | (0.930 to 3.304) |
| **3.Stay home** | 0.493 | 0.711 | 0.696 | 0.724 | 0.813 | 0.208** | 1.072 | 0.839 | 2.424 | 2.376 |
| | (0.200 to 1.217) | (0.174 to 2.894) | (0.170 to 2.854) | (0.257 to 2.037) | (0.291 to 2.273) | (0.072 to 0.602) | (0.229 to 5.018) | (0.177 to 3.982) | (0.845 to 6.954) | (0.907 to 6.221) |
| **0bn.Internal movement** | . | . | . | . | . | . | . | . | . | . |
| **1.Internal movement** | 0.687 | 1.235 | 1.239 | 0.894 | 0.952 | 2.062 | 3.005** | 2.325* | 1.314 | 1.241 |

| | | | | | | | | | | |
|---|---|---|---|---|---|---|---|---|---|---|
| | (0.323 to 1.460) | (0.615 to 2.480) | (0.624 to 2.461) | (0.523 to 1.528) | (0.548 to 1.654) | (0.725 to 5.864) | (1.445 to 6.253) | (1.219 to 4.434) | (0.888 to 1.944) | (0.859 to 1.794) |
| **2.Internal movement** | 0.421* | 0.975 | 0.988 | 1.189 | 1.362 | 0.805 | 0.873 | 1.023 | 1.171 | 1.164 |
| | (0.211 to 0.841) | (0.196 to 4.858) | (0.198 to 4.942) | (0.506 to 2.793) | (0.554 to 3.349) | (0.243 to 2.668) | (0.450 to 1.692) | (0.534 to 1.962) | (0.763 to 1.798) | (0.762 to 1.779) |
| **0bn.Travel** | . | . | . | . | . | . | . | . | . | . |
| **1.Travel** | 0.528 | 0.381 | 0.382* | 0.788 | 0.877 | 0.007*** | 0.037** | 0.695 | 0.018*** | 0.021*** |
| | (0.134 to 2.080) | (0.144 to 1.003) | (0.147 to 0.997) | (0.312 to 1.986) | (0.343 to 2.240) | (0.004 to 0.013) | (0.003 to 0.447) | (0.020 to 23.837) | (0.002 to 0.132) | (0.004 to 0.104) |
| **2.Travel** | 0.262* | 0.245** | 0.247** | 0.401 | 0.856 | 0.045*** | 0.069** | 0.105* | 0.025 | 0.058 |
| | (0.084 to 0.817) | (0.096 to 0.624) | (0.097 to 0.631) | (0.122 to 1.312) | (0.320 to 2.295) | (0.012 to 0.164) | (0.010 to 0.482) | (0.018 to 0.597) | (0.000 to 3.099) | (0.001 to 4.054) |
| **3.Travel** | 0.282** | 0.236** | 0.235** | 0.768 | 1.443 | 0.322* | 0.280** | 0.217*** | 0.470* | 0.477* |
| | (0.120 to 0.659) | (0.086 to 0.647) | (0.086 to 0.642) | (0.400 to 1.471) | (0.841 to 2.474) | (0.132 to 0.784) | (0.114 to 0.685) | (0.090 to 0.521) | (0.223 to 0.989) | (0.229 to 0.995) |
| **4.Travel** | 0.164*** | 0.139*** | 0.140*** | 0.671 | 1.573 | 0.187** | 0.106*** | 0.083*** | 0.581 | 0.657 |
| | (0.078 to 0.344) | (0.053 to 0.363) | (0.053 to 0.370) | (0.307 to 1.463) | (0.760 to 3.254) | (0.065 to 0.535) | (0.040 to 0.286) | (0.029 to 0.233) | (0.229 to 1.472) | (0.270 to 1.602) |
| **0bn. Public information** | . | . | . | . | . | . | . | . | . | . |
| **1.Public information** | 0.390 | 1.213 | 1.253 | 1.049 | 0.574 | 0.638 | 3.550 | 1.209 | 2.622 | 2.237 |
| | (0.102 to 1.488) | (0.259 to 5.674) | (0.252 to 6.233) | (0.368 to 2.994) | (0.162 to 2.033) | (0.157 to 2.598) | (0.383 to 32.876) | (0.094 to 15.534) | (0.724 to 9.493) | (0.718 to 6.977) |
| **2.Public information** | 1.306 | 2.253 | 2.254 | 1.120 | 0.737 | 1.349 | 3.820 | 1.464 | 0.325* | 0.269* |
| | (0.410 to 4.157) | (0.727 to 6.977) | (0.709 to 7.168) | (0.644 to 1.947) | (0.437 to 1.244) | (0.414 to 4.401) | (0.878 to 16.626) | (0.272 to 7.868) | (0.121 to 0.874) | (0.095 to 0.765) |
| **0bn.Testing** | . | . | . | . | . | . | . | . | . | . |
| **1.Testing** | 2.207* | 1.265 | 1.261 | 0.797 | 0.788 | 2.125 | 1.017 | 1.305 | 0.831 | 0.823 |
| | (1.091 to 4.466) | (0.450 to 3.557) | (0.448 to 3.545) | (0.359 to 1.769) | (0.339 to 1.832) | (0.826 to 5.469) | (0.212 to 4.880) | (0.278 to 6.128) | (0.399 to 1.729) | (0.441 to 1.535) |
| **2.Testing** | 2.253* | 1.145 | 1.163 | 1.024 | 1.381 | 2.228 | 1.098 | 1.376 | 0.918 | 0.924 |

| | | | | | | | | | | |
|---|---|---|---|---|---|---|---|---|---|---|
| | (1.021 to 4.969) | (0.438 to 2.992) | (0.448 to 3.018) | (0.463 to 2.263) | (0.570 to 3.349) | (0.942 to 5.265) | (0.284 to 4.246) | (0.380 to 4.983) | (0.481 to 1.751) | (0.526 to 1.623) |
| **3.Testing** | 7.395** | 5.181* | 5.054* | 3.188* | 3.280* | 5.862* | 3.944* | 2.395 | 2.242* | 2.686** |
| | (1.697 to 32.219) | (1.106 to 24.275) | (1.056 to 24.191) | (1.083 to 9.386) | (1.096 to 9.819) | (1.234 to 27.840) | (1.069 to 14.546) | (0.655 to 8.764) | (1.166 to 4.314) | (1.441 to 5.007) |
| **0bn.Contact tracing** | . | . | . | . | . | . | . | . | . | . |
| **1.Contact tracing** | 1.347 | 1.398 | 1.401 | 1.843 | 1.446 | 1.415 | 2.260 | 1.886 | 1.832* | 2.000* |
| | (0.691 to 2.627) | (0.583 to 3.352) | (0.585 to 3.356) | (0.864 to 3.931) | (0.619 to 3.378) | (0.703 to 2.845) | (0.501 to 10.198) | (0.567 to 6.276) | (1.012 to 3.316) | (1.144 to 3.498) |
| **2.Contact tracing** | 2.902** | 1.954 | 1.942 | 2.055* | 1.632 | 3.466** | 2.614 | 1.446 | 1.158 | 1.278 |
| | (1.419 to 5.936) | (0.773 to 4.936) | (0.761 to 4.955) | (1.055 to 4.002) | (0.828 to 3.215) | (1.567 to 7.666) | (0.597 to 11.455) | (0.498 to 4.201) | (0.682 to 1.964) | (0.731 to 2.235) |
| **Date of first case** | | | 0.998 | 1.016 | 1.003 | | | 0.948*** | 0.967*** | 0.973** |
| | | | (0.980 to 1.015) | (0.997 to 1.035) | (0.984 to 1.022) | | | (0.929 to 0.968) | (0.948 to 0.986) | (0.954 to 0.992) |
| **Population density (people per sq.km)** | | | | 1.000 | 1.000 | | | | 1.000 | 1.000 |
| | | | | (1.000 to 1.000) | (1.000 to 1.000) | | | | (1.000 to 1.000) | (1.000 to 1.000) |
| **% Population aged 65+** | | | | 1.089 | 1.094 | | | | 1.038 | 1.062 |
| | | | | (0.995 to 1.192) | (0.994 to 1.205) | | | | (0.962 to 1.119) | (0.993 to 1.137) |
| **% Population male** | | | | 1.008 | 1.010 | | | | 1.101** | 1.111** |
| | | | | (0.950 to 1.069) | (0.940 to 1.086) | | | | (1.025 to 1.183) | (1.035 to 1.194) |
| **Life expectancy at birth (years)** | | | | 1.105* | 1.138* | | | | 1.058 | 1.051 |
| | | | | (1.015 to 1.203) | (1.030 to 1.258) | | | | (0.953 to 1.174) | (0.956 to 1.155) |
| **Hospital beds (per 1000 people)** | | | | 0.909 | 0.952 | | | | 0.876** | 0.861*** |
| | | | | (0.811 to 1.018) | (0.852 to 1.063) | | | | (0.797 to 0.964) | (0.790 to 0.939) |

| | | | | | | | | | | |
|---|---|---|---|---|---|---|---|---|---|---|
| **Physicians (per 1000 people)** | | | | 0.994 | 0.929 | | | | 0.761* | 0.779* |
| | | | | (0.784 to 1.259) | (0.760 to 1.136) | | | | (0.587 to 0.986) | (0.616 to 0.984) |
| **GDP PPP (current international $)** | | | | 1.000 | 1.000 | | | | 1.000 | 1.000 |
| | | | | (1.000 to 1.000) | (1.000 to 1.000) | | | | (1.000 to 1.000) | (1.000 to 1.000) |
| **Manufacturing value added (%GDP)** | | | | 0.982 | 0.970* | | | | 0.998 | 0.998 |
| | | | | (0.962 to 1.003) | (0.946 to 0.994) | | | | (0.963 to 1.034) | (0.965 to 1.033) |
| **Health expenditure (%GDP)** | | | | 1.009 | 0.987 | | | | 1.048 | 1.025 |
| | | | | (0.894 to 1.139) | (0.886 to 1.099) | | | | (0.935 to 1.174) | (0.912 to 1.153) |
| **International tourism, number of arrivals** | | | | 1.000 | 1.000*** | | | | 1.000 | 1.000 |
| | | | | (1.000 to 1.000) | (1.000 to 1.000) | | | | (1.000 to 1.000) | (1.000 to 1.000) |
| **Governance (Voice and Accountability)** | | | | 1.087 | 1.130 | | | | 1.795** | 1.638** |
| | | | | (0.677 to 1.745) | (0.702 to 1.820) | | | | (1.243 to 2.590) | (1.189 to 2.257) |
| **East Asia & Pacific** | | | | . | . | | | | . | . |
| **Europe & Central Asia** | | | | 12.100*** | 9.609*** | | | | 58.793*** | 49.919*** |
| | | | | (4.002 to 36.584) | (3.203 to 28.824) | | | | (11.656 to 296.542) | (9.209 to 270.592) |
| **America & Caribbean** | | | | 7.944*** | 7.874*** | | | | 11.864*** | 10.581** |
| | | | | (2.316 to 27.257) | (2.399 to 25.841) | | | | (3.020 to 46.606) | (2.499 to 44.812) |
| **East & North Africa** | | | | 8.780*** | 8.233*** | | | | 7.541* | 6.969* |

| | | | | | | | | | | |
|---|---|---|---|---|---|---|---|---|---|---|
| | | | | (2.733 to 28.212) | (2.441 to 27.771) | | | | (1.426 to 39.888) | (1.310 to 37.067) |
| **North America** | | | | 2.705 | 0.729 | | | | 0.595 | 0.430 |
| | | | | (0.460 to 15.925) | (0.117 to 4.549) | | | | (0.099 to 3.586) | (0.071 to 2.604) |
| **South Asia** | | | | 1.204 | 0.958 | | | | 0.912 | 0.903 |
| | | | | (0.311 to 4.654) | (0.227 to 4.041) | | | | (0.132 to 6.319) | (0.138 to 5.911) |
| **Sub-Saharan Africa** | | | | 7.317** | 6.633** | | | | 2.501 | 2.539 |
| | | | | (1.878 to 28.504) | (1.587 to 27.728) | | | | (0.505 to 12.391) | (0.479 to 13.466) |
| **Lnalpha** | | 0.940 | 0.941 | 0.482** | 0.000*** | | 0.935 | 0.633 | 0.134*** | 0.007* |
| | | (0.608 to 1.454) | (0.608 to 1.455) | (0.294 to 0.790) | (0.000 to 0.000) | | (0.594 to 1.472) | (0.359 to 1.113) | (0.076 to 0.235) | (0.000 to 0.637) |
| **Observations** | 3250 | 3250 | 3250 | 3250 | 3250 | 3150 | 3150 | 3150 | 3150 | 3150 |
| **Pseudo R-squared** | . | 0.148 | 0.148 | 0.272 | 0.405 | . | 0.196 | 0.245 | 0.413 | 0.459 |

**p-values in parentheses =** * p<0.05 ** p<0.01 *** p<0.001; # includes time, day of week and week of year fixed effects; ~ model does not converge

*Regression results, policy timing. Baseline is policy not introduced within policy analysis period.*

| | 24 Days (Policies implemented before first death) | | | | | | 38 Days (Policies implemented within 14 days after first death) | | | | |
|---|---|---|---|---|---|---|---|---|---|---|---|
| **Incidence Rate Ratio** | **(1)** | **(2)** | **(3)** | **(4)** | **(5) #~** | | **(1)** | **(2)** | **(3)** | **(4)** | **(5) #** |
| **School (Not introduced)** | . | . | . | . | . | **School (Not introduced)** | . | . | . | . | . |
| **School (0-10 days before)** | 0.450* | 0.581 | 0.729 | 0.562 | 0.591 | **School (before)** | 0.336 | 0.376 | 1.580 | 0.765 | 0.586 |
| | (0.226 to 0.898) | (0.304 to 1.109) | (0.372 to 1.430) | (0.302 to 1.045) | (0.275 to 1.270) | | (0.084 to 1.341) | (0.078 to 1.823) | (0.267 to 9.348) | (0.138 to 4.243) | (0.106 to 3.247) |
| **School (11-20 days before)** | 0.193*** | 0.220** | 0.294** | 0.331** | 0.431 | **School (0-7 days after)** | 3.171 | 2.000 | 7.162* | 1.659 | 1.104 |
| | (0.093 to 0.398) | (0.085 to 0.569) | (0.119 to 0.727) | (0.160 to 0.684) | (0.175 to 1.060) | | (0.678 to 14.821) | (0.468 to 8.543) | (1.471 to 34.876) | (0.391 to 7.041) | (0.205 to 5.945) |
| **School (20+ days before)** | 0.131*** | 0.275 | 0.460 | 0.601 | 0.800 | **School (8-14 days after)** | 4.172 | 12.080*** | 12.453*** | 3.928** | 5.873** |
| | (0.062 to 0.278) | (0.075 to 1.014) | (0.116 to 1.821) | (0.152 to 2.369) | (0.152 to 4.206) | | (0.806 to 21.584) | (2.992 to 48.771) | (3.349 to 46.304) | (1.454 to 10.609) | (1.887 to 18.283) |
| **Work (Not introduced)** | . | . | . | . | . | **Work (Not introduced)** | . | . | . | . | . |
| **Work (0-10 days before)** | 0.995 | 1.950* | 2.107* | 1.534* | 1.450 | **Work (before)** | 0.813 | 1.210 | 1.494 | 0.548 | 0.510 |
| | (0.496 to 1.995) | (1.055 to 3.606) | (1.142 to 3.890) | (1.005 to 2.341) | (0.873 to 2.410) | | (0.266 to 2.484) | (0.419 to 3.493) | (0.500 to 4.458) | (0.259 to 1.162) | (0.259 to 1.005) |
| **Work (11-20 days before)** | 0.211*** | 0.490 | 0.519 | 0.922 | 0.767 | **Work (0-7 days after)** | 2.564 | 0.597 | 0.643 | 0.833 | 0.884 |
| | (0.106 to 0.421) | (0.112 to 2.139) | (0.122 to 2.206) | (0.226 to 3.759) | (0.134 to 4.392) | | (0.600 to 10.961) | (0.172 to 2.071) | (0.201 to 2.064) | (0.435 to 1.597) | (0.446 to 1.755) |
| **Work (20+ days before)** | 0.377** | 1.338 | 1.422 | 2.105 | 2.653 | **Work (8-14 days after)** | 7.849** | 5.722** | 4.507* | 2.172* | 2.216* |
| | (0.196 to 0.726) | (0.520 to 3.444) | (0.577 to 3.502) | (0.302 to 14.661) | (0.502 to 14.035) | | (2.207 to 27.915) | (1.625 to 20.153) | (1.343 to 15.129) | (1.152 to 4.094) | (1.168 to 4.204) |
| **Events (Not introduced)** | . | . | . | . | . | **Events (Not introduced)** | . | . | . | . | . |

| | | | | | | | | | | | |
|---|---|---|---|---|---|---|---|---|---|---|---|
| **Events (0-10 days before)** | 0.811 | 0.975 | 0.937 | 1.388 | 1.044 | **Events (before)** | 0.774 | 1.442 | 0.777 | 1.091 | 1.396 |
| | (0.351 to 1.872) | (0.501 to 1.900) | (0.504 to 1.743) | (0.824 to 2.340) | (0.657 to 1.659) | | (0.218 to 2.742) | (0.325 to 6.389) | (0.163 to 3.709) | (0.309 to 3.853) | (0.400 to 4.874) |
| **Events (11-20 days before)** | 0.328** | 0.903 | 0.935 | 1.327 | 1.230 | **Events (0-7 days after)** | 2.878 | 1.142 | 0.615 | 0.879 | 1.233 |
| | (0.146 to 0.739) | (0.349 to 2.340) | (0.362 to 2.414) | (0.694 to 2.537) | (0.654 to 2.314) | | (0.755 to 10.972) | (0.225 to 5.796) | (0.119 to 3.178) | (0.217 to 3.564) | (0.321 to 4.747) |
| **Events (20+ days before)** | 0.182*** | 0.354 | 0.338 | 0.647 | 0.723 | **Events (8-14 days after)** | 4.319 | 1.896 | 1.682 | 0.545 | 0.629 |
| | (0.085 to 0.388) | (0.124 to 1.011) | (0.114 to 1.008) | (0.137 to 3.060) | (0.164 to 3.185) | | (0.989 to 18.868) | (0.579 to 6.204) | (0.489 to 5.782) | (0.226 to 1.316) | (0.244 to 1.625) |
| **Gatherings (Not introduced)** | . | . | . | . | . | **Gatherings (Not introduced)** | . | . | . | . | . |
| **Gatherings (0-10 days before)** | 0.596 | 0.845 | 0.848 | 0.662 | 0.712 | **Gatherings (before)** | 0.246** | 0.961 | 0.649 | 0.709 | 0.738 |
| | (0.279 to 1.274) | (0.474 to 1.506) | (0.474 to 1.516) | (0.423 to 1.034) | (0.452 to 1.121) | | (0.090 to 0.673) | (0.328 to 2.820) | (0.204 to 2.060) | (0.338 to 1.488) | (0.367 to 1.484) |
| **Gatherings (11-20 days before)** | 0.361* | 0.953 | 0.909 | 1.177 | 1.036 | **Gatherings (0-7 days after)** | 1.545 | 3.962* | 1.684 | 1.224 | 1.328 |
| | (0.158 to 0.826) | (0.405 to 2.241) | (0.380 to 2.176) | (0.620 to 2.233) | (0.452 to 2.377) | | (0.469 to 5.091) | (1.087 to 14.439) | (0.466 to 6.092) | (0.438 to 3.420) | (0.501 to 3.520) |
| **Gatherings (20+ days before)** | 0.262*** | 2.084 | 2.344 | 2.977 | 2.450 | **Gatherings (8-14 days after)** | 0.423 | 0.021** | 0.026** | 0.026*** | 0.015*** |
| | (0.133 to 0.517) | (0.611 to 7.116) | (0.656 to 8.376) | (0.599 to 14.795) | (0.675 to 8.896) | | (0.098 to 1.836) | (0.002 to 0.238) | (0.003 to 0.240) | (0.007 to 0.090) | (0.005 to 0.051) |
| **Transport (Not introduced)** | . | . | . | . | . | **Transport (Not introduced)** | . | . | . | . | . |
| **Transport (0-10 days before)** | 1.018 | 0.953 | 0.950 | 1.232 | 1.342 | **Transport (before)** | 0.381* | 0.722 | 0.910 | 1.001 | 1.007 |
| | (0.472 to 2.199) | (0.474 to 1.916) | (0.473 to 1.912) | (0.873 to 1.738) | (0.866 to 2.078) | | (0.159 to 0.911) | (0.287 to 1.812) | (0.375 to 2.210) | (0.587 to 1.705) | (0.621 to 1.634) |

| | | | | | | | | | | | |
|---|---|---|---|---|---|---|---|---|---|---|---|
| **Transport (11-20 days before)** | 0.270*** | 0.804 | 0.788 | 0.942 | 1.182 | **Transport (0-7 days after)** | 0.428 | 0.718 | 1.161 | 0.845 | 0.718 |
| | (0.150 to 0.488) | (0.170 to 3.798) | (0.167 to 3.726) | (0.202 to 4.401) | (0.187 to 7.464) | | (0.171 to 1.072) | (0.301 to 1.714) | (0.486 to 2.773) | (0.442 to 1.616) | (0.378 to 1.363) |
| **Transport (20+ days before)** | 0.431** | 1.279 | 1.166 | 2.046 | 1.608 | **Transport (8-14 days after)** | 2.526 | 0.514 | 0.522 | 0.816 | 0.688 |
| | (0.241 to 0.773) | (0.210 to 7.787) | (0.217 to 6.271) | (0.194 to 21.548) | (0.184 to 14.063) | | (0.427 to 14.938) | (0.128 to 2.067) | (0.139 to 1.963) | (0.317 to 2.099) | (0.242 to 1.956) |
| **Stay home (Not introduced)** | . | . | . | . | . | **Stay home (Not introduced)** | . | . | . | . | . |
| **Stay home (0-10 days before)** | 0.670 | 0.996 | 0.927 | 1.098 | 0.869 | **Stay home (before)** | 0.423* | 2.097 | 1.939 | 2.008 | 2.029 |
| | (0.370 to 1.213) | (0.447 to 2.219) | (0.419 to 2.054) | (0.626 to 1.925) | (0.477 to 1.583) | | (0.180 to 0.992) | (0.473 to 9.301) | (0.414 to 9.083) | (0.924 to 4.365) | (0.972 to 4.237) |
| **Stay home (11-20 days before)** | 0.440 | 1.098 | 1.042 | 0.459 | 0.548 | **Stay home (0-7 days after)** | 1.773 | 0.968 | 1.264 | 1.219 | 1.508 |
| | (0.149 to 1.299) | (0.251 to 4.801) | (0.249 to 4.353) | (0.176 to 1.195) | (0.218 to 1.376) | | (0.534 to 5.882) | (0.320 to 2.929) | (0.411 to 3.885) | (0.570 to 2.609) | (0.663 to 3.429) |
| **Stay home (20+ days before)** | 0.243*** | 1.603 | 1.240 | 0.075 | 0.090* | **Stay home (8-14 days after)** | 2.015 | 0.547 | 0.772 | 0.899 | 1.026 |
| | (0.114 to 0.517) | (0.249 to 10.322) | (0.188 to 8.168) | (0.005 to 1.141) | (0.009 to 0.856) | | (0.524 to 7.753) | (0.198 to 1.507) | (0.273 to 2.183) | (0.419 to 1.927) | (0.534 to 1.972) |
| **Internal movement (Not introduced)** | . | . | . | . | . | **Internal movement (Not introduced)** | . | . | . | . | . |
| **Internal movement (0-10 days before)** | 0.592 | 0.774 | 0.821 | 0.887 | 0.988 | **Internal movement (before)** | 0.431 | 1.659 | 1.965 | 1.147 | 1.259 |
| | (0.318 to 1.101) | (0.350 to 1.711) | (0.377 to 1.786) | (0.481 to 1.635) | (0.561 to 1.742) | | (0.183 to 1.014) | (0.494 to 5.574) | (0.583 to 6.618) | (0.554 to 2.377) | (0.563 to 2.817) |
| **Internal movement (11-20 days before)** | 0.761 | 1.763 | 1.931 | 2.483* | 3.002* | **Internal movement** | 1.815 | 5.596** | 4.750** | 2.829** | 2.353* |

| | | | | | | | | | | | |
|---|---|---|---|---|---|---|---|---|---|---|---|
| | | | | | | **(0-7 days after)** | | | | | |
| | (0.347 to 1.665) | (0.457 to 6.811) | (0.500 to 7.450) | (1.175 to 5.250) | (1.150 to 7.837) | | (0.453 to 7.268) | (1.797 to 17.426) | (1.561 to 14.451) | (1.449 to 5.526) | (1.225 to 4.518) |
| **Internal movement (20+ days before)** | 0.179*** | 0.670 | 0.828 | 3.320 | 3.582 | **Internal movement (8-14 days after)** | 2.673 | 2.774 | 4.447* | 1.260 | 0.862 |
| | (0.070 to 0.455) | (0.127 to 3.532) | (0.163 to 4.217) | (0.288 to 38.221) | (0.410 to 31.278) | | (0.904 to 7.902) | (0.726 to 10.596) | (1.032 to 19.165) | (0.483 to 3.287) | (0.322 to 2.305) |
| **Travel (Not introduced)** | . | . | . | . | . | **Travel (Not introduced)** | . | . | . | . | . |
| **Travel (0-10 days before)** | 0.474 | 0.497 | 0.510 | 1.254 | 1.510 | **Travel (before)** | 0.159*** | 0.397 | 0.368 | 0.714 | 0.735 |
| | (0.167 to 1.351) | (0.223 to 1.106) | (0.238 to 1.092) | (0.771 to 2.039) | (0.900 to 2.536) | | (0.063 to 0.400) | (0.145 to 1.091) | (0.121 to 1.118) | (0.323 to 1.576) | (0.355 to 1.522) |
| **Travel (11-20 days before)** | 0.271** | 0.588 | 0.580 | 1.614 | 1.921* | **Travel (0-7 days after)** | 0.773 | 0.330 | 0.250* | 0.273* | 0.351 |
| | (0.114 to 0.646) | (0.237 to 1.459) | (0.234 to 1.436) | (0.874 to 2.981) | (1.015 to 3.638) | | (0.230 to 2.596) | (0.092 to 1.184) | (0.085 to 0.741) | (0.084 to 0.882) | (0.099 to 1.242) |
| **Travel (20+ days before)** | 0.164*** | 0.197*** | 0.206*** | 0.596* | 1.073 | **Travel (8-14 days after)** | 0.896 | 2.232 | 1.839 | 1.287 | 1.363 |
| | (0.077 to 0.347) | (0.082 to 0.478) | (0.088 to 0.483) | (0.356 to 0.997) | (0.620 to 1.858) | | (0.197 to 4.081) | (0.227 to 21.979) | (0.214 to 15.841) | (0.455 to 3.639) | (0.557 to 3.332) |
| **Public information (Not introduced)** | . | . | . | . | . | **Public information (Not introduced)** | . | . | . | . | . |
| **Public information (0-10 days before)** | 1.766 | 1.024 | 1.156 | 1.012 | 1.124 | **Public information (before)** | 1.289 | 0.874 | 2.197 | 1.161 | 0.928 |
| | (0.415 to 7.507) | (0.344 to 3.052) | (0.374 to 3.579) | (0.553 to 1.852) | (0.463 to 2.729) | | (0.391 to 4.250) | (0.066 to 11.585) | (0.168 to 28.709) | (0.238 to 5.670) | (0.161 to 5.354) |
| **Public information (11-20 days before)** | 1.033 | 1.497 | 1.529 | 0.524* | 0.403** | **Public information (0-7 days after)** | 2.298 | 0.207 | 0.688 | 0.673 | 0.549 |

| | | | | | | | | | | | |
|---|---|---|---|---|---|---|---|---|---|---|---|
| | (0.308 to 3.469) | (0.455 to 4.919) | (0.454 to 5.152) | (0.276 to 0.993) | (0.207 to 0.784) | | (0.482 to 10.946) | (0.011 to 3.924) | (0.040 to 11.897) | (0.150 to 3.023) | (0.108 to 2.797) |
| **Public information (20+ days before)** | 1.211 | 2.475 | 2.636 | 1.216 | 0.888 | **Public information (8-14 days after)** | 0.317 | 0.833 | 0.883 | 1.889 | 18.398* |
| | (0.363 to 4.035) | (0.815 to 7.520) | (0.835 to 8.326) | (0.674 to 2.195) | (0.508 to 1.552) | | (0.068 to 1.480) | (0.033 to 20.823) | (0.035 to 22.391) | (0.064 to 55.833) | (1.420 to 238.347) |
| **Testing (Not introduced)** | . | . | . | . | . | **Testing (Not introduced)** | . | . | . | . | . |
| **Testing (0-10 days before)** | 1.265 | 0.685 | 0.667 | 0.716 | 0.696 | **Testing (before)** | 2.630* | 1.868 | 1.608 | 0.922 | 0.880 |
| | (0.465 to 3.437) | (0.264 to 1.777) | (0.250 to 1.781) | (0.355 to 1.443) | (0.303 to 1.600) | | (1.172 to 5.899) | (0.684 to 5.102) | (0.670 to 3.860) | (0.461 to 1.845) | (0.450 to 1.719) |
| **Testing (11-20 days before)** | 3.683** | 1.293 | 1.308 | 0.921 | 1.107 | **Testing (0-7 days after)** | 1.110 | 2.199 | 2.216 | 2.312 | 2.105 |
| | (1.571 to 8.636) | (0.446 to 3.749) | (0.456 to 3.752) | (0.433 to 1.958) | (0.439 to 2.793) | | (0.369 to 3.342) | (0.326 to 14.822) | (0.532 to 9.236) | (0.882 to 6.061) | (0.859 to 5.161) |
| **Testing (20+ days before)** | 2.274* | 1.207 | 1.210 | 0.657 | 0.667 | **Testing (8-14 days after)** | 0.643 | 4.674 | 4.733 | 0.509 | 0.338 |
| | (1.096 to 4.720) | (0.516 to 2.824) | (0.517 to 2.831) | (0.348 to 1.242) | (0.275 to 1.618) | | (0.223 to 1.860) | (0.528 to 41.337) | (0.622 to 36.008) | (0.167 to 1.548) | (0.108 to 1.058) |
| **Contact tracing (Not introduced)** | . | . | . | . | . | **Contact tracing (Not introduced)** | . | . | . | . | . |
| **Contact tracing (0-10 days before)** | 1.388 | 1.830 | 2.118 | 3.910*** | 4.475*** | **Contact tracing (before)** | 2.774** | 0.960 | 0.815 | 0.926 | 1.042 |
| | (0.546 to 3.528) | (0.800 to 4.185) | (0.922 to 4.865) | (2.105 to 7.264) | (2.200 to 9.101) | | (1.377 to 5.588) | (0.384 to 2.400) | (0.324 to 2.052) | (0.496 to 1.728) | (0.538 to 2.018) |
| **Contact tracing (11-20 days before)** | 3.415** | 3.745** | 3.845** | 3.668*** | 3.540*** | **Contact tracing (0-7 days after)** | 0.742 | 0.820 | 0.849 | 1.103 | 1.095 |
| | (1.569 to 7.433) | (1.533 to 9.148) | (1.607 to 9.200) | (2.095 to 6.421) | (2.098 to 5.975) | | (0.277 to 1.985) | (0.182 to 3.686) | (0.215 to 3.352) | (0.510 to 2.386) | (0.546 to 2.197) |

| | | | | | | | | | | | |
|---|---|---|---|---|---|---|---|---|---|---|---|
| **Contact tracing (20+ days before)** | 1.892 | 2.236* | 1.863 | 2.009* | 1.440 | **Contact tracing (8-14 days after)** | 0.531 | 2.359 | 3.599 | 3.435 | 3.126 |
| | (0.838 to 4.272) | (1.167 to 4.284) | (0.968 to 3.585) | (1.137 to 3.547) | (0.781 to 2.655) | | (0.140 to 2.016) | (0.150 to 37.219) | (0.304 to 42.611) | (0.303 to 38.976) | (0.723 to 13.520) |
| **Date of first case** | | | 0.986 | 0.996 | 0.985* | | | | 0.967*** | 0.989 | 0.995 |
| | | | (0.970 to 1.001) | (0.982 to 1.009) | (0.972 to 0.999) | | | | (0.948 to 0.986) | (0.970 to 1.009) | (0.976 to 1.014) |
| **Population density (people per sq.km)** | | | | 1.000 | 1.000 | | | | | 1.000 | 1.000 |
| | | | | (1.000 to 1.000) | (1.000 to 1.000) | | | | | (1.000 to 1.000) | (1.000 to 1.000) |
| **% Population aged 65+** | | | | 1.144*** | 1.177*** | | | | | 0.991 | 1.000 |
| | | | | (1.069 to 1.224) | (1.102 to 1.258) | | | | | (0.908 to 1.081) | (0.916 to 1.092) |
| **% Population male** | | | | 1.065* | 1.056* | | | | | 0.985 | 0.988 |
| | | | | (1.004 to 1.131) | (1.002 to 1.114) | | | | | (0.921 to 1.054) | (0.926 to 1.055) |
| **Life expectancy at birth (years)** | | | | 1.038 | 1.064 | | | | | 1.074 | 1.090 |
| | | | | (0.977 to 1.104) | (0.991 to 1.142) | | | | | (0.956 to 1.207) | (0.978 to 1.215) |
| **Hospital beds (per 1000 people)** | | | | 0.798** | 0.828* | | | | | 0.923 | 0.944 |
| | | | | (0.693 to 0.918) | (0.711 to 0.964) | | | | | (0.816 to 1.043) | (0.835 to 1.068) |
| **Physicians (per 1000 people)** | | | | 1.012 | 0.914 | | | | | 0.915 | 0.956 |
| | | | | (0.830 to 1.233) | (0.753 to 1.110) | | | | | (0.694 to 1.206) | (0.734 to 1.246) |
| **GDP PPP (current international $)** | | | | 1.000 | 1.000 | | | | | 1.000 | 1.000 |
| | | | | (1.000 to 1.000) | (1.000 to 1.000) | | | | | (1.000 to 1.000) | (1.000 to 1.000) |

| | | | | | | | | | | | |
|---|---|---|---|---|---|---|---|---|---|---|---|
| **Manufacturing value added (%GDP)** | | | | 0.969** | 0.969* | | | | | 0.997 | 1.001 |
| | | | | (0.948 to 0.990) | (0.944 to 0.995) | | | | | (0.958 to 1.037) | (0.962 to 1.041) |
| **Health expenditure (%GDP)** | | | | 0.978 | 0.981 | | | | | 1.119 | 1.097 |
| | | | | (0.899 to 1.064) | (0.892 to 1.079) | | | | | (0.990 to 1.265) | (0.953 to 1.264) |
| **International tourism, number of arrivals** | | | | 1.000 | 1.000*** | | | | | 1.000 | 1.000 |
| | | | | (1.000 to 1.000) | (1.000 to 1.000) | | | | | (1.000 to 1.000) | (1.000 to 1.000) |
| **Governance (Voice and Accountability)** | | | | 0.755 | 0.684 | | | | | 1.466 | 1.293 |
| | | | | (0.507 to 1.124) | (0.426 to 1.099) | | | | | (0.855 to 2.515) | (0.773 to 2.162) |
| **East Asia & Pacific** | | | | . | . | | | | | . | . |
| **Europe & Central Asia** | | | | 8.921*** | 7.369*** | | | | | 42.923*** | 34.261*** |
| | | | | (3.422 to 23.256) | (2.400 to 22.627) | | | | | (10.721 to 171.843) | (7.680 to 152.840) |
| **America & Caribbean** | | | | 4.127** | 4.469* | | | | | 11.357*** | 9.861** |
| | | | | (1.521 to 11.198) | (1.413 to 14.137) | | | | | (2.767 to 46.611) | (2.291 to 42.440) |
| **East & North Africa** | | | | 2.622 | 2.477 | | | | | 13.349** | 10.359** |
| | | | | (0.947 to 7.262) | (0.754 to 8.133) | | | | | (2.753 to 64.714) | (2.316 to 46.340) |
| **North America** | | | | 3.363 | 1.105 | | | | | 3.186 | 1.371 |
| | | | | (0.878 to 12.885) | (0.281 to 4.354) | | | | | (0.323 to 31.425) | (0.183 to 10.267) |
| **South Asia** | | | | 0.736 | 0.784 | | | | | 2.560 | 2.147 |

| | | | | | | | | | | | |
|---|---|---|---|---|---|---|---|---|---|---|---|
| | | | | (0.198 to 2.741) | (0.169 to 3.634) | | | | | (0.591 to 11.097) | (0.568 to 8.112) |
| **Sub-Saharan Africa** | | | | 1.386 | 1.711 | | | | | 3.980 | 4.510 |
| | | | | (0.438 to 4.385) | (0.469 to 6.240) | | | | | (0.635 to 24.935) | (0.742 to 27.422) |
| **Lnalpha** | | 0.941 | 0.904 | 0.452** | 0.000*** | | | 0.645 | 0.641 | 0.133*** | 0.006* |
| | | (0.642 to 1.379) | (0.617 to 1.323) | (0.268 to 0.762) | (0.000 to 0.000) | | | (0.377 to 1.106) | (0.382 to 1.075) | (0.077 to 0.232) | (0.000 to 0.801) |
| **Observations** | 3250 | 3250 | 3250 | 3250 | 3250 | 3150 | 3150 | 3150 | 3150 | 3150 | 3150 |
| **Pseudo R-squared** | | 0.165 | 0.170 | 0.284 | 0.415 | | | 0.229 | 0.245 | 0.416 | 0.461 |

**p-values in parentheses =** * p<0.05 ** p<0.01 *** p<0.001; # includes time, day of week and week of year fixed effects; ~ model does not converge

#### Regression results – Dropping China/Belgium specifications

*Regression results, mean policy strictness (combination of timing and strictness)*

| | 24 Days (Policies implemented before first death) | | | | | 38 Days (Policies implemented within 14 days after first death) | | | | |
|---|---|---|---|---|---|---|---|---|---|---|
| **Average deaths per day over time period, per million** | **(1)** | **(2)** | **(3)** | **(4)** | **(5) #** | **(1)** | **(2)** | **(3)** | **(4)** | **(5) #** |
| **School** | -0.202** | -0.239* | -0.244* | -0.155 | -0.111 | -0.922** | -0.964* | -0.853* | -0.903** | -0.834** |
| | (-0.329 to -0.076) | (-0.428 to -0.049) | (-0.433 to -0.054) | (-0.318 to 0.007) | (-0.269 to 0.047) | (-1.506 to -0.339) | (-1.700 to -0.228) | (-1.528 to -0.177) | (-1.495 to -0.311) | (-1.458 to -0.210) |
| **Work** | -0.164* | 0.015 | 0.014 | -0.165* | -0.188* | -0.573* | -0.035 | -0.043 | -0.397* | -0.445* |
| | (-0.314 to -0.014) | (-0.137 to 0.166) | (-0.135 to 0.162) | (-0.308 to -0.021) | (-0.345 to -0.031) | (-1.068 to -0.079) | (-0.377 to 0.306) | (-0.388 to 0.302) | (-0.756 to -0.038) | (-0.822 to -0.068) |
| **Events** | -0.247** | 0.075 | 0.072 | -0.015 | -0.023 | -0.968** | 0.154 | 0.147 | 0.150 | 0.261 |
| | (-0.425 to -0.068) | (-0.160 to 0.310) | (-0.160 to 0.304) | (-0.198 to 0.168) | (-0.232 to 0.186) | (-1.633 to -0.303) | (-0.584 to 0.891) | (-0.595 to 0.888) | (-0.437 to 0.738) | (-0.413 to 0.935) |
| **Gatherings** | -0.137* | -0.000 | 0.000 | 0.140* | 0.159* | -0.514* | 0.042 | 0.044 | 0.327* | 0.339* |
| | (-0.246 to -0.027) | (-0.166 to 0.166) | (-0.167 to 0.167) | (0.016 to 0.264) | (0.034 to 0.285) | (-0.935 to -0.093) | (-0.337 to 0.421) | (-0.328 to 0.416) | (0.052 to 0.602) | (0.059 to 0.620) |
| **Transport** | -0.199* | 0.132 | 0.135 | 0.128 | 0.088 | -0.650* | 0.407 | 0.392 | 0.297 | 0.267 |
| | (-0.352 to -0.045) | (-0.151 to 0.415) | (-0.149 to 0.419) | (-0.122 to 0.378) | (-0.162 to 0.338) | (-1.166 to -0.134) | (-0.145 to 0.958) | (-0.132 to 0.915) | (-0.232 to 0.825) | (-0.258 to 0.791) |
| **Stay home** | -0.203* | 0.023 | 0.025 | -0.077 | -0.025 | -0.599** | 0.522 | 0.528 | 0.220 | 0.233 |
| | (-0.358 to -0.048) | (-0.245 to 0.291) | (-0.244 to 0.294) | (-0.282 to 0.127) | (-0.252 to 0.202) | (-1.028 to -0.170) | (-0.161 to 1.206) | (-0.150 to 1.207) | (-0.322 to 0.762) | (-0.364 to 0.830) |

| | | | | | | | | | | |
|---|---|---|---|---|---|---|---|---|---|---|
| **Internal movement** | -0.206 | 0.005 | 0.002 | 0.096 | 0.088 | -0.876** | -0.507 | -0.500 | -0.176 | -0.137 |
| | (-0.433 to 0.020) | (-0.347 to 0.357) | (-0.363 to 0.367) | (-0.109 to 0.302) | (-0.149 to 0.325) | (-1.463 to -0.288) | (-1.107 to 0.094) | (-1.074 to 0.075) | (-0.585 to 0.234) | (-0.590 to 0.315) |
| **Travel** | -0.151** | -0.175* | -0.176* | -0.069 | -0.067 | -0.749*** | -0.834*** | -0.834*** | -0.475* | -0.386 |
| | (-0.257 to -0.045) | (-0.309 to -0.041) | (-0.309 to -0.042) | (-0.180 to 0.042) | (-0.186 to 0.052) | (-1.184 to -0.313) | (-1.323 to -0.345) | (-1.322 to -0.347) | (-0.888 to -0.062) | (-0.863 to 0.091) |
| **Public information** | 0.004 | 0.114 | 0.111 | -0.056 | -0.070 | -0.083 | 0.651 | 0.686 | 0.297 | 0.210 |
| | (-0.171 to 0.179) | (-0.126 to 0.354) | (-0.146 to 0.369) | (-0.265 to 0.153) | (-0.274 to 0.135) | (-0.689 to 0.523) | (-0.244 to 1.546) | (-0.255 to 1.627) | (-0.570 to 1.164) | (-0.606 to 1.026) |
| **Testing** | 0.085 | 0.098 | 0.100 | 0.201* | 0.200* | 0.115 | 0.191 | 0.152 | 0.509* | 0.549* |
| | (-0.033 to 0.203) | (-0.072 to 0.268) | (-0.078 to 0.279) | (0.043 to 0.358) | (0.024 to 0.375) | (-0.137 to 0.368) | (-0.192 to 0.574) | (-0.273 to 0.577) | (0.091 to 0.926) | (0.090 to 1.008) |
| **Contact tracing** | 0.132 | 0.184* | 0.192* | 0.114 | 0.097 | 0.406 | 0.625** | 0.542* | 0.426 | 0.424 |
| | (-0.034 to 0.298) | (0.006 to 0.363) | (0.023 to 0.360) | (-0.050 to 0.278) | (-0.063 to 0.257) | (-0.049 to 0.861) | (0.161 to 1.089) | (0.108 to 0.977) | (-0.005 to 0.857) | (-0.029 to 0.878) |
| **Date of first case** | | | 0.001 | 0.007* | 0.005 | | | -0.007 | 0.004 | 0.004 |
| | | | (-0.005 to 0.007) | (0.001 to 0.013) | (-0.002 to 0.013) | | | (-0.027 to 0.012) | (-0.015 to 0.023) | (-0.020 to 0.029) |
| **Population density (people per sq.km)** | | | | -0.000 | -0.000 | | | | -0.000* | -0.000* |
| | | | | (-0.000 to 0.000) | (-0.000 to 0.000) | | | | (-0.001 to -0.000) | (-0.001 to -0.000) |
| **% Population aged 65+** | | | | 0.015 | 0.014 | | | | 0.003 | 0.014 |
| | | | | (-0.019 to 0.048) | (-0.024 to 0.051) | | | | (-0.097 to 0.104) | (-0.100 to 0.128) |
| **% Population male** | | | | 0.002 | -0.001 | | | | -0.003 | 0.000 |

| | | | | | | | | | | |
|---|---|---|---|---|---|---|---|---|---|---|
| | | | | (-0.015 to 0.019) | (-0.020 to 0.018) | | | | (-0.064 to 0.058) | (-0.062 to 0.062) |
| **Life expectancy at birth (years)** | | | | 0.017 | 0.023* | | | | 0.045 | 0.045 |
| | | | | (-0.000 to 0.035) | (0.002 to 0.043) | | | | (-0.015 to 0.105) | (-0.017 to 0.107) |
| **Hospital beds (per 1000 people)** | | | | -0.075** | -0.064* | | | | -0.223** | -0.208* |
| | | | | (-0.122 to -0.028) | (-0.113 to -0.015) | | | | (-0.376 to -0.071) | (-0.373 to -0.043) |
| **Physicians (per 1000 people)** | | | | 0.046 | 0.027 | | | | 0.192 | 0.146 |
| | | | | (-0.049 to 0.140) | (-0.054 to 0.108) | | | | (-0.116 to 0.499) | (-0.138 to 0.430) |
| **GDP PPP (current international $)** | | | | -0.000 | -0.000 | | | | -0.000 | -0.000 |
| | | | | (-0.000 to 0.000) | (-0.000 to 0.000) | | | | (-0.000 to 0.000) | (-0.000 to 0.000) |
| **Manufacturing value added (%GDP)** | | | | -0.009 | -0.011 | | | | -0.006 | -0.010 |
| | | | | (-0.023 to 0.005) | (-0.025 to 0.004) | | | | (-0.040 to 0.027) | (-0.046 to 0.026) |
| **Health expenditure (%GDP)** | | | | -0.023 | -0.011 | | | | -0.055 | -0.020 |
| | | | | (-0.069 to 0.023) | (-0.062 to 0.039) | | | | (-0.200 to 0.089) | (-0.145 to 0.105) |
| **International tourism, number of arrivals** | | | | 0.000 | 0.000 | | | | 0.000 | 0.000 |
| | | | | (-0.000 to 0.000) | (-0.000 to 0.000) | | | | (-0.000 to 0.000) | (-0.000 to 0.000) |

| | | | | | | | | | | |
|---|---|---|---|---|---|---|---|---|---|---|
| **Governance (Voice and Accountability)** | | | | 0.109 | 0.095 | | | | 0.156 | 0.072 |
| | | | | (-0.043 to 0.261) | (-0.091 to 0.280) | | | | (-0.290 to 0.602) | (-0.422 to 0.566) |
| **East Asia & Pacific** | | | | . | . | | | | . | . |
| **Europe & Central Asia** | | | | 0.464** | 0.330 | | | | 1.762*** | 1.501** |
| | | | | (0.134 to 0.794) | (-0.121 to 0.780) | | | | (0.809 to 2.714) | (0.392 to 2.609) |
| **America & Caribbean** | | | | 0.100 | -0.004 | | | | 0.855 | 0.727 |
| | | | | (-0.161 to 0.361) | (-0.338 to 0.330) | | | | (-0.030 to 1.740) | (-0.276 to 1.730) |
| **East & North Africa** | | | | 0.169 | 0.089 | | | | 0.853 | 0.624 |
| | | | | (-0.089 to 0.426) | (-0.219 to 0.397) | | | | (-0.098 to 1.804) | (-0.383 to 1.630) |
| **North America** | | | | -0.295 | -0.510 | | | | -0.017 | -0.564 |
| | | | | (-0.835 to 0.246) | (-1.069 to 0.049) | | | | (-1.992 to 1.957) | (-2.316 to 1.187) |
| **South Asia** | | | | 0.093 | -0.012 | | | | 0.584 | 0.503 |
| | | | | (-0.304 to 0.490) | (-0.399 to 0.376) | | | | (-0.594 to 1.762) | (-0.750 to 1.756) |
| **Sub-Saharan Africa** | | | | 0.200 | 0.174 | | | | 1.465* | 1.438* |
| | | | | (-0.115 to 0.515) | (-0.184 to 0.531) | | | | (0.260 to 2.670) | (0.118 to 2.758) |
| **Constant** | | 0.359*** | -12.040 | -158.851* | -118.323 | | 1.834*** | 166.139 | -91.670 | -100.524 |
| | | (0.154 to 0.564) | (-144.787 to 120.706) | (-291.366 to -26.336) | (-277.350 to 40.704) | | (1.015 to 2.653) | (-252.828 to 585.107) | (-507.200 to 323.861) | (-636.490 to 435.443) |
| **Observations** | 3200 | 3200 | 3200 | 3200 | 3200 | 3100 | 3100 | 3100 | 3100 | 3100 |
| **R-squared** | | 0.074 | 0.074 | 0.201 | 0.282 | | 0.242 | 0.244 | 0.462 | 0.490 |

| | | | | | | | | | | |
|---|---|---|---|---|---|---|---|---|---|---|
| **Adjusted R-squared** | | 0.070 | 0.070 | 0.194 | 0.265 | | 0.239 | 0.241 | 0.457 | 0.477 |

***p-values in parentheses** = * p<0.05  ** p<0.01  *** p<0.001; # includes time, day of week and week of year fixed effects*

*Regression results, policy strictness. Baseline is policy not introduced within policy analysis period*

| | **24 Days (Policies implemented before first death)** | | | | | **38 Days (Policies implemented within 14 days after first death)** | | | | |
|---|---|---|---|---|---|---|---|---|---|---|
| **Average deaths per day over time period, per million** | **(1)** | **(2)** | **(3)** | **(4)** | **(5) #** | **(1)** | **(2)** | **(3)** | **(4)** | **(5) #** |
| **0bn.School** | . | . | . | . | . | . | . | . | . | . |
| **1.School** | -0.492** | -0.391 | -0.405 | -0.098 | -0.479 | 4.823*** | 6.288*** | 6.976*** | 4.598*** | 4.206*** |
| | (-0.835 to -0.150) | (-0.829 to 0.047) | (-0.862 to 0.052) | (-0.963 to 0.768) | (-1.578 to 0.620) | (3.510 to 6.136) | (4.362 to 8.214) | (4.625 to 9.327) | (2.118 to 7.078) | (1.894 to 6.517) |
| **2.School** | -0.350 | -0.087 | -0.059 | -0.233 | -0.390** | -0.399 | 0.466 | 1.338 | 0.318 | 0.068 |
| | (-0.748 to 0.048) | (-0.369 to 0.196) | (-0.344 to 0.227) | (-0.517 to 0.050) | (-0.669 to -0.110) | (-1.821 to 1.022) | (-1.093 to 2.024) | (-0.490 to 3.166) | (-1.478 to 2.113) | (-1.739 to 1.876) |
| **3.School** | -0.378* | -0.233 | -0.209 | -0.257 | -0.344* | -0.280 | 0.675 | 1.495 | 0.425 | 0.138 |
| | (-0.726 to -0.029) | (-0.568 to 0.102) | (-0.546 to 0.127) | (-0.545 to 0.031) | (-0.651 to -0.037) | (-1.635 to 1.076) | (-0.792 to 2.142) | (-0.403 to 3.394) | (-0.815 to 1.665) | (-1.058 to 1.333) |
| **0bn.Work** | . | . | . | . | . | . | . | . | . | . |
| **1.Work** | 0.141 | 0.321* | 0.329* | 0.073 | 0.025 | 0.607 | -0.251 | -0.331 | -0.263 | -0.335 |
| | (-0.296 to 0.579) | (0.015 to 0.628) | (0.025 to 0.633) | (-0.204 to 0.349) | (-0.268 to 0.319) | (-0.627 to 1.842) | (-1.299 to 0.797) | (-1.326 to 0.664) | (-1.044 to 0.518) | (-1.103 to 0.434) |
| **2.Work** | -0.065 | 0.230 | 0.226 | 0.107 | 0.085 | 0.571 | 0.438 | 0.313 | -0.430 | -0.538 |
| | (-0.298 to 0.167) | (-0.041 to 0.501) | (-0.048 to 0.500) | (-0.091 to 0.305) | (-0.110 to 0.279) | (-0.627 to 1.769) | (-0.699 to 1.575) | (-0.738 to 1.364) | (-1.069 to 0.209) | (-1.224 to 0.147) |
| **3.Work** | -0.113 | 0.341* | 0.342* | 0.126 | 0.104 | 0.040 | 0.039 | 0.001 | -0.301 | -0.468 |
| | (-0.317 to 0.092) | (0.082 to 0.600) | (0.080 to 0.603) | (-0.107 to 0.359) | (-0.133 to 0.341) | (-0.609 to 0.690) | (-0.939 to 1.017) | (-0.918 to 0.919) | (-0.922 to 0.320) | (-1.147 to 0.211) |
| **0bn.Events** | . | . | . | . | . | . | . | . | . | . |
| **1.Events** | -0.262 | -0.278 | -0.275 | -0.085 | -0.074 | -0.295 | -1.403 | -1.503 | -0.215 | -0.119 |
| | (-0.716 to 0.191) | (-0.671 to 0.115) | (-0.662 to 0.112) | (-0.434 to 0.264) | (-0.463 to 0.314) | (-1.428 to 0.838) | (-3.303 to 0.498) | (-3.352 to 0.347) | (-1.186 to 0.757) | (-1.256 to 1.018) |
| **2.Events** | -0.296 | -0.172 | -0.172 | -0.131 | -0.177 | -0.042 | 0.125 | 0.171 | 0.814* | 0.946* |
| | (-0.639 to 0.046) | (-0.508 to 0.163) | (-0.507 to 0.164) | (-0.364 to 0.102) | (-0.437 to 0.082) | (-0.958 to 0.874) | (-1.086 to 1.336) | (-0.948 to 1.290) | (0.028 to 1.600) | (0.130 to 1.761) |

| | | | | | | | | | | |
|---|---|---|---|---|---|---|---|---|---|---|
| **0bn.Gatherings** | . | . | . | . | . | . | . | . | . | . |
| **1.Gatherings** | -0.040 | -0.023 | -0.007 | -0.061 | 0.009 | -1.128 | -1.317 | -1.264 | -2.153** | -2.231** |
| | (-0.575 to 0.496) | (-0.356 to 0.310) | (-0.334 to 0.320) | (-0.309 to 0.187) | (-0.302 to 0.320) | (-2.513 to 0.257) | (-3.510 to 0.875) | (-3.117 to 0.590) | (-3.760 to -0.545) | (-3.802 to -0.659) |
| **2.Gatherings** | -0.108 | -0.078 | -0.091 | -0.112 | -0.139 | -0.034 | -2.025 | -2.713* | -1.003 | -0.653 |
| | (-0.416 to 0.199) | (-0.399 to 0.244) | (-0.435 to 0.252) | (-0.583 to 0.360) | (-0.608 to 0.329) | (-2.418 to 2.350) | (-4.141 to 0.092) | (-5.266 to -0.159) | (-2.311 to 0.305) | (-1.912 to 0.606) |
| **3.Gatherings** | -0.268* | -0.023 | -0.009 | 0.128 | 0.128 | -1.089 | -1.930 | -1.618 | -0.636 | -0.583 |
| | (-0.515 to -0.020) | (-0.244 to 0.197) | (-0.245 to 0.226) | (-0.133 to 0.390) | (-0.154 to 0.410) | (-2.486 to 0.307) | (-4.076 to 0.217) | (-3.336 to 0.099) | (-1.547 to 0.276) | (-1.501 to 0.335) |
| **4.Gatherings** | -0.142 | 0.122 | 0.120 | 0.199 | 0.204 | -0.893 | -1.841 | -1.822 | -0.966* | -0.930* |
| | (-0.428 to 0.143) | (-0.142 to 0.386) | (-0.146 to 0.385) | (-0.150 to 0.548) | (-0.181 to 0.589) | (-2.263 to 0.477) | (-4.092 to 0.411) | (-3.839 to 0.194) | (-1.858 to -0.075) | (-1.821 to -0.039) |
| **0bn.Transport** | . | . | . | . | . | . | . | . | . | . |
| **1.Transport** | 0.066 | 0.081 | 0.081 | 0.186 | 0.160 | 0.100 | 0.265 | 0.446 | 0.454 | 0.377 |
| | (-0.269 to 0.402) | (-0.158 to 0.319) | (-0.159 to 0.320) | (-0.062 to 0.433) | (-0.091 to 0.411) | (-0.877 to 1.078) | (-0.512 to 1.042) | (-0.391 to 1.284) | (-0.119 to 1.027) | (-0.195 to 0.949) |
| **2.Transport** | -0.130 | -0.014 | -0.019 | 0.012 | -0.056 | -0.571* | 0.216 | 0.171 | 0.604* | 0.596* |
| | (-0.297 to 0.038) | (-0.220 to 0.191) | (-0.230 to 0.192) | (-0.211 to 0.234) | (-0.304 to 0.193) | (-1.046 to -0.096) | (-0.288 to 0.719) | (-0.313 to 0.655) | (0.052 to 1.156) | (0.065 to 1.126) |
| **0bn.Stay home** | . | . | . | . | . | . | . | . | . | . |
| **1.Stay home** | -0.012 | 0.110 | 0.098 | -0.095 | -0.118 | 0.028 | 0.385 | 0.373 | 0.047 | 0.017 |
| | (-0.259 to 0.234) | (-0.101 to 0.320) | (-0.122 to 0.318) | (-0.291 to 0.101) | (-0.336 to 0.100) | (-0.864 to 0.919) | (-0.390 to 1.159) | (-0.322 to 1.067) | (-0.511 to 0.605) | (-0.546 to 0.580) |
| **2.Stay home** | -0.204* | -0.136 | -0.148 | -0.184 | -0.218 | -0.082 | 0.680 | 0.615 | -0.010 | 0.004 |
| | (-0.385 to -0.023) | (-0.352 to 0.080) | (-0.376 to 0.079) | (-0.429 to 0.061) | (-0.501 to 0.065) | (-0.969 to 0.805) | (-0.386 to 1.746) | (-0.326 to 1.556) | (-0.600 to 0.579) | (-0.619 to 0.627) |
| **3.Stay home** | -0.156 | -0.152 | -0.162 | -0.113 | -0.113 | -0.688* | 0.704 | 0.711 | 0.445 | 0.576 |
| | (-0.380 to 0.068) | (-0.450 to 0.147) | (-0.469 to 0.146) | (-0.428 to 0.203) | (-0.447 to 0.221) | (-1.356 to -0.020) | (-0.526 to 1.935) | (-0.423 to 1.846) | (-0.328 to 1.219) | (-0.254 to 1.406) |
| **0bn.Internal movement** | . | . | . | . | . | . | . | . | . | . |

| | | | | | | | | | | |
|---|---|---|---|---|---|---|---|---|---|---|
| **1.Internal movement** | -0.082 | 0.100 | 0.103 | -0.074 | -0.086 | 0.829 | 1.007 | 0.761 | 0.695 | 0.704 |
| | (-0.321 to 0.157) | (-0.071 to 0.271) | (-0.069 to 0.275) | (-0.232 to 0.085) | (-0.255 to 0.083) | (-0.504 to 2.163) | (-0.248 to 2.261) | (-0.293 to 1.814) | (-0.119 to 1.508) | (-0.162 to 1.570) |
| **2.Internal movement** | -0.192* | -0.020 | -0.012 | -0.016 | 0.009 | -0.467 | -0.452 | -0.475 | -0.423* | -0.428* |
| | (-0.382 to -0.003) | (-0.315 to 0.275) | (-0.311 to 0.287) | (-0.218 to 0.186) | (-0.214 to 0.232) | (-1.128 to 0.194) | (-0.953 to 0.050) | (-0.974 to 0.023) | (-0.801 to -0.045) | (-0.826 to -0.030) |
| **0bn.Travel** | . | . | . | . | . | . | . | . | . | . |
| **1.Travel** | -0.861* | -0.695* | -0.697* | -0.449 | -0.568 | -4.457*** | -3.396** | -2.148 | -3.280*** | -3.372*** |
| | (-1.533 to -0.189) | (-1.296 to -0.095) | (-1.297 to -0.098) | (-0.972 to 0.075) | (-1.204 to 0.068) | (-6.763 to -2.152) | (-5.803 to -0.988) | (-5.160 to 0.864) | (-5.184 to -1.376) | (-5.185 to -1.558) |
| **2.Travel** | -0.793* | -0.710* | -0.711* | -0.514 | -0.551 | -4.320*** | -4.339*** | -4.250*** | -4.457*** | -4.247*** |
| | (-1.502 to -0.083) | (-1.266 to -0.154) | (-1.268 to -0.155) | (-1.187 to 0.160) | (-1.248 to 0.146) | (-6.632 to -2.007) | (-6.149 to -2.529) | (-6.149 to -2.352) | (-6.130 to -2.785) | (-5.827 to -2.667) |
| **3.Travel** | -0.773* | -0.790* | -0.793* | -0.385 | -0.466 | -3.324** | -3.177*** | -3.078** | -2.761*** | -2.886*** |
| | (-1.457 to -0.089) | (-1.430 to -0.150) | (-1.434 to -0.151) | (-0.950 to 0.180) | (-1.078 to 0.146) | (-5.731 to -0.918) | (-4.923 to -1.432) | (-4.938 to -1.219) | (-4.053 to -1.470) | (-4.036 to -1.737) |
| **4.Travel** | -0.889** | -0.909** | -0.897** | -0.441 | -0.493 | -4.032*** | -4.043*** | -3.804*** | -3.105*** | -3.122*** |
| | (-1.558 to -0.221) | (-1.534 to -0.283) | (-1.511 to -0.283) | (-1.031 to 0.149) | (-1.111 to 0.125) | (-6.362 to -1.702) | (-5.577 to -2.508) | (-5.513 to -2.095) | (-4.350 to -1.859) | (-4.272 to -1.973) |
| **0bn. Public information** | . | . | . | . | . | . | . | . | . | . |
| **1.Public information** | -0.193 | 0.256 | 0.283 | 0.440 | 0.386 | -0.246 | 0.723 | 0.946 | 0.894 | 0.838 |
| | (-0.534 to 0.147) | (-0.293 to 0.805) | (-0.318 to 0.885) | (-0.191 to 1.071) | (-0.275 to 1.046) | (-1.085 to 0.592) | (-1.338 to 2.784) | (-1.368 to 3.260) | (-0.319 to 2.107) | (-0.506 to 2.181) |
| **2.Public information** | 0.022 | 0.363 | 0.374 | 0.387 | 0.380 | 0.091 | 1.624 | 1.768 | 0.211 | 0.163 |
| | (-0.329 to 0.373) | (-0.182 to 0.909) | (-0.195 to 0.943) | (-0.236 to 1.009) | (-0.259 to 1.020) | (-0.726 to 0.909) | (-0.444 to 3.693) | (-0.338 to 3.874) | (-0.836 to 1.257) | (-1.097 to 1.424) |
| **0bn.Testing** | . | . | . | . | . | . | . | . | . | . |
| **1.Testing** | 0.185* | 0.039 | 0.036 | -0.055 | -0.103 | 0.432 | 0.367 | 0.404 | 0.567 | 0.438 |

| | | | | | | | | | | |
|---|---|---|---|---|---|---|---|---|---|---|
| | (0.007 to 0.362) | (-0.225 to 0.304) | (-0.231 to 0.303) | (-0.357 to 0.246) | (-0.415 to 0.209) | (-0.186 to 1.049) | (-0.357 to 1.092) | (-0.366 to 1.175) | (-0.160 to 1.295) | (-0.310 to 1.186) |
| **2.Testing** | 0.187 | 0.006 | 0.007 | -0.030 | -0.059 | 0.457 | 0.255 | 0.252 | 0.187 | 0.177 |
| | (-0.029 to 0.403) | (-0.269 to 0.280) | (-0.265 to 0.280) | (-0.317 to 0.257) | (-0.379 to 0.262) | (-0.085 to 1.000) | (-0.549 to 1.059) | (-0.557 to 1.062) | (-0.571 to 0.945) | (-0.608 to 0.963) |
| **3.Testing** | 0.109 | 0.059 | 0.046 | 0.153 | 0.235 | 0.163 | 0.530 | 0.199 | 0.714 | 0.869 |
| | (-0.271 to 0.489) | (-0.405 to 0.523) | (-0.437 to 0.530) | (-0.298 to 0.603) | (-0.322 to 0.792) | (-0.260 to 0.587) | (-0.235 to 1.295) | (-0.594 to 0.992) | (-0.267 to 1.695) | (-0.236 to 1.974) |
| **0bn.Contact tracing** | . | . | . | . | . | . | . | . | . | . |
| **1.Contact tracing** | 0.060 | 0.128 | 0.128 | 0.171 | 0.196* | 0.157 | 0.359 | 0.266 | 0.422 | 0.504 |
| | (-0.075 to 0.195) | (-0.099 to 0.355) | (-0.098 to 0.353) | (-0.002 to 0.344) | (0.013 to 0.378) | (-0.174 to 0.488) | (-0.364 to 1.082) | (-0.425 to 0.958) | (-0.341 to 1.185) | (-0.276 to 1.283) |
| **2.Contact tracing** | 0.263* | 0.287* | 0.280* | 0.261* | 0.255* | 0.660 | 0.636 | 0.452 | 0.877* | 0.993** |
| | (0.023 to 0.502) | (0.010 to 0.563) | (0.006 to 0.554) | (0.032 to 0.490) | (0.027 to 0.482) | (-0.002 to 1.323) | (-0.183 to 1.456) | (-0.269 to 1.173) | (0.200 to 1.554) | (0.279 to 1.708) |
| **Date of first case** | | | -0.002 | 0.007 | 0.004 | | | -0.032* | -0.002 | 0.007 |
| | | | (-0.008 to 0.005) | (-0.001 to 0.014) | (-0.003 to 0.011) | | | (-0.060 to - 0.004) | (-0.017 to 0.014) | (-0.012 to 0.027) |
| **Population density (people per sq.km)** | | | | -0.000 | -0.000 | | | | -0.000* | -0.000* |
| | | | | (-0.000 to 0.000) | (-0.000 to 0.000) | | | | (-0.000 to - 0.000) | (-0.000 to - 0.000) |
| **% Population aged 65+** | | | | 0.035* | 0.039* | | | | 0.032 | 0.046 |
| | | | | (0.007 to 0.063) | (0.008 to 0.070) | | | | (-0.043 to 0.107) | (-0.040 to 0.132) |
| **% Population male** | | | | 0.004 | 0.002 | | | | 0.013 | 0.025 |
| | | | | (-0.018 to 0.025) | (-0.023 to 0.026) | | | | (-0.042 to 0.068) | (-0.032 to 0.082) |
| **Life expectancy at birth (years)** | | | | 0.019 | 0.020 | | | | 0.007 | 0.004 |
| | | | | (-0.001 to 0.038) | (-0.002 to 0.042) | | | | (-0.054 to 0.069) | (-0.061 to 0.069) |

| | | | | | | | | | | |
|---|---|---|---|---|---|---|---|---|---|---|
| **Hospital beds (per 1000 people)** | | | | -0.073** | -0.056* | | | | -0.201*** | -0.174** |
| | | | | (-0.118 to -0.028) | (-0.099 to -0.013) | | | | (-0.311 to -0.090) | (-0.293 to -0.054) |
| **Physicians (per 1000 people)** | | | | 0.035 | 0.001 | | | | 0.057 | 0.056 |
| | | | | (-0.103 to 0.172) | (-0.126 to 0.127) | | | | (-0.201 to 0.316) | (-0.183 to 0.295) |
| **GDP PPP (current international $)** | | | | -0.000 | -0.000 | | | | -0.000* | -0.000* |
| | | | | (-0.000 to 0.000) | (-0.000 to 0.000) | | | | (-0.000 to -0.000) | (-0.000 to -0.000) |
| **Manufacturing value added (%GDP)** | | | | -0.002 | -0.006 | | | | -0.015 | -0.020 |
| | | | | (-0.013 to 0.008) | (-0.018 to 0.005) | | | | (-0.041 to 0.010) | (-0.047 to 0.007) |
| **Health expenditure (%GDP)** | | | | -0.030 | -0.013 | | | | 0.026 | 0.046 |
| | | | | (-0.069 to 0.009) | (-0.053 to 0.027) | | | | (-0.117 to 0.169) | (-0.095 to 0.188) |
| **International tourism, number of arrivals** | | | | 0.000 | 0.000 | | | | 0.000** | 0.000** |
| | | | | (-0.000 to 0.000) | (-0.000 to 0.000) | | | | (0.000 to 0.000) | (0.000 to 0.000) |
| **Governance (Voice and Accountability)** | | | | 0.023 | 0.012 | | | | 0.104 | 0.017 |
| | | | | (-0.129 to 0.176) | (-0.157 to 0.182) | | | | (-0.301 to 0.509) | (-0.417 to 0.451) |
| **East Asia & Pacific** | | | | . | . | | | | . | . |
| **Europe & Central Asia** | | | | 0.484* | 0.276 | | | | 0.944 | 0.621 |
| | | | | (0.027 to 0.942) | (-0.307 to 0.859) | | | | (-0.187 to 2.074) | (-0.587 to 1.829) |

| | | | | | | | | | | |
|---|---|---|---|---|---|---|---|---|---|---|
| **America & Caribbean** | | | | 0.249 | 0.067 | | | | 0.473 | 0.290 |
| | | | | (-0.010 to 0.509) | (-0.252 to 0.385) | | | | (-0.353 to 1.299) | (-0.570 to 1.150) |
| **East & North Africa** | | | | 0.235 | 0.170 | | | | -0.214 | -0.353 |
| | | | | (-0.091 to 0.561) | (-0.230 to 0.570) | | | | (-1.456 to 1.029) | (-1.618 to 0.911) |
| **North America** | | | | 0.217 | -0.092 | | | | -0.018 | -0.456 |
| | | | | (-0.562 to 0.996) | (-0.879 to 0.695) | | | | (-1.721 to 1.684) | (-2.182 to 1.270) |
| **South Asia** | | | | 0.256 | 0.069 | | | | 0.911 | 0.992* |
| | | | | (-0.064 to 0.576) | (-0.236 to 0.374) | | | | (-0.085 to 1.907) | (0.032 to 1.952) |
| **Sub-Saharan Africa** | | | | 0.455** | 0.308 | | | | 0.345 | 0.313 |
| | | | | (0.169 to 0.741) | (-0.011 to 0.627) | | | | (-0.844 to 1.534) | (-0.970 to 1.597) |
| **Constant** | . | 0.659* | 35.122 | -145.905 | -84.805 | | 2.330 | 699.965* | 42.647 | -161.127 |
| | . | (0.143 to 1.174) | (-106.600 to 176.845) | (-302.473 to 10.663) | (-243.949 to 74.339) | | (-0.527 to 5.188) | (91.809 to 1308.121) | (-298.297 to 383.590) | (-592.161 to 269.906) |
| **Observations** | 3200 | 3200 | 3200 | 3200 | 3200 | 3100 | 3100 | 3100 | 3100 | 3100 |
| **R-squared** | | 0.121 | 0.122 | 0.226 | 0.325 | | 0.311 | 0.349 | 0.561 | 0.597 |
| **Adjusted R-squared** | | 0.113 | 0.113 | 0.215 | 0.304 | | 0.304 | 0.343 | 0.554 | 0.584 |

**p-values in parentheses** = * p<0.05 ** p<0.01 *** p<0.001; # includes time, day of week and week of year fixed effects

*Regression results, policy timing. Baseline is policy not introduced within policy analysis period.*

| | **24 Days (Policies implemented before first death)** | | | | | | **38 Days (Policies implemented within 14 days after first death)** | | | | |
|---|---|---|---|---|---|---|---|---|---|---|---|
| **Average deaths per day over time period, per million** | **(1)** | **(2)** | **(3)** | **(4)** | **(5) #** | | **(1)** | **(2)** | **(3)** | **(4)** | **(5) #** |
| **School (Not introduced)** | . | . | . | . | . | **School (Not introduced)** | . | . | . | . | . |
| **School (0-10 days before)** | -0.278 | -0.216 | -0.194 | -0.335* | -0.445** | **School (before)** | -0.670 | -0.313 | 0.194 | -0.436 | -0.865 |
| | (-0.640 to 0.084) | (-0.516 to 0.084) | (-0.500 to 0.113) | (-0.606 to -0.064) | (-0.779 to -0.112) | | (-1.987 to 0.647) | (-1.594 to 0.969) | (-1.102 to 1.491) | (-1.391 to 0.519) | (-2.012 to 0.283) |
| **School (11-20 days before)** | -0.451* | -0.448** | -0.417** | -0.498** | -0.593** | **School (0-7 days after)** | 0.875 | 0.460 | 0.915 | 0.485 | 0.233 |
| | (-0.799 to -0.104) | (-0.766 to -0.131) | (-0.728 to -0.107) | (-0.820 to -0.177) | (-0.969 to -0.218) | | (-0.849 to 2.599) | (-1.175 to 2.095) | (-0.844 to 2.673) | (-0.633 to 1.603) | (-1.050 to 1.516) |
| **School (20+ days before)** | -0.493** | -0.225 | -0.160 | -0.425 | -0.462 | **School (8-14 days after)** | 2.760 | 3.031 | 3.132 | 2.333** | 2.346** |
| | (-0.838 to -0.148) | (-0.577 to 0.126) | (-0.507 to 0.187) | (-0.877 to 0.027) | (-0.964 to 0.039) | | (-1.053 to 6.574) | (-0.251 to 6.313) | (-0.122 to 6.385) | (0.923 to 3.743) | (0.877 to 3.815) |
| **Work (Not introduced)** | . | . | . | . | . | **Work (Not introduced)** | . | . | . | . | . |
| **Work (0-10 days before)** | 0.057 | 0.288* | 0.290* | 0.208* | 0.218* | **Work (before)** | -0.116 | -0.013 | 0.024 | -0.306 | -0.475 |
| | (-0.185 to 0.299) | (0.061 to 0.516) | (0.061 to 0.520) | (0.027 to 0.390) | (0.013 to 0.424) | | (-0.717 to 0.484) | (-0.633 to 0.607) | (-0.558 to 0.607) | (-0.729 to 0.118) | (-0.966 to 0.016) |
| **Work (11-20 days before)** | -0.244* | 0.026 | 0.022 | 0.106 | 0.158 | **Work (0-7 days after)** | 0.147 | -0.631 | -0.675 | -0.839** | -1.098*** |
| | (-0.429 to -0.058) | (-0.273 to 0.326) | (-0.281 to 0.324) | (-0.150 to 0.362) | (-0.154 to 0.469) | | (-0.659 to 0.953) | (-1.649 to 0.387) | (-1.691 to 0.341) | (-1.364 to -0.314) | (-1.723 to -0.474) |
| **Work (20+ days before)** | -0.180 | 0.058 | 0.061 | -0.116 | -0.043 | **Work (8-14 days after)** | 3.616* | 2.246* | 2.107 | 1.860*** | 1.498** |
| | (-0.369 to 0.009) | (-0.168 to 0.284) | (-0.164 to 0.285) | (-0.448 to 0.216) | (-0.399 to 0.313) | | (0.438 to 6.793) | (0.110 to 4.383) | (-0.053 to 4.267) | (0.889 to 2.830) | (0.401 to 2.595) |

| | | | | | | | | | | | |
|---|---|---|---|---|---|---|---|---|---|---|---|
| **Events (Not introduced)** | . | . | . | . | . | **Events (Not introduced)** | . | . | . | . | . |
| **Events (0-10 days before)** | -0.202 | -0.122 | -0.112 | -0.119 | -0.154 | **Events (before)** | -0.421 | -0.330 | -0.518 | -0.192 | 0.148 |
| | (-0.565 to 0.161) | (-0.453 to 0.209) | (-0.443 to 0.219) | (-0.369 to 0.131) | (-0.442 to 0.135) | | (-1.275 to 0.432) | (-1.634 to 0.975) | (-1.755 to 0.720) | (-0.911 to 0.528) | (-0.672 to 0.967) |
| **Events (11-20 days before)** | -0.356* | -0.163 | -0.143 | -0.107 | -0.120 | **Events (0-7 days after)** | 1.351 | 0.050 | -0.184 | 0.243 | 0.790 |
| | (-0.699 to -0.012) | (-0.455 to 0.130) | (-0.443 to 0.157) | (-0.371 to 0.157) | (-0.418 to 0.178) | | (-0.660 to 3.363) | (-1.697 to 1.797) | (-1.918 to 1.549) | (-0.579 to 1.066) | (-0.190 to 1.771) |
| **Events (20+ days before)** | -0.430* | -0.210 | -0.198 | 0.111 | 0.241 | **Events (8-14 days after)** | 2.429 | -0.728 | -0.750 | -0.389 | 0.051 |
| | (-0.765 to -0.095) | (-0.561 to 0.141) | (-0.564 to 0.167) | (-0.324 to 0.546) | (-0.312 to 0.794) | | (-1.080 to 5.938) | (-4.507 to 3.052) | (-4.366 to 2.866) | (-1.745 to 0.968) | (-1.514 to 1.617) |
| **Gatherings (Not introduced)** | . | . | . | . | . | **Gatherings (Not introduced)** | . | . | . | . | . |
| **Gatherings (0-10 days before)** | -0.128 | 0.048 | 0.047 | 0.148 | 0.144 | **Gatherings (before)** | -1.144 | -0.063 | -0.159 | 0.699* | 0.757* |
| | (-0.403 to 0.148) | (-0.129 to 0.224) | (-0.129 to 0.224) | (-0.046 to 0.342) | (-0.071 to 0.359) | | (-2.497 to 0.208) | (-0.693 to 0.568) | (-0.817 to 0.499) | (0.142 to 1.255) | (0.144 to 1.369) |
| **Gatherings (11-20 days before)** | -0.243 | 0.097 | 0.090 | 0.178 | 0.167 | **Gatherings (0-7 days after)** | -0.095 | 0.443 | 0.196 | 0.770* | 0.744 |
| | (-0.495 to 0.009) | (-0.121 to 0.316) | (-0.127 to 0.307) | (-0.077 to 0.433) | (-0.123 to 0.456) | | (-1.685 to 1.495) | (-0.516 to 1.402) | (-0.826 to 1.218) | (0.052 to 1.487) | (-0.068 to 1.556) |
| **Gatherings (20+ days before)** | -0.291* | 0.085 | 0.091 | -0.007 | 0.028 | **Gatherings (8-14 days after)** | -0.889 | -3.935* | -4.054* | -3.937*** | -4.037*** |
| | (-0.523 to -0.059) | (-0.219 to 0.389) | (-0.222 to 0.404) | (-0.353 to 0.339) | (-0.356 to 0.413) | | (-2.412 to 0.634) | (-7.158 to -0.713) | (-7.155 to -0.954) | (-5.261 to -2.613) | (-5.487 to -2.587) |
| **Transport (Not introduced)** | . | . | . | . | . | **Transport (Not introduced)** | . | . | . | . | . |
| **Transport (0-10 days before)** | 0.049 | 0.032 | 0.025 | 0.086 | 0.023 | **Transport (before)** | -0.434 | -0.104 | -0.077 | 0.341* | 0.271 |

| | | | | | | | | | | | |
|---|---|---|---|---|---|---|---|---|---|---|---|
| | (-0.220 to 0.317) | (-0.215 to 0.279) | (-0.221 to 0.271) | (-0.138 to 0.309) | (-0.223 to 0.269) | | (-0.961 to 0.093) | (-0.469 to 0.261) | (-0.434 to 0.279) | (0.010 to 0.671) | (-0.064 to 0.606) |
| **Transport (11-20 days before)** | -0.219** | -0.079 | -0.088 | 0.051 | -0.021 | **Transport (0-7 days after)** | -0.381 | 0.166 | 0.284 | 0.152 | 0.135 |
| | (-0.360 to -0.078) | (-0.371 to 0.212) | (-0.388 to 0.212) | (-0.286 to 0.388) | (-0.394 to 0.352) | | (-0.943 to 0.181) | (-0.238 to 0.571) | (-0.143 to 0.711) | (-0.222 to 0.525) | (-0.279 to 0.550) |
| **Transport (20+ days before)** | -0.162* | -0.034 | -0.054 | 0.538 | 0.276 | **Transport (8-14 days after)** | 1.975 | 1.399 | 1.298 | 1.702** | 1.555* |
| | (-0.309 to -0.014) | (-0.388 to 0.320) | (-0.410 to 0.302) | (-0.024 to 1.100) | (-0.303 to 0.855) | | (-2.825 to 6.774) | (-1.143 to 3.941) | (-1.221 to 3.817) | (0.433 to 2.972) | (0.317 to 2.794) |
| **Stay home (Not introduced)** | . | . | . | . | . | **Stay home (Not introduced)** | . | . | . | . | . |
| **Stay home (0-10 days before)** | -0.085 | -0.086 | -0.092 | -0.158 | -0.196 | **Stay home (before)** | -0.507 | 0.230 | 0.202 | -0.403 | -0.472* |
| | (-0.276 to 0.106) | (-0.350 to 0.177) | (-0.357 to 0.174) | (-0.401 to 0.085) | (-0.474 to 0.082) | | (-1.178 to 0.164) | (-0.434 to 0.894) | (-0.479 to 0.883) | (-0.829 to 0.022) | (-0.937 to -0.008) |
| **Stay home (11-20 days before)** | -0.177 | -0.071 | -0.070 | -0.210 | -0.195 | **Stay home (0-7 days after)** | -0.033 | -0.121 | -0.031 | -0.187 | -0.208 |
| | (-0.415 to 0.061) | (-0.462 to 0.320) | (-0.461 to 0.321) | (-0.543 to 0.122) | (-0.534 to 0.143) | | (-0.912 to 0.846) | (-0.853 to 0.612) | (-0.730 to 0.669) | (-0.724 to 0.350) | (-0.795 to 0.378) |
| **Stay home (20+ days before)** | -0.256** | -0.150 | -0.181 | -0.589* | -0.523* | **Stay home (8-14 days after)** | 0.831 | 0.292 | 0.385 | -0.273 | -0.447 |
| | (-0.429 to -0.083) | (-0.573 to 0.274) | (-0.605 to 0.244) | (-1.109 to -0.070) | (-1.045 to -0.002) | | (-1.192 to 2.855) | (-0.654 to 1.239) | (-0.525 to 1.295) | (-0.792 to 0.246) | (-1.038 to 0.145) |
| **Internal movement (Not introduced)** | . | . | . | . | . | **Internal movement (Not introduced)** | . | . | . | . | . |
| **Internal movement (0-10 days before)** | -0.117 | -0.062 | -0.052 | -0.095 | -0.085 | **Internal movement (before)** | -0.425 | 0.200 | 0.232 | 0.269 | 0.312 |
| | (-0.299 to 0.065) | (-0.269 to 0.146) | (-0.261 to 0.156) | (-0.278 to 0.089) | (-0.281 to 0.111) | | (-1.006 to 0.156) | (-0.355 to 0.755) | (-0.318 to 0.782) | (-0.073 to 0.611) | (-0.040 to 0.664) |

| | | | | | | | | | | |
|---|---|---|---|---|---|---|---|---|---|---|
| **Internal movement (11-20 days before)** | -0.051 | 0.121 | 0.138 | 0.129 | 0.180 | **Internal movement (0-7 days after)** | -0.185 | 0.601 | 0.519 | 0.223 | 0.139 |
| | (-0.297 to 0.196) | (-0.196 to 0.439) | (-0.188 to 0.464) | (-0.108 to 0.367) | (-0.078 to 0.437) | | (-0.894 to 0.524) | (-0.143 to 1.345) | (-0.197 to 1.234) | (-0.373 to 0.819) | (-0.507 to 0.786) |
| **Internal movement (20+ days before)** | -0.279*** | 0.070 | 0.107 | 0.181 | 0.372 | **Internal movement (8-14 days after)** | 1.186 | 0.109 | 0.201 | 0.192 | 0.182 |
| | (-0.441 to -0.118) | (-0.210 to 0.351) | (-0.180 to 0.394) | (-0.214 to 0.577) | (-0.111 to 0.855) | | (-0.405 to 2.778) | (-0.922 to 1.139) | (-0.852 to 1.253) | (-0.440 to 0.824) | (-0.551 to 0.915) |
| **Travel (Not introduced)** | . | . | . | . | . | **Travel (Not introduced)** | . | . | . | . | . |
| **Travel (0-10 days before)** | -0.766* | -0.707* | -0.696* | -0.263 | -0.350 | **Travel (before)** | -4.078*** | -2.998* | -2.990* | -2.247*** | -2.322*** |
| | (-1.453 to -0.079) | (-1.344 to -0.070) | (-1.330 to -0.062) | (-0.747 to 0.221) | (-0.925 to 0.225) | | (-6.390 to -1.767) | (-5.294 to -0.703) | (-5.259 to -0.721) | (-3.183 to -1.310) | (-3.321 to -1.323) |
| **Travel (11-20 days before)** | -0.783* | -0.640* | -0.639* | -0.179 | -0.233 | **Travel (0-7 days after)** | -1.704 | -2.125 | -2.203 | -2.406*** | -2.311*** |
| | (-1.468 to -0.099) | (-1.272 to -0.009) | (-1.272 to -0.007) | (-0.650 to 0.293) | (-0.767 to 0.302) | | (-5.362 to 1.954) | (-4.485 to 0.236) | (-4.519 to 0.112) | (-3.309 to -1.503) | (-3.175 to -1.448) |
| **Travel (20+ days before)** | -0.889** | -0.898* | -0.893* | -0.468 | -0.537 | **Travel (8-14 days after)** | -1.258 | 1.000 | 1.071 | 1.251 | 1.123 |
| | (-1.558 to -0.221) | (-1.606 to -0.190) | (-1.599 to -0.188) | (-1.049 to 0.113) | (-1.172 to 0.098) | | (-6.255 to 3.738) | (-3.917 to 5.917) | (-3.877 to 6.019) | (-1.268 to 3.770) | (-1.310 to 3.556) |
| **Public information (Not introduced)** | . | . | . | . | . | **Public information (Not introduced)** | . | . | . | . | . |
| **Public information (0-10 days before)** | 0.188 | 0.226 | 0.231 | 0.463 | 0.435 | **Public information (before)** | 0.039 | 0.962 | 1.397 | 0.868 | 0.961 |
| | (-0.386 to 0.763) | (-0.285 to 0.736) | (-0.284 to 0.745) | (-0.110 to 1.035) | (-0.164 to 1.035) | | (-0.775 to 0.854) | (-1.964 to 3.888) | (-1.653 to 4.446) | (-1.324 to 3.061) | (-1.243 to 3.165) |
| **Public information (11-20 days before)** | -0.015 | 0.375 | 0.370 | 0.345 | 0.362 | **Public information** | 0.884 | -0.522 | -0.093 | -0.353 | -0.167 |

| | | | | | | | | | | | |
|---|---|---|---|---|---|---|---|---|---|---|---|
| | | | | | | **(0-7 days after)** | | | | | |
| | (-0.378 to 0.348) | (-0.238 to 0.988) | (-0.240 to 0.979) | (-0.265 to 0.955) | (-0.266 to 0.989) | | (-1.054 to 2.821) | (-3.813 to 2.770) | (-3.516 to 3.330) | (-2.596 to 1.889) | (-2.435 to 2.101) |
| **Public information (20+ days before)** | -0.018 | 0.453 | 0.463 | 0.457 | 0.453 | **Public information (8-14 days after)** | -0.383 | 1.741 | 2.140 | -0.597 | -0.366 |
| | (-0.376 to 0.340) | (-0.198 to 1.105) | (-0.194 to 1.120) | (-0.225 to 1.138) | (-0.219 to 1.124) | | (-1.182 to 0.415) | (-2.295 to 5.777) | (-1.910 to 6.189) | (-3.253 to 2.059) | (-3.107 to 2.375) |
| **Testing (Not introduced)** | . | . | . | . | . | **Testing (Not introduced)** | . | . | . | . | . |
| **Testing (0-10 days before)** | 0.050 | -0.089 | -0.085 | -0.154 | -0.233 | **Testing (before)** | 0.449* | 0.785 | 0.705 | 0.608* | 0.474 |
| | (-0.138 to 0.238) | (-0.398 to 0.220) | (-0.387 to 0.217) | (-0.481 to 0.172) | (-0.614 to 0.147) | | (0.001 to 0.897) | (-0.060 to 1.630) | (-0.165 to 1.576) | (0.037 to 1.179) | (-0.146 to 1.094) |
| **Testing (11-20 days before)** | 0.249* | -0.020 | -0.019 | -0.018 | -0.057 | **Testing (0-7 days after)** | 0.041 | 0.897 | 0.666 | 0.644 | 0.472 |
| | (0.000 to 0.499) | (-0.379 to 0.339) | (-0.374 to 0.336) | (-0.343 to 0.307) | (-0.425 to 0.310) | | (-0.405 to 0.487) | (-0.268 to 2.062) | (-0.443 to 1.775) | (-0.328 to 1.616) | (-0.672 to 1.616) |
| **Testing (20+ days before)** | 0.190 | 0.015 | 0.008 | 0.034 | -0.011 | **Testing (8-14 days after)** | -0.133 | 1.365 | 1.174 | 1.348** | 1.581* |
| | (-0.001 to 0.382) | (-0.268 to 0.297) | (-0.278 to 0.295) | (-0.244 to 0.313) | (-0.319 to 0.296) | | (-0.448 to 0.183) | (-0.506 to 3.237) | (-0.779 to 3.127) | (0.414 to 2.282) | (0.349 to 2.813) |
| **Contact tracing (Not introduced)** | . | . | . | . | . | **Contact tracing (Not introduced)** | . | . | . | . | . |
| **Contact tracing (0-10 days before)** | 0.078 | 0.156 | 0.176 | 0.305* | 0.389** | **Contact tracing (before)** | 0.498* | 0.629 | 0.539 | 0.492* | 0.439 |
| | (-0.138 to 0.294) | (-0.135 to 0.446) | (-0.111 to 0.463) | (0.051 to 0.559) | (0.099 to 0.678) | | (0.044 to 0.953) | (-0.054 to 1.312) | (-0.146 to 1.225) | (0.051 to 0.934) | (-0.053 to 0.931) |
| **Contact tracing (11-20 days before)** | 0.275* | 0.367* | 0.375* | 0.286* | 0.330* | **Contact tracing (0-7 days after)** | -0.098 | 0.446 | 0.414 | 0.682 | 0.478 |
| | (0.027 to 0.523) | (0.068 to 0.666) | (0.079 to 0.671) | (0.002 to 0.570) | (0.033 to 0.628) | | (-0.403 to 0.207) | (-0.931 to 1.823) | (-0.962 to 1.789) | (-0.304 to 1.668) | (-0.604 to 1.561) |

| | | | | | | | | | | | |
|---|---|---|---|---|---|---|---|---|---|---|---|
| **Contact tracing (20+ days before)** | 0.153 | 0.317* | 0.290 | 0.232 | 0.221 | **Contact tracing (8-14 days after)** | -0.178 | 1.578* | 1.583* | 0.871* | 1.023** |
| | (-0.075 to 0.381) | (0.019 to 0.614) | (-0.007 to 0.588) | (-0.024 to 0.489) | (-0.017 to 0.458) | | (-0.490 to 0.134) | (0.030 to 3.126) | (0.088 to 3.079) | (0.122 to 1.620) | (0.281 to 1.765) |
| **Date of first case** | | | -0.002 | 0.006 | 0.005 | | | | -0.014 | 0.004 | 0.000 |
| | | | (-0.007 to 0.003) | (-0.001 to 0.012) | (-0.003 to 0.012) | | | | (-0.030 to 0.002) | (-0.009 to 0.016) | (-0.015 to 0.015) |
| **Population density (people per sq.km)** | | | | -0.000 | -0.000 | | | | | -0.000 | -0.000 |
| | | | | (-0.000 to 0.000) | (-0.000 to 0.000) | | | | | (-0.000 to 0.000) | (-0.000 to 0.000) |
| **% Population aged 65+** | | | | 0.038* | 0.048* | | | | | 0.027 | 0.025 |
| | | | | (0.005 to 0.071) | (0.010 to 0.086) | | | | | (-0.042 to 0.095) | (-0.052 to 0.103) |
| **% Population male** | | | | 0.013 | 0.016 | | | | | 0.035 | 0.041 |
| | | | | (-0.007 to 0.033) | (-0.007 to 0.038) | | | | | (-0.009 to 0.080) | (-0.005 to 0.087) |
| **Life expectancy at birth (years)** | | | | 0.015 | 0.018 | | | | | 0.014 | 0.019 |
| | | | | (-0.003 to 0.033) | (-0.002 to 0.037) | | | | | (-0.034 to 0.062) | (-0.032 to 0.070) |
| **Hospital beds (per 1000 people)** | | | | -0.072** | -0.053* | | | | | -0.213*** | -0.167** |
| | | | | (-0.116 to -0.028) | (-0.102 to -0.004) | | | | | (-0.323 to -0.104) | (-0.280 to -0.053) |
| **Physicians (per 1000 people)** | | | | 0.046 | 0.009 | | | | | 0.269* | 0.236* |
| | | | | (-0.064 to 0.155) | (-0.100 to 0.117) | | | | | (0.028 to 0.509) | (0.015 to 0.458) |
| **GDP PPP (current international $)** | | | | -0.000 | -0.000 | | | | | -0.000 | 0.000 |
| | | | | (-0.000 to 0.000) | (-0.000 to 0.000) | | | | | (-0.000 to 0.000) | (-0.000 to 0.000) |

| | | | | | | | | | | | |
|---|---|---|---|---|---|---|---|---|---|---|---|
| **Manufacturing value added (%GDP)** | | | | -0.006 | -0.011 | | | | | -0.013 | -0.017 |
| | | | | (-0.019 to 0.007) | (-0.026 to 0.005) | | | | | (-0.032 to 0.005) | (-0.038 to 0.004) |
| **Health expenditure (%GDP)** | | | | -0.048 | -0.030 | | | | | -0.024 | 0.012 |
| | | | | (-0.098 to 0.003) | (-0.075 to 0.015) | | | | | (-0.110 to 0.061) | (-0.082 to 0.107) |
| **International tourism, number of arrivals** | | | | 0.000 | 0.000 | | | | | 0.000*** | 0.000*** |
| | | | | (-0.000 to 0.000) | (-0.000 to 0.000) | | | | | (0.000 to 0.000) | (0.000 to 0.000) |
| **Governance (Voice and Accountability)** | | | | 0.012 | -0.027 | | | | | 0.097 | 0.080 |
| | | | | (-0.134 to 0.158) | (-0.200 to 0.146) | | | | | (-0.180 to 0.375) | (-0.241 to 0.402) |
| **East Asia & Pacific** | | | | . | . | | | | | . | . |
| **Europe & Central Asia** | | | | 0.453* | 0.212 | | | | | 1.389*** | 0.966* |
| | | | | (0.073 to 0.832) | (-0.302 to 0.726) | | | | | (0.668 to 2.111) | (0.213 to 1.719) |
| **America & Caribbean** | | | | 0.198 | -0.019 | | | | | 0.935* | 0.648 |
| | | | | (-0.058 to 0.454) | (-0.335 to 0.297) | | | | | (0.205 to 1.666) | (-0.098 to 1.395) |
| **East & North Africa** | | | | 0.210 | 0.106 | | | | | 0.043 | -0.127 |
| | | | | (-0.094 to 0.515) | (-0.249 to 0.460) | | | | | (-0.698 to 0.784) | (-0.914 to 0.660) |
| **North America** | | | | 0.346 | 0.089 | | | | | -1.244 | -2.057 |
| | | | | (-0.450 to 1.143) | (-0.689 to 0.868) | | | | | (-3.010 to 0.521) | (-4.166 to 0.051) |
| **South Asia** | | | | 0.307 | 0.238 | | | | | 1.715*** | 1.703** |

| | | | | | | | | | | | |
|---|---|---|---|---|---|---|---|---|---|---|---|
| | | | | (-0.099 to 0.713) | (-0.154 to 0.631) | | | | | (0.755 to 2.675) | (0.584 to 2.823) |
| **Sub-Saharan Africa** | | | | 0.463** | 0.388* | | | | | 1.339** | 1.163* |
| | | | | (0.164 to 0.763) | (0.046 to 0.730) | | | | | (0.370 to 2.309) | (0.130 to 2.196) |
| **Constant** | | 0.653* | 46.889 | -123.537 | -106.864 | | | 1.365 | 311.603 | -85.252 | -9.557 |
| | | (0.077 to 1.230) | (-62.125 to 155.904) | (-261.536 to 14.462) | (-273.767 to 60.039) | | | (-2.643 to 5.373) | (-30.794 to 654.000) | (-365.136 to 194.633) | (-340.999 to 321.884) |
| **Observations** | 3200 | 3200 | 3200 | 3200 | 3200 | | 3100 | 3100 | 3100 | 3100 | 3100 |
| **R-squared** | | 0.132 | 0.132 | 0.236 | 0.334 | | | 0.441 | 0.447 | 0.647 | 0.683 |
| **Adjusted R-squared** | | 0.122 | 0.123 | 0.224 | 0.313 | | | 0.434 | 0.441 | 0.641 | 0.673 |

**p-values in parentheses =** * p<0.05 ** p<0.01 *** p<0.001; # includes time, day of week and week of year fixed effects

### References

1. European Observatory. How comparable is COVID-19 mortality across countries? 2020. https://analysis.covid19healthsystem.org/index.php/2020/06/04/how-comparable-is-covid-19-mortality-across-countries/ (accessed 18/06/2020.
